# Cross-dataset pan-cancer detection: Correlating cell-free DNA fragment coverage with open chromatin sites across cell types

**DOI:** 10.1101/2024.11.26.24317971

**Authors:** Ludvig Renbo Olsen, Denis Odinokov, Jakob Qvortrup Holsting, Karoline Kondrup, Laura Iisager, Maria Rusan, Simon Buus, Britt Elmedal Laursen, Michael Borre, Mads Ryø Jochumsen, Kirsten Bouchelouche, Amanda Frydendahl, Mads Heilskov Rasmussen, Tenna Vesterman Henriksen, Marijana Nesic, Christina Demuth, Sia Viborg Lindskrog, Iver Nordentoft, Philippe Lamy, Christina Therkildsen, Lars Dyrskjøt, Karina Dalsgaard Sørensen, Claus Lindbjerg Andersen, Anders Jakobsen Skanderup, Søren Besenbacher

## Abstract

The fragmentation patterns of whole genome sequenced cell-free DNA are promising features for tumor-agnostic cancer detection. However, systematic biases challenge their cross-cohort generalization. We introduce LIONHEART, a novel, open source cancer detection method specifically optimized to generalize across datasets. The method correlates bias-corrected cfDNA fragment coverage across the genome with the locations of accessible chromatin regions from 898 cell and tissue type features. We use these correlations to detect changes in the cell-free DNA cell type composition caused by cancer. We test LIONHEART on nine datasets and fourteen cancer types (1106 non-cancer controls, 1449 cancers) obtained from different studies and show that it can distinguish cancer samples from non-cancer controls across cohorts with ROC AUC scores ranging from 0.62-0.95 (mean = 0.83, std = 0.12). We further validate the method on an external dataset, achieving a ROC AUC of 0.917.

## Introduction

Sequencing of plasma cell-free DNA (cfDNA) offers a cost-effective and non-invasive strategy for early cancer detection. A method to detect low amounts of circulating tumor DNA (ctDNA) with high specificity and without prior knowledge of the tumor’s molecular characteristics could radically improve cancer screening. Fragmentomics, which analyzes the sequence, size, and genomic position of cfDNA fragments, is one promising approach for such tumor-agnostic detection of ctDNA^1^.

The fragmentation patterns of cfDNA differ systematically by the cell type and release mechanism, enabling us to infer changes in the contributions of different cell types to the cfDNA. Studies estimating the tissue of origin of cfDNA in healthy individuals agree that most fragments originate from hematopoietic cells with minor contributions from vascular endothelial cells and hepatocytes, although there is no consensus on the exact proportions of these components^2,3^. Many common solid tumor types shed noticeable amounts of tumor DNA to the circulation^1,4^, making ctDNA a promising biomarker for cancer detection. The mechanisms by which DNA fragments enter the bloodstream include both simple release from cell death (apoptosis, necrosis, and NETosis) and active secretion^5^. Multiple nucleases contribute to the fragmentation of cfDNA. During apoptosis, endonucleases like DFFB and DNASE1L3 become active and begin to cleave the DNA between the nucleosomes. After release from cells, the cfDNA is further fragmented by nucleases in the extracellular environment, such as DNASE1 and DNASE1L3^5,6^. The exact cleavage position depends on multiple factors, including which endonucleases are present^7^, whether the DNA is methylated^8^, and whether nucleosomes protect the DNA^9^. DNA wrapped around nucleosomes resists early fragmentation, delaying its breakdown in the bloodstream. Regions of accessible chromatin that allow the binding of transcription-modulating proteins are characterized by nucleosome depletion and, consequently, faster fragment breakdown. As a result, the blood contains fewer cfDNA fragments from the accessible than inaccessible genomic regions. Since the chromatin accessibility profile varies across cell types, reflecting their transcriptional activities, the presence of a given cell type is reflected by reduced fragment coverage at specific locations in the genome. When ctDNA fragments dilute the cfDNA mixture, we thus expect lower sequencing coverage in the tumor-tissue-specific accessible chromatin regions and vice versa for the typically present cell types, like hematopoietic cells.

Since cfDNA fragmentation patterns reflect a fragment’s tissue of origin, they are promising features for tumor-agnostic cancer detection. In practice, however, identifying these patterns in sequencing data is not trivial. Preanalytical (storage, cfDNA extraction protocols, NGS library preparation, and amplification) and analytical (sequencing platform) factors systematically bias and obscure the fragmentation patterns^10,11^. Even after correcting these biases, the vast size of the human genome (∼3Gbp) and the low sample size of most studies mean some regions will differ between cancer and non-cancer samples simply by chance. This multitude of possible features relative to current sample sizes, combined with the technical biases, offers severe risks to the generalization (i.e., the ability to perform similarly well on unseen data) of fragmentomics methods.

To better identify real cancer signals consistent across patients, fragmentomics analyses can be informed by non-cfDNA reference knowledge. Previous studies have used public non-cfDNA datasets as a reference when extracting nucleosome positioning patterns in cfDNA for cancer detection purposes. For instance, Ulz et al. and Doebley et al. detected transcription factor activity from sequencing coverage around binding sites specified in public databases^12,13^, which Sarkar et al. and Rao et al. used to subtype samples within various types of cancers^14,15^. Stanley et al. ranked the proportions of 456 cell types in cfDNA by correlating coverage-based features to their single-cell RNA-sequencing measures^16^. Zhu et al. used RNA-sequencing data to identify a small number of promoters and exon-intron junctions where cfDNA coverage could be leveraged to predict ctDNA burden^17^.

Besides gene expression data and databases of known binding sites, there are more direct functional assays of open chromatin, such as DNase and ATAC sequencing, that measure the chromatin accessibility in a given cell or tissue sample. We can use such data to inform our expectations of the fragmentation patterns, given the presence of different cell types. By comparing how the cfDNA sequencing coverage correlates with such reference knowledge in cases and controls, we may identify more robust cancer-relevant signals.

A few studies have attempted to deal with confounding biases in cfDNA. Wan et al. used different confounder-based cross-validation variations to explicitly reduce the effect of confounders on classification^18^. Jamshidi et al. reduced the systematic biases identified in the first five principal components of the variation within the control samples^19^. Passemiers et al. corrected for pre-analytical variables using optimal transport-based domain adaption^20^. Yet, as most published methods are only validated within-dataset or on a single external dataset, it is unclear whether current methods generalize across labs and relevant demographical variables. In this study, we develop a novel pan-cancer detection method that generalizes across ten datasets and fourteen cancer types (Fig. 1).

**Figure 1:**
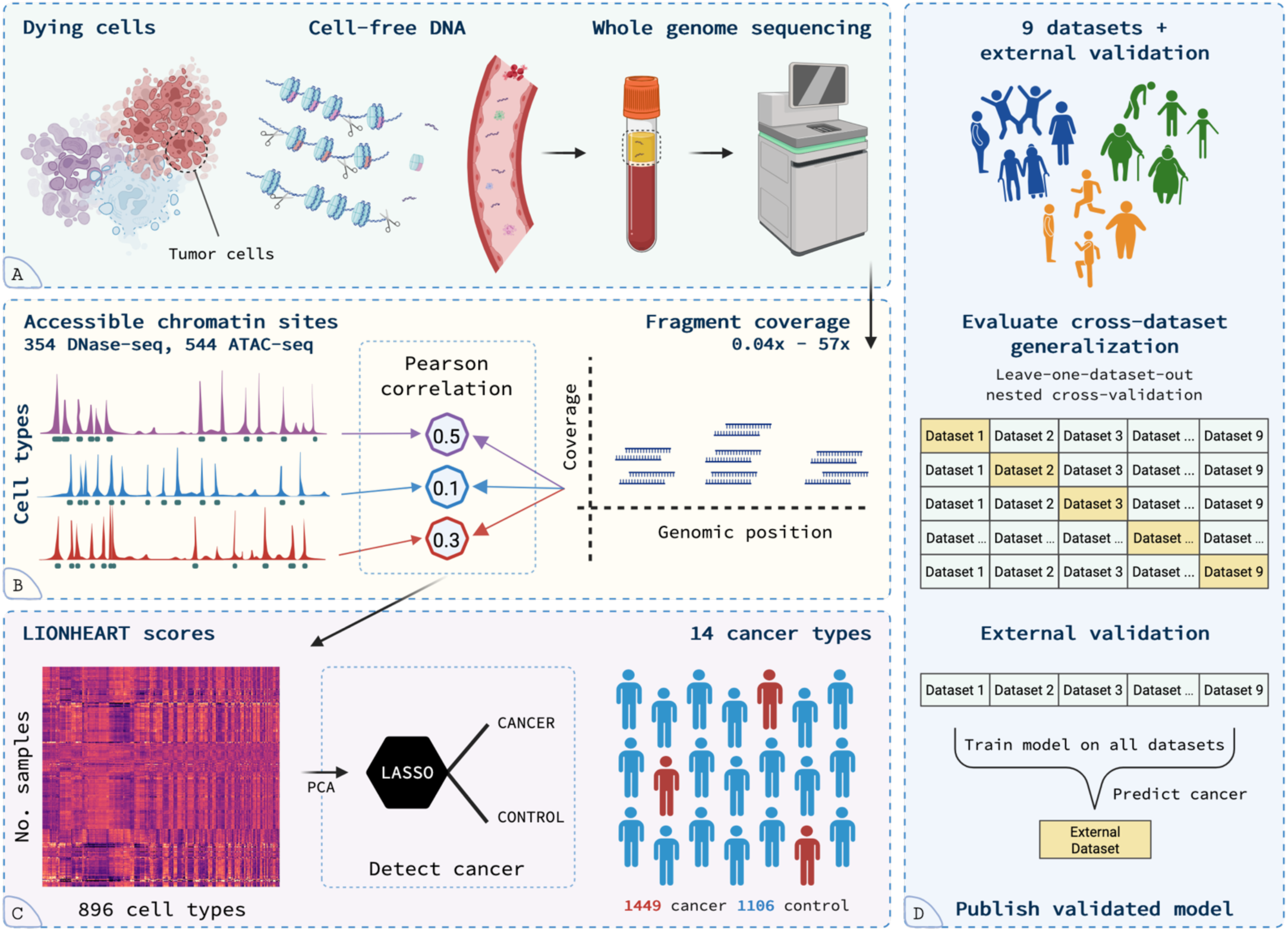
Simplified overview of the study: A) Dying cells release cell-free DNA fragments into the bloodstream. The fragments are extracted from the plasma and whole genome sequenced. B) The fragment coverage is counted in 10bp bins and correlated to the accessible chromatin locations of 896 cell types*. C) The correlation coefficients are PCA-transformed and fed to a LASSO logistic regression model that classifies samples as either cancer or control. D) The cross-dataset generalization of the model is estimated with leave-one-dataset-out nested cross-validation for nine datasets. Finally, a model is trained on all nine datasets and validated on independent external data. This model is made available to the community. **Not illustrated**: Bias correction of the fragment coverage. Sample-and cell-type-standardizations of the correlation coefficients. *Two consensus-site features are included as well (Methods). Created in BioRender: https://BioRender.com/mymrqn0

## Results

Sequencing coverage is reduced in regions where the cfDNA-contributing cells have accessible chromatin. To investigate whether this signal reveals cancer-relevant changes in the cfDNA cell type composition, we collected publicly available chromatin accessibility datasets for 700 unique cell and tissue types, represented by 898 features (354 DNase-seq and 544 ATAC-seq). For each of these 898 cell and tissue type features (hereafter, just “cell types”), we calculated the Pearson correlation between the bias-corrected genome-wide sequencing coverage and the accessible chromatin positions in 10bp bins and standardized the correlation coefficients for each cfDNA sample (Fig. 1B). We name these sample-standardized correlation coefficients LIONHEART scores (**li**quid b**i**opsy c**<u>o</u>**rrelati**<u>n</u>**g c**<u>h</u>**romatin acc**<u>e</u>**ssibility and cfDN**<u>A</u>** cove**<u>r</u>**age across cell **<u>t</u>**ypes). We expect cell types that comprise a large proportion of the cfDNA to have a *lower* LIONHEART score (i.e., a correlation closer to -1.0 (pre-standardization) between coverage and accessible chromatin) than less abundant cell types; this means that we expect healthy individuals to have lower LIONHEART scores for hematopoietic cell types than other cell types. Investigating cfDNA from 244 healthy donors from Cristiano et al.^21^, we observed such lower scores for T-cells, cd14^+^ monocytes, natural killer cells, and B-cells compared to lung, tongue, retina, and brain cell types (Fig. 2A). With the introduction of tumor DNA, the *proportion* of the hematopoietic cell types decrease. We thus expect the LIONHEART scores for the hematopoietic cell types to increase (i.e., correlations closer to 1.0, pre-standardization). We confirmed this by correlating the LIONHEART scores of typically present hematopoietic cell types against the ichorCNA-estimated tumor fractions in a cohort of 361 prostate cancer cases (337 metastatic, 24 localized): B-cells (r = 0.73 for ATAC-seq; r = 0.64 for DNase-seq), cd14^+^ monocytes (r = 0.77 for DNase-seq), cd4^+^ αβ T-cells (r = 0.53 for ATAC-seq; r = 0.39 for DNase-seq), cd8^+^ αβ T-cells (r = 0.59 for ATAC-seq; r = 0.38 for DNase-seq), and natural killer cells (r = 0.76 for ATAC-seq; r = 0.67 for DNase-seq) (Fig. 2B). For cell types that are more abundant in the tumor, we expect the reverse relationship. We confirmed this for the two prostate cancer cell lines, PC3 (r = -0.82; DNase-seq) and PRAD (r = -0.91; ATAC-seq) (Fig. 2C), as well as for non-cancerous prostate gland tissue (r = -0.87 for ATAC-seq; r = -0.68 for DNase-seq). PRAD had the most negative correlation of all cell types, but other non-prostate cell types also had highly negative correlations, like tissue from the left colon (r = - 0.87; ATAC-seq) and the MCF-7 breast cancer cell line (r = -0.90; ATAC-seq).

**Figure 2:**
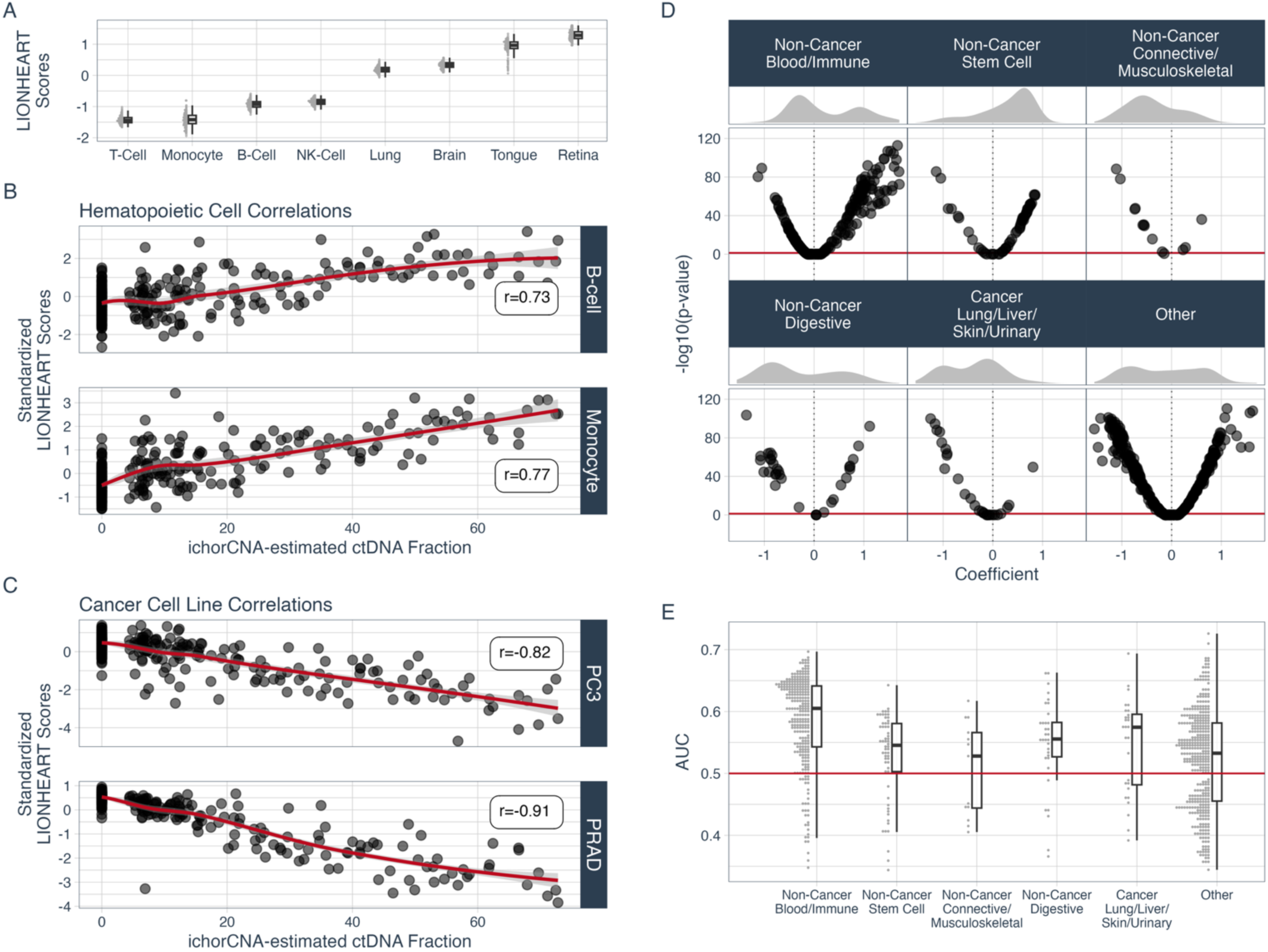
Univariate analyses of LIONHEART scores. A) The calculated LIONHEART scores for different cell types in 244 non-cancer control samples from Cristiano et al.^21^. The cell types expected to contribute to cfDNA in healthy individuals (t-cells, cd14+ monocytes, b-cells and natural killer cells) have lower LIONHEART scores than cell types not expected to contribute to cfDNA in healthy individuals (lung, brain, tongue, and retina). B) The correlations between the ichorCNA-estimated ctDNA fraction and the cell-type-standardized LIONHEART scores for the ATAC-sequenced B-cell (top; r=0.73) and DNase-sequenced cd14^+^ monocytes (bottom; r=0.77). Each point is a cancer sample from the Prostate Cancer (Aarhus cohorts) dataset. C) The correlations between cell-type-standardized LIONHEART scores for the PC3 (top; r=0.82; DNase-seq) and PRAD (bottom; r=0.91; ATAC-seq) prostate cancer cell lines and the ichorCNA-estimated ctDNA fraction. Each point is a cancer sample from the Prostate Cancer (Aarhus cohorts) dataset. D) Logistic regression coefficients and their Bonferroni-corrected negative log-p-values from univariate models classifying “Control (0) vs. Cancer (1)” on all datasets combined. The cell types are grouped into five relevant categories plus an “Other” category for the remaining cell types. Each point is a model coefficient for a single cell type. **Top distributions**: The densities of samples. **Horizontal line**: Significance threshold. **Positive coefficients**: A lower relative presence of the cell type indicates cancer. **Negative coefficients**: A higher relative presence of the cell type indicates cancer. E) Leave-one-dataset-out cross-validated ROC AUC scores from univariate cancer detection models. The cell types are grouped into five relevant categories plus an “Other” category for the remaining cell types. Each point is an AUC score for a single cell type. **Horizontal line**: Chance level. Cell types scoring below the chance level had reverse relationships to the cancer status in the different datasets.

### Pan-cancer vs. non-cancer controls

We included nine whole genome sequenced (WGS) cfDNA datasets comprising 1449 cfDNA samples from patients with one of 14 cancer types and 1106 samples from non-cancer controls (Supp. Table 1). The Prostate Cancer (Aarhus cohorts) dataset contained only 18 controls and was thus mainly used for model training, leaving eight test datasets.

To explore the LIONHEART scores across all the cfDNA samples from the datasets, we fitted a univariate pan-cancer vs. non-cancer-control logistic regression model for each cell type. We then categorized the cell types by their tissue-/organ-system (blood/immune, digestive system, lungs, liver, etc.) and inspected the general trends of the model coefficients per category (Fig. 2D; Supp. Fig. 1). The blood/immune cell types had mostly positive model coefficients, indicating that a lower proportion of blood-related cells is a sign of cancer. In contrast, the cancer-derived lung, liver, skin, and urinary cell types had mostly negative model coefficients, unsurprisingly indicating that a higher proportion of cancer cells from these tissues/organs is a sign of cancer. In total, 777 of the 898 cell types had significantly (α = 0.05, Bonferroni-corrected) non-zero coefficients. The cell types that were best at classifying cancer presence achieved leave-one-dataset-out cross-validated ROC AUCs up to 0.73 (Fig. 2E).

We hypothesized that combining these many weak predictors would likely lead to a better predictive model. Thus, we constructed a pan-cancer vs. non-cancer-control LASSO logistic regression classifier using the LIONHEART scores of all the cell types (Fig. 3A-3B). Using nested cross-validation, we trained and evaluated a separate LASSO classifier for each dataset. This achieved an average within-dataset ROC AUC of 0.852 (0.872 if weighted by dataset size; hereafter just “weighted”). To assess whether or not the classifier generalizes to completely unseen cohorts (i.e., cross-dataset generalization), we then ran leave-one-dataset-out nested cross-validation, where, for each test dataset, the classifier was first trained on the eight other datasets and then predicted the cancer statuses in the remaining test dataset. This resulted in an average cross-dataset ROC AUC of 0.826 (weighted: 0.850). For two of the datasets, the classifier achieved higher ROC AUC scores cross-dataset than within-dataset, namely the Endoscopy II cohort (within-dataset: 0.602, cross-dataset: 0.728) and the dataset from Mathios et al.^22^ (within-dataset: 0.735, cross-dataset: 0.787). Supplementary Figure 2 shows the ROC curves per cancer stage and dataset. Further, to compare the LIONHEART scores to other fragmentomics features that can be extracted from WGS cfDNA data, we evaluated the cross-dataset generalization of LASSO logistic regression classifiers based on the fragment length distributions (average AUC: 0.729, weighted: 0.736), the ratios of short to long fragments in 1Mb bins (average AUC: 0.801, weighted: 0.812), and the average coverage in 1Mb bins (average AUC: 0.713, weighted: 0.739) (Fig. 3A-3B; Supp. Table 2). Supplementary Table 3 has the ROC AUC scores per cancer type and dataset. The probabilities predicted by LIONHEART were correlated with the probabilities predicted by the other feature sets (Pearson r: 0.53-0.62; Supp. Fig. 3), with mean absolute differences between 0.20 and 0.21 (Supp. Fig. 4). To assess the clinical screening potential, we calculated the sensitivities and specificities per dataset when fixing the other metric at 0.95 and 0.98 (Supp. Fig. 5). At a specificity of 0.95, LIONHEART achieved an average sensitivity of 0.495 (weighted: 0.558). In comparison, the short/long ratios achieved a slightly higher average sensitivity (0.507) but a lower *weighted* average sensitivity (0.516). At a sensitivity of 0.95, LIONHEART achieved a specificity of 0.383. None of the other feature sets came close to this average specificity (Supp. Fig. 5). Further, we computed the precision-recall curves (Supp. Fig. 6A) and calculated the area under the precision-recall curves (AUPRC) (Supp. Fig. 6B). On average, LIONHEART (avg. AUPRC: 0.860, weighted: 0.858) and short/long ratios (avg. AUPRC: 0.862, weighted: 0.845) performed the best. All AUPRC scores were above the prevalence-based baseline (avg. prevalence: 0.569, weighted: 0.500).

**Figure 3:**
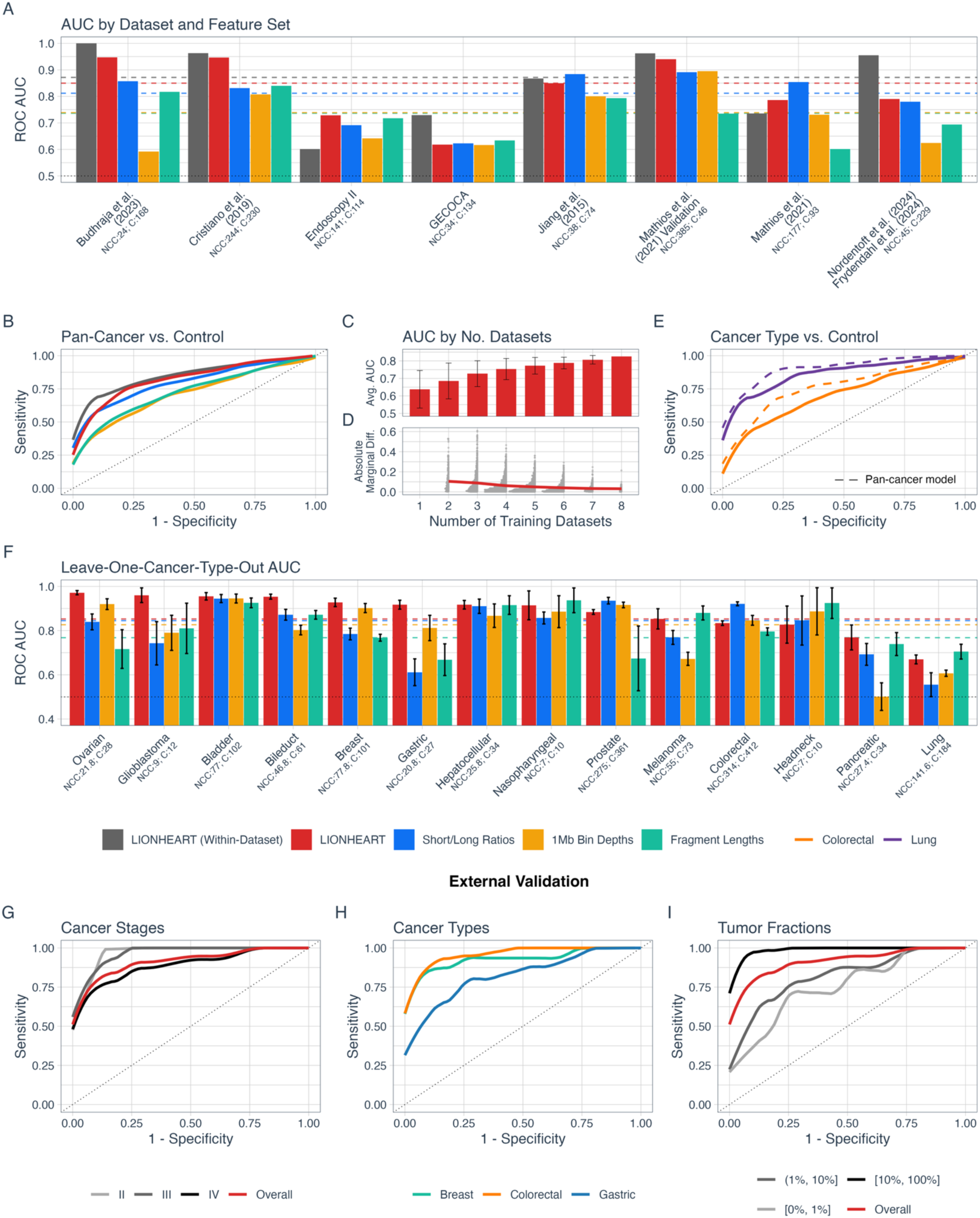
Experimental results. The ROC curves (B, C, D) are calculated per model instance (one per feature set, dataset, and repetition), averaged, and minimally loess smoothed. (…) Dotted lines are chance-level baselines. Counts: **NCC**=Non-Cancer Controls; **C**=Cancer subjects. A) ROC AUC scores per dataset and feature set for classifying pan-cancer vs. non-cancer-control. (---) Dashed lines are the dataset-weighted average AUC scores per feature set. B) ROC curves for pan-cancer vs. non-cancer-control classification, comparing the four feature sets. C) Average ROC AUC scores for leave-one-dataset-out cross-validated LIONHEART models with different numbers of training datasets. Based on all combinations of datasets. More training datasets generally yield higher generalization to unseen datasets. Error bars represent one standard deviation. D) Absolute marginal differences in ROC AUC scores from adding a dataset to the training relative to the same combination of training datasets without that dataset. Whether positive or negative, the impact of adding a dataset reduces the more training datasets that are already present. The training dataset count (x-axis) includes the added dataset. E) ROC curves for single-cancer-type vs. non-cancer-control classification for lung and colorectal cancer. (---) Dashed lines are ROC curves for the pan-cancer model predictions of the same samples. F) ROC AUC scores per cancer type and feature set from leave-one-<u>cancer-type</u>-out nested cross-validation. Each fold is a cancer type with proportionally sampled controls (different sampling per repetition). Error bars represent one standard deviation from the 10 cross-validation repetitions. The results in F do <u>not</u> represent cross-dataset generalization. (---) Dashed lines are the weighted average AUC scores per feature set. G,H,I) ROC curves from external validation on the Zhu et al.^26^ dataset for cancer stage (F), cancer type (G), and Fragle-estimated tumor fractions in bins (H).

Having demonstrated that LIONHEART scores reflect both the origin of the introduced ctDNA fragments and the dilution of the hematopoietic cfDNA, we wanted to ensure that the pan-cancer classifier was not relying solely on the dilution signal, as this might not be cancer-specific. Thus, we tested two variations of the pan-cancer classifier. The first variation included only the 315 cell types from the blood/immune category and achieved an average cross-dataset ROC AUC of 0.788. The second variation instead excluded those same cell types and achieved an average cross-dataset ROC AUC of 0.809. This confirmed that the classifier utilizes both signal types.

To assess the effect of individual datasets on LIONHEART model generalization, we ran leave-one-dataset-out nested cross-validation on all combinations of the datasets and compared the results when including (Supp. Fig. 7) or excluding (Supp. Fig. 8) each dataset from the model training. We first found that the average ROC AUC increased with the number of training datasets (*r* = 0.47, *p* < .001; Fig. 3C). This held true when excluding experiments with training datasets from the same lab/institution as the test dataset (*r* = 0.50, *p* < .001). We then calculated the marginal differences in ROC AUC scores when *adding* a given dataset to the model training, compared to the dataset combination with just that dataset removed (Supp. Fig. 9). We found a negative relationship between the number of already present training datasets and the absolute marginal difference in ROC AUC scores from adding an extra dataset (*r* = -0.27, *p* < .001; Fig. 3D). I.e., the more datasets currently present in the training, the lower the impact of adding an additional dataset. We fit per-dataset one-sample t-tests on the marginal differences and found that seven of the datasets increased the ROC AUC scores when being added to the model training (Δ range = 0.014-0.060, all *p* < .001) while the dataset from Budhraja et al.^23^ had no effect (Δ = -0.001, *p* = .722) and the combined dataset from Nordentoft et al.^24^ and Frydendahl et al.^25^ decreased it (Δ = -0.015, *p* < .001) (Supp. Table 4).

### Pan-cancer classifier vs. cancer-type-specific classifier

We expected the LIONHEART scores to contain both general cancer signals and cancer-type-specific signals. As such, training a classifier only on the samples with a given cancer type and the non-cancer controls could potentially outperform the pan-cancer classifier at detecting that cancer type. On the other hand, the pan-cancer classifier has much more training data available, which is often the bottleneck for model generalization. We compared these two modeling approaches for the two major cancer types that were present in multiple datasets (i.e., colorectal cancer vs. non-cancer-control and lung cancer vs. non-cancer-control) (Fig. 3E). We used leave-one-dataset-out nested cross-validation with the datasets that contained the cancer type in question. The non-cancer controls from the other datasets were included during training. For *colorectal cancer* vs. non-cancer-control, the cancer-type-specific classifier achieved an average ROC AUC score of 0.707 (weighted: 0.717), while the pan-cancer classifier’s predictions for the same samples yielded an average ROC AUC score of 0.764 (weighted: 0.787). For *lung cancer* vs. non-cancer-control, the cancer-type-specific classifier achieved an average ROC AUC score of 0.863 (weighted: 0.853), while the pan-cancer classifier’s predictions for the same samples yielded an average ROC AUC score of 0.906 (weighted: 0.905). The pan-cancer classifier was thus better for both cancer types. Supplementary Table 5 contains the ROC AUC scores per dataset and experiment.

### Generalization to out-of-dataset cancer types

To test whether our pan-cancer classifier is likely to generalize to out-of-dataset cancer types, we merged all datasets and trained pan-cancer vs. non-cancer-control classifiers in nested leave-one-*cancer-type*-out cross-validation, where each cancer type was a fold along with a proportional number of sampled controls (different sampling per cross-validation repetition). ROC AUC scores ranged from 0.667 to 0.972 with an average of 0.882 (weighted: 0.853) (Fig. 3F). On average, LIONHEART generalized better to out-of-sample cancer types than all three benchmarks (Supp. Table 6). This indicates that the method uses a general cancer/non-healthy signal. Unfortunately, we could not measure the cross-dataset generalization of this task, as 10 of the 14 cancer types only appeared in a single dataset.

### External validation of pan-cancer model

After finalizing the method development, we gathered a validation dataset from Zhu et al.^26^, consisting of 96 first-timepoint cancer samples (40 colorectal cancer, 25 gastric cancer, and 31 breast cancer) and 53 non-cancer control samples. We then fit our pan-cancer vs. non-cancer-control LASSO classifier on all nine primary datasets combined to get a general cancer detection model for external use. The model could predict its training data with an average dataset-wise ROC AUC of 0.900. We applied the model to the validation dataset and achieved a ROC AUC of 0.917. We stratified the cancer data on cancer stage (Fig. 3G), cancer type (Fig. 3H), and Fragle^26^-and IchorCNA^27^-estimated tumor fraction (Fig. 3I) (Supp. Fig. 10) and evaluated them against the controls. The model worked for all present cancer stages with ROC AUC scores of 0.969 (II; n=6), 0.967 (III; n=12), and 0.893 (IV; n=68). It detected cancer for all three cancer types with ROC AUC scores of 0.962 (colorectal cancer; n=40), 0.826 (gastric cancer; n=25), and 0.932 (breast cancer; n=31). For intervals of the Fragle-estimated tumor fractions, it achieved ROC AUC scores of 0.755 for fractions below 1% (n=7), 0.821 between 1% and 10% (n=32), and 0.991 for fractions above 10% (n=57). For intervals of the ichorCNA-estimated tumor fractions, it achieved ROC AUC scores of 0.814 for fractions below 3% (n=34), 0.928 between 3% and 10% (n=20), and 0.995 for fractions above 10% (n=42). Supplementary Tables 7-8 contain the stratified results.

To ensure the model could be trusted on external data, we compared the probabilities predicted on the training datasets to those predicted on the external validation dataset (Supp. Fig. 11). The sensitivities and specificities achieved at different probability thresholds on the training data differed only slightly to the actual values on the validation dataset (Supp. Fig. 12). When choosing a threshold for an expected specificity of 0.99, the observed specificity was 1.0 (Supp. Fig. 13A). For an expected sensitivity of 0.99, the observed sensitivity was 1.0 (Supp. Fig. 13B). The threshold yielding the maximum expected sum of sensitivity and specificity (Youden’s J) gave a confusion matrix with 85 (57%) true positives, 41 (27.5%) true negatives, 12 (8.1%) false positives, and 11 (7.4%) false negatives (Supp. Fig. 13C).

Further, we calculated the contributions of each cell type to the classifier and confirmed that cell types in the blood/immune category had a highly positive relationship to the probability of cancer, while the majority of the other categories had a negative relationship to the probability of cancer (Supp. Fig. 14).

## Discussion

Improved early detection of cancer could enhance patient outcomes and reduce healthcare costs. Such a system should preferably work across many cancer types and diverse populations. The fragmentation patterns in WGS cfDNA have been proposed as useful biomarkers for tumor-agnostic cancer detection. Still, most existing fragmentomics methods were validated only within a single dataset or on their generalization to 1-2 external datasets. By optimizing new methods to generalize across cohorts, labs, and relevant demographical variables, we can better isolate the general cancer signal from the noise and confounders, thus increasing the likelihood of successful clinical implementation.

In this study, we developed LIONHEART, a novel fragmentomics method that generalizes across many datasets. The method leverages the genome-wide correlations between cfDNA sequence coverage and the positions of accessible chromatin for 898 cell types (LIONHEART scores). We first showed that the LIONHEART scores generally had the expected relationships with cancer status, validating that higher chromatin accessibility leads to fewer fragments in the WGS cfDNA data. We then combined the LIONHEART scores into our main pan-cancer vs. non-cancer-control classifier and showed high cross-dataset generalization. For two of the eight test datasets, the cross-dataset classifier, trained on the other datasets, predicted the cancer status better than the within-dataset models trained on those datasets specifically. This was likely due to the higher amount of training data available to the cross-dataset models. The LIONHEART pan-cancer classifier performed on par with the fragment length ratio model reported by Cristiano et al.^21^, who reported a within-dataset cross-validation ROC AUC of 0.94 on their dataset, whereas LIONHEART achieved a within-dataset cross-validation ROC AUC of 0.963 and a cross-dataset ROC AUC of 0.947 on the same dataset. On the other hand, Mathios et al.^22^ reported a within-dataset cross-validation ROC AUC of 0.93 on their LUCAS cohort (w/o prior cancer), whereas LIONHEART achieved a within-dataset ROC AUC of 0.735 and a cross-dataset ROC AUC of 0.787. For the GECOCA dataset, neither LIONHEART nor the benchmarks generalized well cross-dataset, although the LIONHEART within-dataset cross-validation ROC AUC was *decent* (0.729). Across the test datasets, LIONHEART performed better on average than all three benchmark feature sets.

Unsurprisingly, we found that adding more datasets to the model training had a positive effect on the generalization to unseen datasets, although the *marginal* effect of adding a dataset decreased with the number of already present datasets. We would expect our current logistic regression model to benefit from more training data, although a more complex, non-linear model architecture might take more advantage of much larger amounts of data. Only the combined dataset from Nordentoft et al.^24^ and Frydendahl et al.^25^ had a significant *negative* marginal effect on the generalization. This dataset had a much higher sequencing depth than most of the other datasets, with many samples having a very low ctDNA fraction (median ctDNA fraction in the colorectal cancer samples^25^: 1.7 ×10^-3^). Such low ctDNA fractions may have caused the model to fit to non-cancerous differences (random or batch effects), if little cancer signal was present in the LIONHEART scores.

We showed that the pan-cancer classifier uses both the cancer and blood-dilution signals in the LIONHEART scores. This was supported by the classifier’s generalization to out-of-data cancer types, although we could not measure this generalization cross-dataset due to the limited data available for the individual cancer types. The pan-cancer classifier further outperformed cancer-type-specific classifiers in detecting lung and colorectal cancer, likely due to the higher availability of training data.

Finally, we validated the LIONHEART classifier on an external dataset from Zhu et al.^26^. It detected cancer in all cancer stages and all three cancer types with ROC AUC scores between 0.826 and 0.969, although it did have problems on samples with Fragle-estimated tumor fractions below 1%. While we directly optimized our method towards high cross-dataset generalization, there was still a risk of overfitting to the utilized datasets, requiring external validation of the final method. The trained model and code to extract LIONHEART scores are publicly available via GitHub, enabling researchers without large training datasets to use LIONHEART on their data. Several previous articles have utilized nucleosome positioning features for cancer detection or subtyping^13–16^. However, the present study is the first to comprehensively test the cross-dataset applicability of such a method. The demonstrated cross-dataset applicability of LIONHEART means that we expect it to work on external datasets without retraining, assuming a similar age range and sequencing depth (Supp. Fig. 15).

A key bottleneck to fragmentomics-based cancer detection methods is the scarcity of available data for many cancer types, given the high heterogeneity of cancer cases. Even with nine datasets, most of the present cancer types were sparsely represented, while most known cancer types were not even included. To generalize to most individuals in a population, the data must adequately represent both the healthy states and the relevant cancer signals in that population. This includes discerning cancer from other cfDNA-affecting states like pregnancy^28^ and autoimmune disease^29^. For samples with low tumor fraction, it is, for instance, possible that the model relies on immune response signals rather than direct signals of ctDNA presence^30,31^. Such a cancer-immune response signal could be similar to that of, e.g., non-cancerous inflammatory conditions^32,33^. With enough data, the model could either learn to discern these states, or we could create separate models for high and low tumor fractions to increase model interpretation and patient stratification.

Whereas privacy concerns often make sharing genomic data cumbersome, the summary-statistic nature of LIONHEART features could make it easier in some countries. This could allow the collection and sharing of additional data, enabling better and more complex models.

In conclusion, we used the correlations between WGS cfDNA sequencing coverage and accessible chromatin positions of different cell types to detect cancer. We then evaluated the cross-dataset generalizability of the method to strengthen our confidence in its ability to detect general cancer signals. With cfDNA-based methods getting closer to clinical utilization (e.g., via the Galleri test from Grail and the FirstLook test from DELFI Diagnostics), we propose integrating cross-cohort and cross-lab generalization measures as key optimization metrics in the field to ensure robustness and reliability across a multitude of settings and to facilitate clinical implementation.

## Methods

### cfDNA data

We employed ten datasets (nine for the primary analysis and one for validation) with mostly low-coverage whole genome sequenced blood plasma cell-free DNA from ^21–26,34,35^ and some in-house datasets (Supp. Table 1). The datasets contained 14 cancer types (colorectal, lung, breast, prostate, bile duct, ovarian, gastric, bladder, pancreatic, glioblastoma, melanoma, hepatocellular carcinoma, head and neck squamous cell carcinoma, and nasopharyngeal carcinoma) with various histologic subtypes (Supp. Fig. 16). All the datasets contained non-cancer controls. Sequencing depths in the nine primary datasets varied from ∼0.02x to ∼57x, with 63.3% of samples being ∼1-3x, while all samples in the external validation dataset had a depth of ∼1.25 (Supp. Table 9). Only one sample was included per subject. All datasets were (re)mapped to hg38. Supplementary Figure 15 shows the age, sex, and sampling depth distributions per primary dataset and group.

#### Endoscopy II: Symptomatic colorectal cancer WGS cohort

Plasma samples were obtained from the previously reported Danish Endoscopy II study^36,37^. The Endoscopy II protocol was initiated in 2010 and terminated in 2012 after having included 4698 individuals referred to diagnostic colonoscopy due to symptoms attributable to colorectal cancer. Of the recruited individuals, 512 were diagnosed with colorectal cancer, 689 with colorectal adenoma, and 3497 did not have colorectal neoplasia (non-cancer controls). Written informed consent was obtained from all participants, and the study was performed according to the Declaration of Helsinki. The Endoscopy II study was approved by the Scientific Ethics Committees for the Capital Region of Denmark (j. no H-3-2009-110) and by the GDPR-compliant Danish Data Protection Agency (2008-41-2252). Here, we included plasma from 114 colorectal cancer patients and 141 non-cancer controls. The present analysis was approved by the Danish National Committee on Health Research Ethics (j. no. 2001584).

In brief, EDTA blood was collected before colonoscopy. Within two hours of the blood draw, the samples were centrifuged at 3,000g for 10 min at 21°C, and plasma was separated from the red blood cells and buffy-coat layer and stored at -80°C. cfDNA was isolated from ∼4 ml of plasma using the Qiagen QIAamp Circulating Nucleic Acids Kit (Qiagen GmbH). Extracted cfDNA was eluted in 52ul into LoBind tubes (Eppendorf AG) and quantified using the Bioanalyzer 2100 (Agilent Technologies).

Genomic library construction and sequencing: As previously described, cfDNA libraries for next-generation whole-genome sequencing were prepared with 15 ng of cfDNA when available or the entire purified amount when less than 15 ng^21,22^. The genomic libraries were prepared using the NEBNext DNA Library Prep Kit for Illumina (New England Biolab) with four main modifications to the manufacturer’s guidelines: (i) the library purification steps followed the on-bead AMPure XP (Beckman Coulter) approach to minimize sample loss during elution and tube transfer steps; (ii) NEBNext End Repair, A-tailing, and adapter ligation enzyme and buffer volumes were adjusted as appropriate to accommodate on-bead AMPure XP purification; (iii) Illumina dual index adapters were used in the ligation reaction; and (iv) cfDNA libraries were amplified with Phusion Hot Start Polymerase. All samples underwent a 4-cycle PCR amplification after the DNA ligation step. Genomic sequencing was performed in batches that included samples from individuals with or without cancer to reduce the possibility that differences between patients with or without cancer were due to batch variability. Whole-genome libraries were sequenced using 100-bp paired-end runs (200 cycles) on the Illumina HiSeq2500 platform. The median sequencing coverage was 1.17× (IQR: 0.31).

#### GECOCA WGS cohort

We selected 134 EDTA blood samples from treatment-naïve colorectal cancer patients (UICC stages I-IV) from the GECOCA cohort and 34 EDTA blood samples from non-cancer, healthy controls obtained from the Blood Bank at Aarhus University Hospital, Denmark. Informed consent was obtained from all subjects involved, and the GECOCA study was approved by the Danish National Committee on Health Research Ethics (j. no. 2208092). Processing of blood to plasma and extraction of cfDNA from plasma was done as described above for the Endoscopy II cohort, except that 8 mL plasma aliquots were used. cfDNA was ligated to xGen UDI-UMI Adapters (Integrated DNA Technologies Inc., IDT, Coralville, IA, USA) using the KAPA HyperPrep library kit (Roche, Basel, Switzerland). All cfDNA extracted from 8 mL plasma was used as input except for samples with more than 80 ng total cfDNA, in which case input was equally split into two libraries (for a total of up to 160 ng cfDNA per sample).

Post-ligation clean-up was done using AMPURE beads in a 1.4× beads/DNA ratio to retain short fragments. After ligation, libraries were split into four separate reactions and amplified for seven cycles of PCR before finally being purified using a 1.0× beads/DNA ratio and quantified using the Qubit dsDNA BR Assay Kit (ThermoFisher). The libraries were subjected to 2 x 151 bp paired-end whole genome sequencing on an Illumina NovaSeq v1.5. The median sequencing coverage was 2.23× (IQR: 0.33).

#### Prostate Cancer, Aarhus cohorts

This dataset included previously published plasma low-coverage WGS data from 143 prostate patients (described in detail in ^35^) as well as newly generated plasma low-coverage WGS data from another set of 218 prostate cancer patients and 18 male non-cancer controls (cancer-negative prostate biopsy). All blood samples were collected at Aarhus University Hospital, Denmark, between 2006 and 2023. Within 2 hours of the blood draw, plasma was separated from cellular components by centrifugation (3000g for 10 minutes at 20°C) and stored in cryo tubes (TPP) at -80°C. cfDNA was purified from 4 mL plasma and quantified as described previously^35^. The prostate cancer study was carried out in accordance with local guidelines and regulations and approved by either the Central Denmark Region Committees on Health Research Ethics (no. 1-10-72-139-15, no. 1-10-72-262-21, no. 1-10-72-367-13, and no. 1-10-72-337-16) or The National Committee on Health Research Ethics (no. 1302791 and no. 1901101) and notified to the Danish Data Protection Agency (no. 1–16–02-330-13, no. 1–16–02-366-15, no. 1-16-02-388-21, no.1-16-02-248-14, and no. 1-16-02-686-16). The study adhered to the principles of the Declaration of Helsinki. All patients provided written informed consent for biobanking. The requirement for patient consent to the specific analyses in this study was waived. Libraries for low-coverage WGS were prepared from up to 100 ng cfDNA (3.6ng-100ng) using the KAPA HyperPrep kit (KAPA Biosystems) according to the manufacturer’s instructions, except for only using half volumes of all reagents. After ligation of xGen CS-adapters – Tech Access (IDT-DNA) and clean-up using 1.4x AMPure XP Beads (Beckman Coulter), the total amount of input DNA in each sample was equalized to 100 ng by the addition of lambda filler DNA, containing a 1:1 ratio mixture of unmethylated and in vitro methylated lambda phage DNA amplicons, according to the protocol described in Sheen et al.^38^. Thus, a 7.9 µL aliquot of each library was purified and eluted in 50 µL buffer C using the IPure kit v2 (Diagenode). Each library sample was amplified using the KAPA HiFi Hotstart ReadyMix (Roche) and uniquely indexed primer pairs (IDT-DNA). Indexed libraries were purified using 1.0x AMPure XP Beads (Beckman Coulter), quantified using the Qubit dsDNA High Sensitivity assay (ThermoFischer Scientifics), and paired-end sequenced (2×151bp) on an Illumina® Novaseq 6000 instrument. The median sequencing coverage was 0.47× (IQR: 0.14).

### Accessible chromatin locations

We downloaded DNase-seq narrow peak intervals for 1363 samples from the ENCODE project^39,40^, representing 353 cell-and tissue types (referred to as ‘cell types’). We further downloaded ATAC-seq narrow peak intervals for 358 samples from the ENCODE project, 1493 samples from ATACdb^41^, and 23 samples from Corces et al.^42^, together representing 543 cell types. The samples from ATACdb were converted to hg38 using LiftOver^43^. Besides tissue-of-origin, we considered cell types as separate based on their biosample type (primary cell, cell line, in vitro differentiated cell, induced pluripotent stem cell line, tissue, stem cell, and “other”), health status (derived from asthma, branchio-oculo-facial syndrome, caid syndrome, cancer, cardiomyopathy, cirrhosis, coronary artery disease, diabetes, emphysema, Epstein-Barr virus, frontotemporal degeneration, HIV-1, hypertension, hypogammaglobulinemia, hemoglobinopathies, sclerosis, ischemia, systemic lupus erythematosus, yellow fever, and other/healthy), and whether the sample was derived from an adult or an embryo. We treated the DNase-seq and ATAC-seq sets separately throughout the data processing until the machine learning pipeline. Combined, we ended up with 896 (700 unique) cell types (Supp. Table 10). All samples from a given cell type were merged such that positions considered part of a peak in at least 30% of the samples were kept, with the rest being discarded as potential noise. The *raw* ATAC-seq files had an average of 61,442.5 (std: 64,949.8) peaks with an average length of 625.5bp (pooled std: 985.6bp, avg. median: 442.9bp, min: 87bp, max: 2,451,863bp), while the raw DNase-seq files had an average of 104,853.3 (std: 83,984.7) peaks with an average length of 194.3bp (pooled std: 103.2bp, avg. median: 184.3bp, min: 1bp, max: 30,796bp).

#### Processing of position masks

We split the hg38 autosomes into *263,051,621* bins, each 10bp in size, excluding non-mappable regions from the ENCODE blacklist v2^44^ and the umap k100 exclusion file^45^. For each bin, we extracted the GC fractions in a 100bp context for later use in bias correction. For each cell type and bin, we extracted the percentage of positions that overlapped an open chromatin site in that cell type.

### cfDNA sample processing

For each 10bp bin, we used a modified version of mosdepth^46^ to extract the average position coverage (here from “coverage”) and the average position-overlapping insert size (fragment length). Only fragments of size 100-220bp were included as the datasets seemed most consistent in that range (Supp. Fig. 17). These values were saved chromosome-wise as sparse arrays using SciPy^47^. Supplementary Table 11 visualizes the relationship between variables used in the cfDNA processing, using hypothetical data per 10bp bin.

#### Outlier removal

Despite excluding fragments with low mappability, we found extreme outliers among the bin coverages. We fitted chromosome-wise zero-inflated Poisson distributions to the coverages and removed 422,587 bins (∼0.16%) that had a probability p(C ≥ c) < 1e-4 (i.e., complementary cumulative distribution function) in 25% of samples in any of the nine datasets. We further truncated sample-specific extreme coverages above the average coverage with a probability p(C ≥ c) < 1/num_bins (where num_bins = 263,051,621) to the first discrete coverage value with a probability above this threshold. This approach generalizes to out-of-sample data. 24,776 bins (0.009%) with zero coverage across all datasets were also removed.

#### Bias correction

To increase generalization across datasets, we sequentially corrected the GC bias and homogenized the average overlapping insert size bias, sample- and chromosome-wise.

#### GC-bias correction

We split the distribution of GC fractions in the reference genome into 43 GC fraction bins by first binning the ordered fractions into 100 bins of equal mass and then removing duplicate bin edges. We calculated the average bin coverage within each GC fraction bin, smoothed them with a Gaussian kernel, and scaled them to be 1-centered (hereafter referred to as correction factors). We corrected the coverages by dividing each coverage value by the correction factor for its GC fraction bin, flattening the GC bias. To avoid extreme corrections of 10bp bins with extreme GC fractions, we excluded all 10bp bins with GC fractions below 20% or above 67%, together representing 2/100^ths^ of the 10bp bins. The GC bias differed within and between datasets (Supp. Fig. 18).

#### Average overlapping insert size transformation

We extracted the average overlapping insert size for each 10bp bin using a modified version of *mosdepth* and calculated the average coverage per 3bp interval of insert sizes. These distributions varied by sample and dataset regarding mean, spread, skewness, and “noise”. To increase the similarity of the datasets, we removed the noise and skewness and shifted the mean to 166bp for all samples (see Supp. Note 1 for a more detailed explanation of these corrections).

The distribution spread was assumed to be a product of the central limit theorem, with the *10bp bin-wise* spread of overlapping insert sizes being inversely proportional to the square root of the coverage. I.e., the central limit theorem affected each 10bp bin separately. We captured this dynamic with a mixture of skewed Student’s t distributions, truncated to the possible insert sizes (100-220bp). The mixture had a distribution per unique coverage value (rounded to whole numbers) with an appropriately scaled spread. The distributions used the same optimized mean and skewness parameter values and were combined as a sum, weighted by the frequency of the coverage values.

We performed three sequential transformations for each sample and chromosome. First, we fitted the mixture distribution as described. A correction factor was calculated as the difference between the observed and fitted distributions to remove the “noise”. A second correction factor removed the skewness. Since removing the skewness altered the mean value, we refitted the mixture distribution and calculated a third correction factor as the relative difference between this fitted distribution and the same distribution with its mean shifted to 166bp. After applying this correction, all samples had the same mean, no noise, and no skewness (Supp. Fig. 19; Supp. Note 1). Finally, the few 10bp bins with average overlapping insert sizes below 113bp or above 217bp were excluded to avoid extreme corrections.

#### Normalization of copy number alterations

To reduce the potential effect of copy number alterations, we calculated the average coverage in 5Mb bins across the genome with a 500kb stride and divided each coverage value by the average overlapping 5Mb bin average coverage (Supp. Fig. 20). This averaging approach weights the final average coverage scaling factor by the proximity to the 10bp bin.

### Features

#### LIONHEART scores

We calculated Pearson R correlations between the bias-corrected coverages and the cell type-specific open chromatin site overlap fractions. This gave us 896 correlation coefficients per cfDNA sample, each describing the presence of a cell type. Consensus bins, where 90% or more of the cell types were indicated as open, were excluded from the cell type correlations and added as separate correlation features (one for ATAC-seq and one for DNase-seq). We standardized the features sample-wise to reduce the effects of sampling depth. For each dataset, we thus had a 2-dimensional NumPy array with shape (# samples, 898). Supplementary Figure 21 shows a heatmap of the LIONHEART scores sorted by dataset and cancer status, while Supplementary Figure 22 shows the average standardized LIONHEART scores.

#### Benchmark features

For benchmarking, we extracted three common fragmentomics feature sets for each sample: a) the fragment length distribution from 80-400bp, b) the ratio of short (100-150bp) to long (151-220bp) fragment counts in 1Mb bins, and c) the coverage depth in 1Mb bins normalized to have a sample mean of 1. We used ctDNAtool (https://github.com/BesenbacherLab/ctDNAtool) to extract the length distribution in bins and used these to calculate the benchmark features. No corrections were applied to the benchmark features.

### Machine Learning

We fitted LASSO-regularized logistic regression models in leave-one-*dataset*-out nested cross-validation (except when specified otherwise). The inner loop selected the set of hyperparameter values (e.g., the LASSO regularization strength) that achieved the highest balanced accuracy in a grid search, using either leave-one-*dataset*-out cross-validation (when possible) or classic 10-repetition 5-fold cross-validation. The outer loop estimated the out-of-sample dataset-wise ROC AUC. The training loss was weighted by the target class (first within each dataset, then overall in the training data). Predictions from the outer loop were evaluated with ROC AUC. **Preprocessing**: Based on parameters learned from the training data, the LIONHEART scores were feature-standardized, while the benchmark features were feature-centered. We performed principal component analysis and selected the first principal components that explained between 98.7% and 99.7% of the variance (hyperparameter). Finally, the PCA-transformed features were standardized and fed to the model. **Testing**: The *test* samples were treated independently to ensure the model generalized to single samples. **Validation**: We fitted a model on all the data at once and used it to predict the cancer status of the samples in the validation dataset. The probability thresholds for achieving the maximum Youden’s J and certain levels of sensitivity and specificity were calculated from the model’s predictions on its training data. We evaluated the predictions at different tumor fraction intervals estimated with Fragle^26^ and ichorCNA^27^. The model is available in the public repository. Supplementary Figure 23 shows the pre-processed features selected by the validated LASSO model, sorted by the absolute model coefficient.

We used the LASSO logistic regression implementation and a modified cross-validation function from scikit-learn^48^. Depending on the analysis, evaluation metrics were calculated with either scikit-learn^48^, the cvms R package^49^, or manually in the *generalize* python package (see Code availability).

## Code availability

The code for extracting LIONHEART scores and predicting cancer status is available as a command-line tool on GitHub ( https://github.com/BesenbacherLab/lionheart). Resources and trained models are available via Zenodo (link on GitHub). Note that the models were trained on data that can only be used for non-commercial research purposes. The code for running leave-one-dataset-out nested cross-validation and univariate modeling is available in the “*generalize*” Python package ( https://github.com/LudvigOlsen/generalize).

## Data availability

The calculated LIONHEART scores are available on Zenodo (https://zenodo.org/records/15747531), allowing the model to be updated with local data. These summary statistics were calculated from data that can only be used for non-commercial research purposes.

The low-coverage WGS data generated for this article (*GECOCA*, *Endoscopy II*, and the *Prostate Cancer, Aarhus Cohorts*) are available through controlled access from GenomeDK (https://genome.au.dk/library) (GDK000013, GDK000014, GDK000015).

External WGS datasets are available at EGA (EGAD00001007796, EGAD00001005339, EGAD50000000167, EGAD00001005093), dbGaP (phs003170.v1.p1), and GenomeDK (GDK000009). The dataset from Nordentoft et al.^24^ is not publicly available.

ATAC-seq and DNase-seq data are available from ENCODE (https://www.encodeproject.org/), ATACdb (http://www.licpathway.net/ATACdb), and TCGA (https://gdc.cancer.gov/about-data/publications/ATACseq-AWG). See Supplementary Table 10 for the sample IDs of the used files.

## Acknowledgements

We thank and acknowledge the Danish Cancer Biobank, the Endoscopy II study teams at the university hospitals in Aarhus, Hvidovre, and Bispebjerg, and the Regional hospitals in Herning, Hillerød, Horsens, and Randers for providing access to blood and tissue materials.

This work was supported by Aarhus University Research Foundation (grant no. AUFF-F-2020-7-8 to S.B.), the Novo Nordisk Foundation (grant no. NNF22OC0074415 to C.L.A.), the Danish Cancer Society (grant numbers R146-A9466-16-S2 (C.L.A.), R231-A13845 (C.L.A.), and R257-A14700 (C.L.A.)). Delfi Diagnostics Inc. covered the cost of sequencing the Endoscopy II symptomatic colorectal cancer cohort.

The Aarhus prostate cancer studies were supported by grants from the Novo Nordisk Foundation (no. NNF20OC0059410 to K.D.S) and the Danish Cancer Society (no. R281-A16122 to K.D.S. and no. R306-A18131 to M.R.).

## Contributions

L.R.O., J.Q.H., A.J.S., and S.B. designed the study. L.R.O. implemented the software and ran computational experiments. D.O. ran feature extraction for external validation. S.Bu., M.B., M.R.J., K.B., and C.T. provided patient samples or clinical data. K.K., L.I., M.R., B.E.L., A.F., M.H.R., T.V.H, M.N., C.D., S.V.L., I.N., P.L., L.D., K.D.S., C.L.A., and A.J.S. organized sample data and generated or oversaw the generation of sequencing data. L.R.O. and S.B. wrote the article with large contributions from C.L.A., M.H.R., and K.K. All authors read, commented, and approved the final manuscript.

