## Supplementary Material for "Cross-dataset pan-cancer detection: Correlating cell-free DNA fragment coverage with open chromatin sites across cell types"

#### Results

| Dataset | Controls | Cancers | Cancer Types | Notes |
|---|---|---|---|---|
| **Main analysis** | | | | |
| Budhraja et al., 2023 | 24 | 168 | Melanoma (73), Breast (47), Bile Duct (36), Glioblastoma (12) | |
| Nordentoft et al., 2024<br>Frydendahl et al., 2024 | 45 | 229 | Colorectal (127), Bladder (102) | These datasets are from the same lab and were merged.<br>High sequencing depth: ~11-57x. |
| GECOCA | 34 | 134 | Colorectal (134) | Includes 10 high-coverage cancer samples. |
| Endoscopy II | 141 | 114 | Colon (70), Rectal (44) | Symptomatic colorectal cancer cohort. |
| Jiang et al., 2015 | 38 | 74 | Hepatocellular Carcinoma (34), Colorectal (10), Lung (10), Head and Neck Squamous Cell Carcinoma (10), Nasopharyngeal Carcinoma (10) | |
| Cristiano et al., 2019 | 244 | 230 | Breast (54), Lung (35), Pancreatic (34), Ovarian (28), Colorectal (27), Gastric (27), Bile Duct (25) | |
| Mathios et al., 2021 | 177 | 93 | Lung (88), Met. to lung (5) | W/o prior cancer. Excluded 6 control samples with cancer diagnosis within a year of blood draw and 1 missing sample. |
| Mathios et al., 2021<br>Validation Cohort | 385 | 46 | Lung (46) | |
| Prostate Cancer, Aarhus cohorts<br>Nørgaard et al., 2023 and unpublished | 18 | 361 | Prostate (361) | Mix of published and unpublished in-house cohorts. Mainly used for training and feature development. Contains too few control samples to properly test cancer vs. control classifiers on. |
| Combined | 1106 | 1449 | Colorectal (412), Lung (184), … | |
| **Validation** | | | | |
| Zhu et al., 2023 | 53 | 96 | Colorectal (40), Breast (31), Gastric (25) | Only includes the pre-treatment samples. Excludes duplicates. |

**Supplementary Table 1**: Number of samples per dataset, split by cancer status. The *GECOCA*, *Endoscopy II*, and parts of *Prostate Cancer, Aarhus Cohorts* datasets are presented as part of this article.

### Univariate modeling

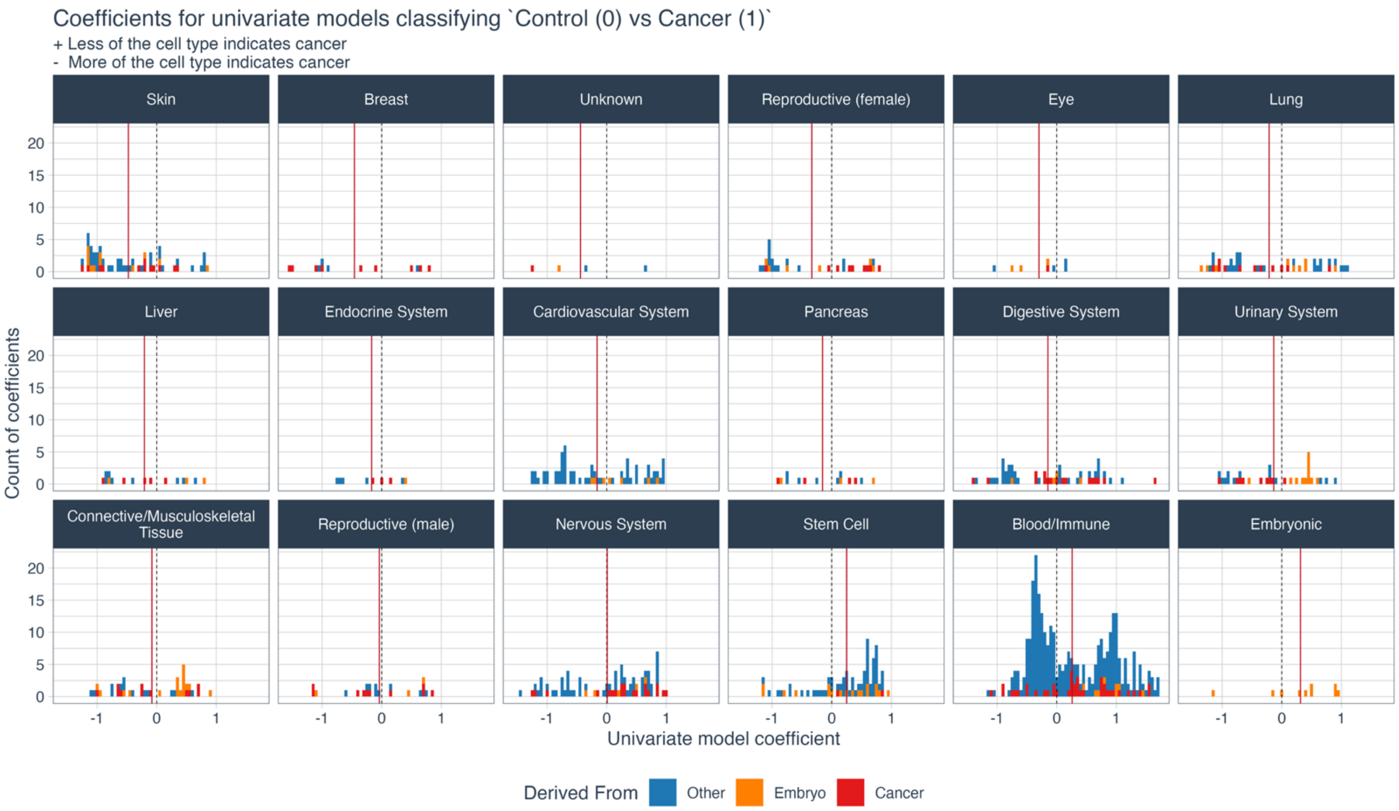


**Supplementary Figure 1:** Histograms of univariate modeling coefficients (one per cell type feature) in categories. Each data point in the histogram is a coefficient for a single cell type feature. Color: Whether the cell is derived from cancer, an embryo, or neither. Dashed grey line: The zero-coefficient (i.e., x-axis is 0). Red vertical line: The average coefficient in the category.

#### Pan-cancer vs. non-cancer controls

| Test Dataset | Cross-dataset ROC AUC | | | | Number of samples | |
|---|---|---|---|---|---|---|
| | LIONHEART | Depth in 1Mb bins | Short/Long Ratios in 1Mb bins | Fragment Length Distribution | N - | N + |
| Budhraja et al., 2023 | 0.947 | 0.592 | 0.857 | 0.816 | 24 | 168 |
| Cristiano et al., 2019 | 0.947 | 0.807 | 0.831 | 0.840 | 244 | 230 |
| Endoscopy II | 0.728 | 0.642 | 0.691 | 0.718 | 141 | 114 |
| Mathios et al., 2021 | 0.787 | 0.731 | 0.854 | 0.602 | 177 | 93 |
| Mathios et al., 2021 Validation Cohort | 0.940 | 0.895 | 0.891 | 0.735 | 385 | 46 |
| Nordentoft et al., 2024; Frydendahl et al., 2024 | 0.790 | 0.624 | 0.780 | 0.693 | 45 | 229 |
| Jiang et al, 2015 | 0.850 | 0.800 | 0.884 | 0.793 | 38 | 74 |
| GECOCA | 0.618 | 0.616 | 0.623 | 0.634 | 34 | 134 |
| **Average** | **0.826** | 0.713 | 0.801 | 0.729 | | |
| **Weighted Average** | **0.850** | 0.739 | 0.812 | 0.736 | | |
| | Within-dataset ROC AUC | | | | | |
| | LIONHEART | Depth in 1Mb bins | Short/Long Ratios in 1Mb bins | Fragment Length Distribution | N - | N + |
| Budhraja et al., 2023 | 1.00 | 0.992 | 0.840 | 0.891 | 24 | 168 |
| Cristiano et al., 2019 | 0.963 | 0.931 | 0.920 | 0.933 | 244 | 230 |
| Endoscopy II | 0.602 | 0.574 | 0.665 | 0.659 | 141 | 114 |
| Mathios et al., 2021 | 0.735 | 0.806 | 0.733 | 0.720 | 177 | 93 |
| Mathios et al., 2021 Validation Cohort | 0.962 | 0.982 | 0.924 | 0.951 | 385 | 46 |
| Nordentoft et al., 2024; Frydendahl et al., 2024 | 0.955 | 0.883 | 0.813 | 0.681 | 45 | 229 |
| Jiang et al, 2015 | 0.867 | 0.775 | 0.691 | 0.767 | 38 | 74 |
| GECOCA | 0.729 | 0.620 | 0.588 | 0.642 | 34 | 134 |
| **Average** | **0.852** | 0.820 | 0.7717 | 0.780 | | |
| **Weighted Average** | **0.872** | 0.851 | 0.810 | 0.812 | | |

**Supplementary Table 2**: ROC AUC scores from pan-cancer vs. non-cancer-control binary classification experiments. **Cross-dataset**: From leave-one-*dataset*-out nested cross-validation. Each score is the test score when trained on all other datasets. The Prostate Cancer (Aarhus cohorts) dataset was only used for training due to the low number of controls (n=18). **Within-dataset**: Per-dataset regular nested cross-validation results for comparison. **N-**: Number of control subjects; **N+**: Number of cancer subjects.

| Cancer Type | Test Dataset | Cross-dataset ROC AUC | | | | Number of samples | |
|---|---|---|---|---|---|---|---|
| | | LIONHEART | Depth in 1Mb bins | Short/Long Ratios in 1Mb bins | Fragment Length Distribution | N - | N + |
| **Colorectal** | | | | | | | |
| | GECOCA | 0.618 | 0.616 | 0.623 | **0.634** | 34 | 134 |
| | Nordentoft et al., 2024; Frydendahl et al., 2024 | 0.781 | 0.586 | **0.815** | 0.720 | 45 | 127 |
| | Endoscopy II | **0.728** | 0.642 | 0.691 | 0.718 | 141 | 114 |
| | Cristiano et al., 2019 | **0.961** | 0.772 | 0.896 | 0.913 | 244 | 27 |
| | Jiang et al., 2015 | 0.729 | 0.597 | **0.926** | 0.811 | 38 | 10 |
| | **Average \| Total** | 0.764 | 0.643 | **0.790** | 0.759 | 502 | 412 |
| | **Weighted Average** | **0.787** | 0.663 | 0.775 | 0.766 | | |
| **Lung** | | | | | | | |
| | Mathios et al., 2021 | 0.787 | 0.731 | **0.854** | 0.602 | 177 | 93 |
| | Mathios et al., 2021 Validation Cohort | **0.940** | 0.895 | 0.891 | 0.735 | 385 | 46 |
| | Cristiano et al., 2019 | 0.959 | 0.915 | **0.972** | 0.917 | 244 | 35 |
| | Jiang et al., 2015 | 0.939 | **0.989** | 0.966 | 0.892 | 38 | 10 |
| | **Average \| Total** | 0.906 | 0.883 | **0.921** | 0.786 | 844 | 184 |
| | **Weighted Average** | 0.905 | 0.862 | **0.907** | 0.757 | | |
| **Bladder** | | | | | | | |
| | Nordentoft et al., 2024; Frydendahl et al., 2024 | **0.801** | 0.672 | 0.735 | 0.660 | 45 | 102 |
| **Breast** | | | | | | | |
| | Budhraja et al., 2023 | **0.932** | 0.527 | 0.804 | 0.746 | 24 | 47 |
| | Cristiano et al., 2019 | **0.966** | 0.917 | 0.781 | 0.791 | 244 | 54 |
| **Hepatocellular Carcinoma** | | | | | | | |
| | Jiang et al., 2015 | 0.875 | 0.840 | **0.926** | 0.762 | 38 | 34 |
| **Pancreatic** | | | | | | | |
| | Cristiano et al., 2019 | 0.858 | 0.535 | 0.789 | **0.891** | 244 | 34 |
| **Ovarian** | | | | | | | |
| | Cristiano et al., 2019 | **0.993** | 0.961 | 0.900 | 0.855 | 244 | 28 |
| **Gastric** | | | | | | | |
| | Cristiano et al., 2019 | **0.953** | 0.874 | 0.624 | 0.664 | 244 | 27 |
| **Bile Duct** | | | | | | | |
| | Budhraja et al., 2023 | 0.991 | 0.818 | **0.993** | 0.846 | 24 | 36 |
| | Cristiano et al., 2019 | **0.933** | 0.582 | 0.831 | 0.904 | 244 | 25 |
| **Melanoma** | | | | | | | |
| | Budhraja et al., 2023 | **0.929** | 0.516 | 0.852 | 0.820 | 24 | 73 |
| **Glioblastoma** | | | | | | | |
| | Budhraja et al., 2023 | **0.986** | 0.628 | 0.889 | 0.799 | 24 | 12 |
| **Head and Neck Squamous Cell Carcinoma** | | | | | | | |
| | Jiang et al., 2015 | 0.818 | **0.824** | 0.721 | 0.779 | 38 | 10 |
| **Nasopharyngeal Carcinoma** | | | | | | | |
| | Jiang et al., 2015 | **0.824** | 0.658 | 0.779 | 0.797 | 38 | 10 |

**Supplementary Table 3**: Cross-dataset ROC AUC scores per cancer type and dataset based on the pan-cancer vs. non-cancer-control model predictions from leave-one-dataset-out nested cross-validation. The model predictions for a given cancer type were evaluated against the controls in the same dataset.

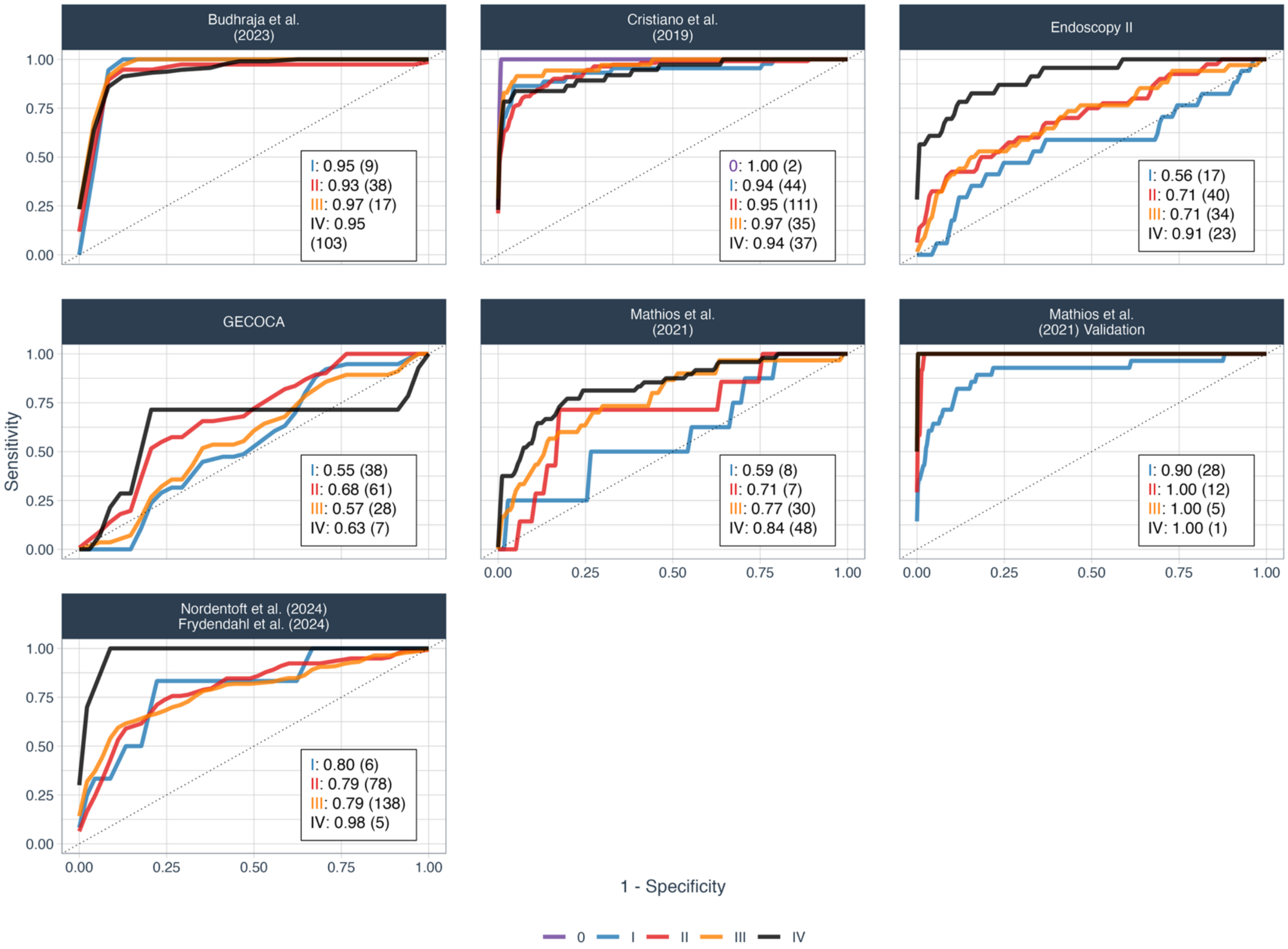


**Supplementary Figure 2**: ROC curves per dataset and cancer stage from leave-one-dataset-out cross-validation of the LIONHEART pan-cancer vs. non-cancer control classifier. Annotated with the ROC AUC score and sample size per stage. Staging information was missing for one gastric cancer sample from Cristiano et al., two bladder cancer samples from Nordentoft et al., and all samples from Jiang et al.

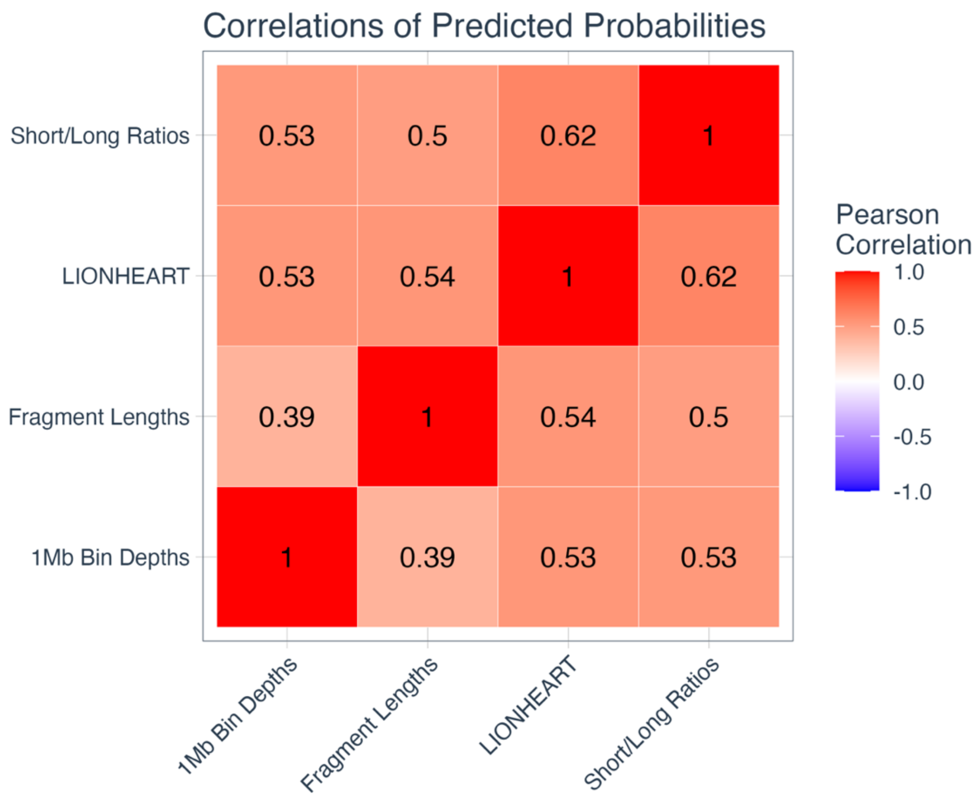


**Supplementary Figure 3**: Pairwise Pearson correlations between the sets of predicted probabilities by models trained on each feature set.

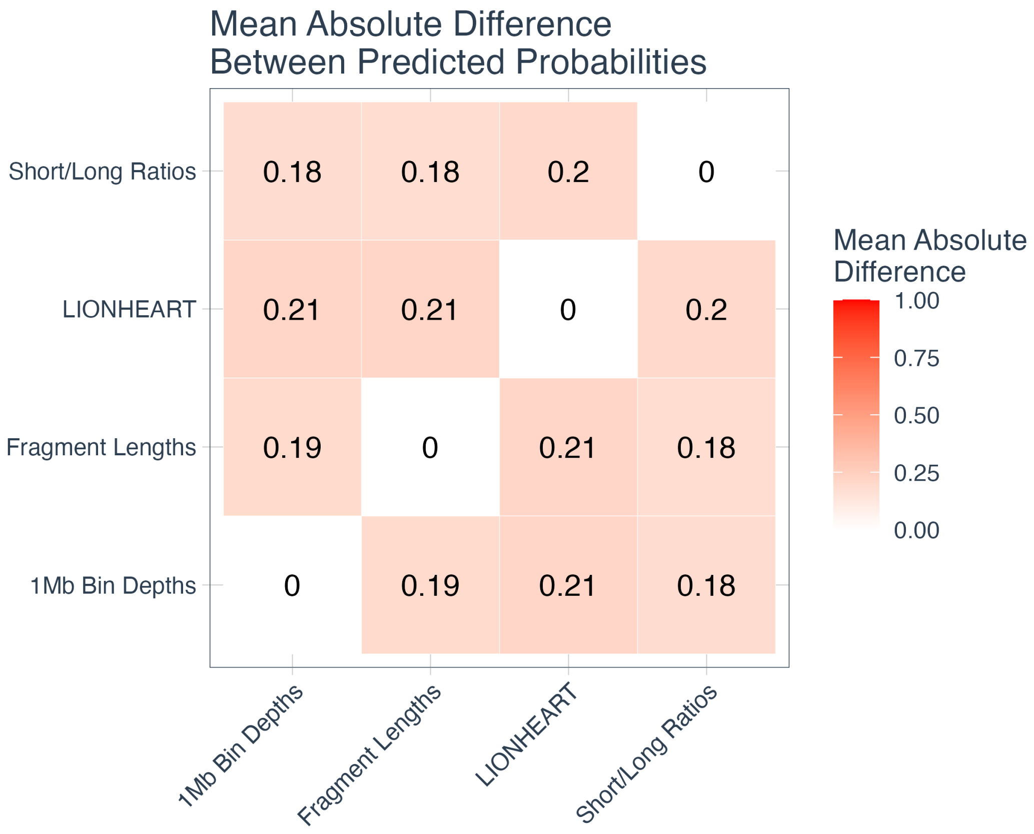


**Supplementary Figure 4**: Pairwise mean absolute differences between the sets of predicted probabilities by models trained on each feature set.

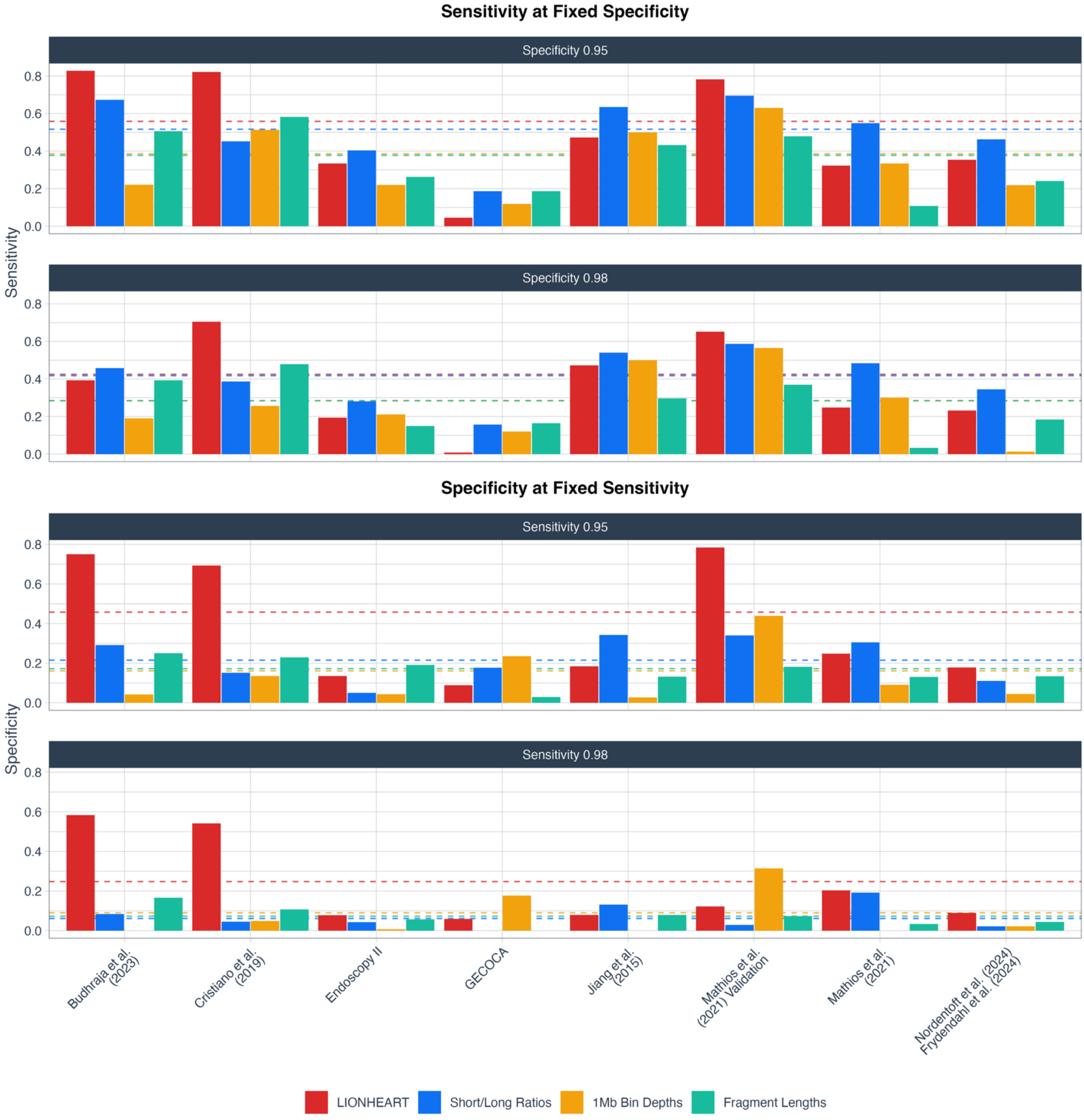


**Supplementary Figure 5**: Sensitivities and specificities per dataset and feature set at fixed points. From leave-one-dataset-out cross-validation. Top: Sensitivities at fixed specificities of 0.95 and 0.98. Bottom: Specificities at fixed sensitivities of 0.95 and 0.98. (--) Dashed lines are the dataset-weighted average AUC scores per feature set.

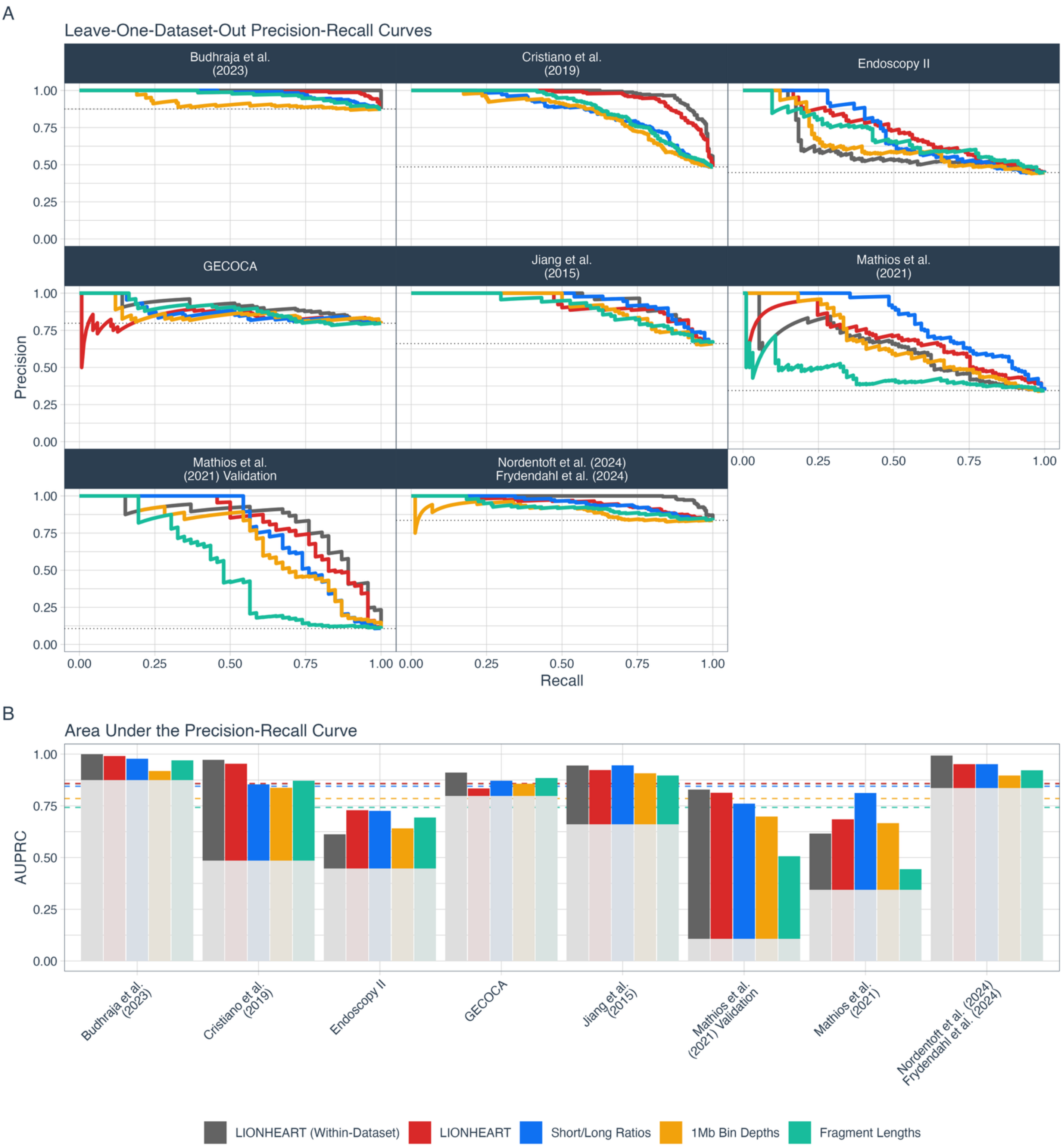


**Supplementary Figure 6**: Precision-Recall (PR) per dataset and feature set from leave-one-dataset-out cross-validation. A) PR curves. (..) Dotted lines are the baselines (prevalence). B) Area under the PR curve (AUPRC) scores. The bars are greyed out underneath the baselines (prevalence). (--) Dashed lines are the dataset-weighted average AUPRC scores per feature set.

##### *Cross-dataset generalization*

To assess the cross-dataset generalization and inter-dynamics of the datasets, we ran leave-one-dataset-out nested cross-validation for all combinations of datasets. The following plots show the ROC AUC scores per test dataset (panes on the y-axis (columns)) when including or excluding a given dataset (panes on the x-axis (rows)) in the model training.

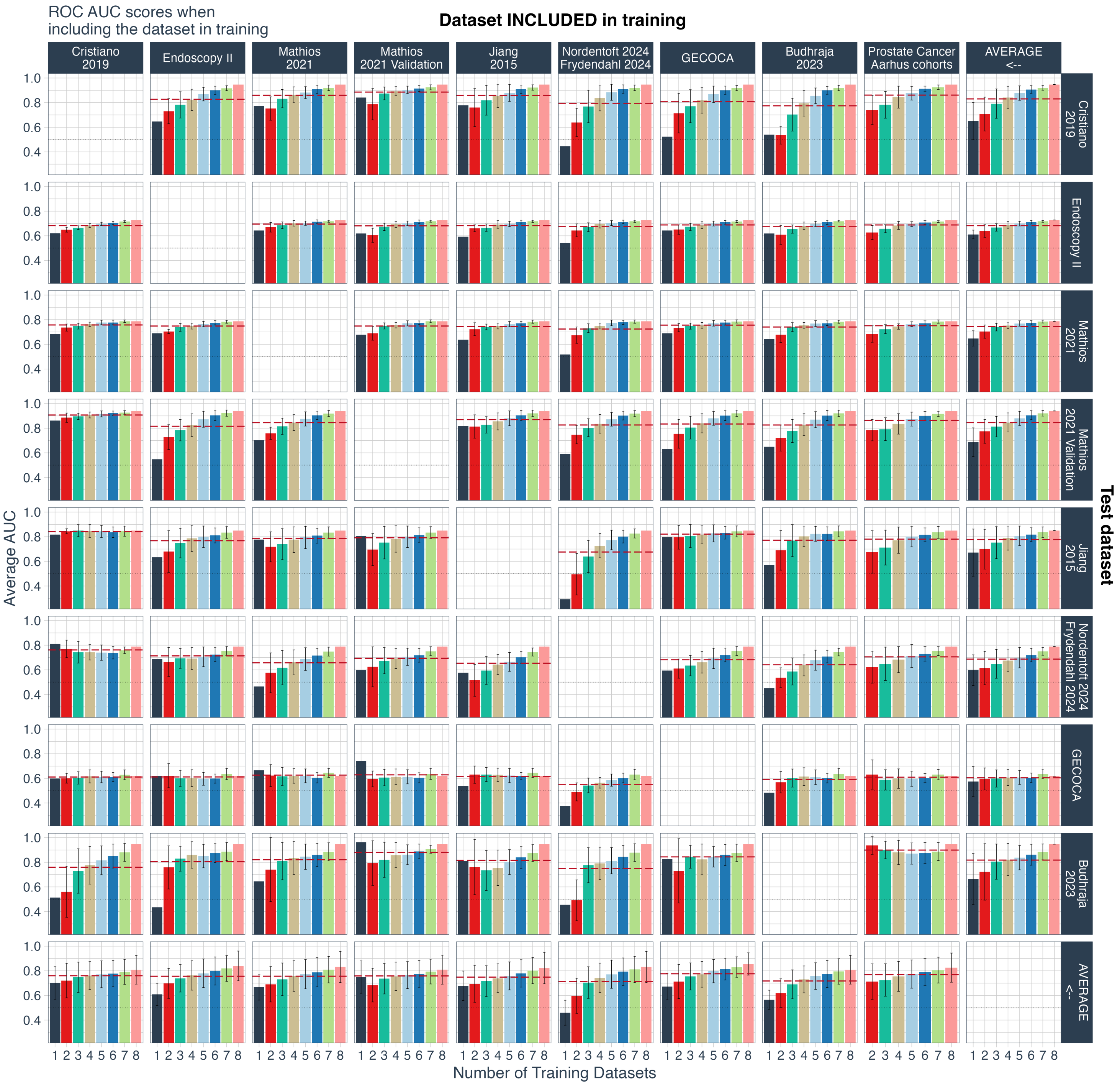


**Supplementary Figure 7**: ROC AUC scores for pan-cancer vs. non-cancer-control LIONHEART classifiers that *include* a given dataset (column panes) in training and predict the cancer statuses in the test dataset (row panes). Based on leave-one-dataset-out nested cross-validation results for all combinations of the datasets. The bottom row contains the average ROC AUC scores per included training dataset. The rightmost column contains the average ROC AUC scores for each test dataset. The Prostate Cancer (Aarhus cohorts) dataset was only used for training, along with at least one other dataset, and thus starts at 2 on the x-axis. (--) The dashed lines are the mean AUC scores. (..) The dotted lines are the chance-level baseline scores. The bar colors specify the number of training datasets (i.e., the x-axis value).

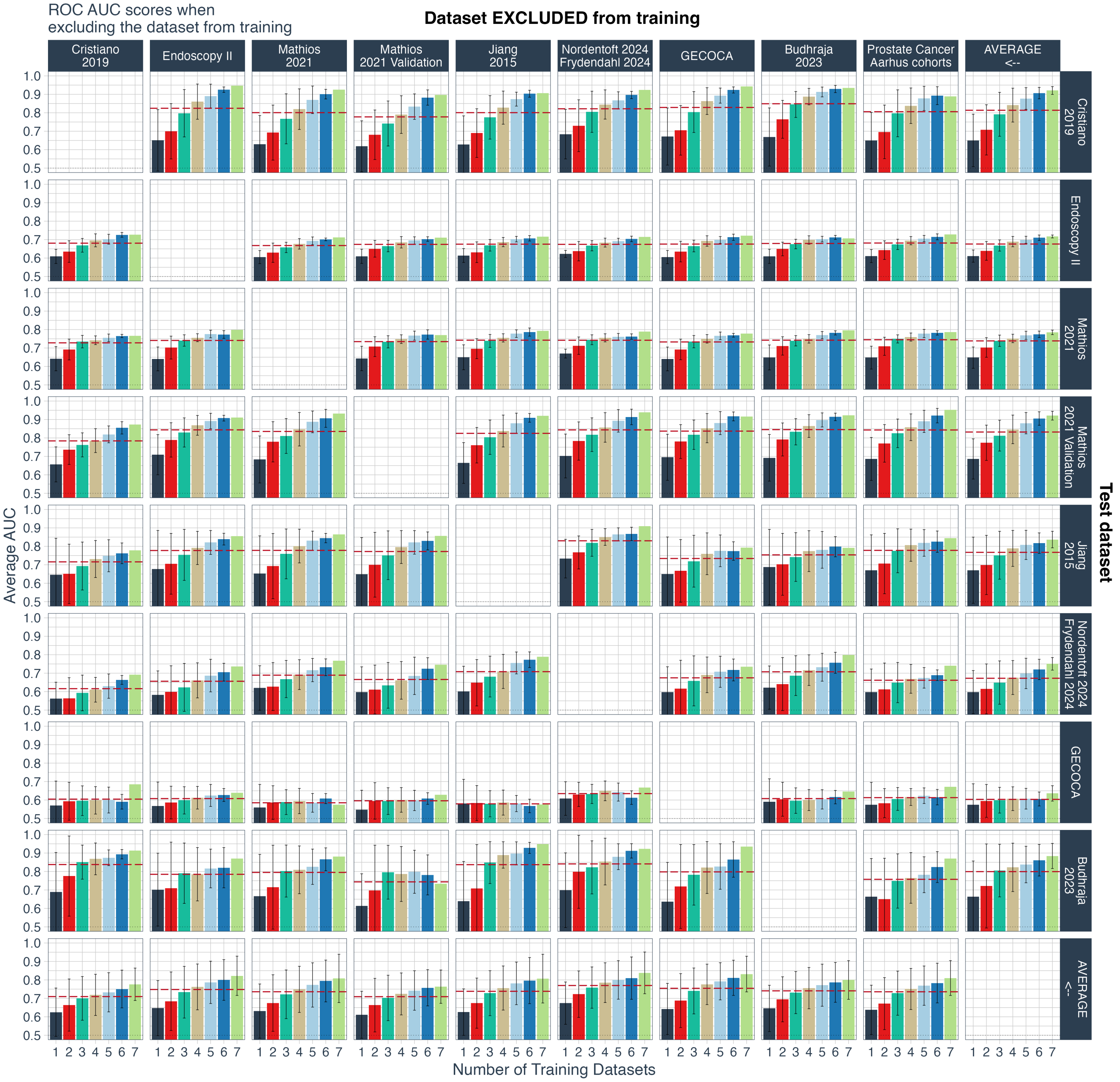


**Supplementary Figure 8**: ROC AUC scores for pan-cancer vs. non-cancer-control LIONHEART classifiers that *exclude* a given dataset (column panes) from training and predict the cancer statuses in the test dataset (row panes). I.e., the scores from all other training dataset combinations. Based on leave-one-dataset-out nested cross-validation results for all combinations of the datasets. The bottom row contains the average ROC AUC scores per excluded training dataset. The rightmost column contains the average ROC AUC scores for each test dataset. (--) The dashed lines are the mean AUC scores. (..) The dotted lines are the chance-level baseline scores. The bar colors specify the number of training datasets (i.e., the x-axis value).

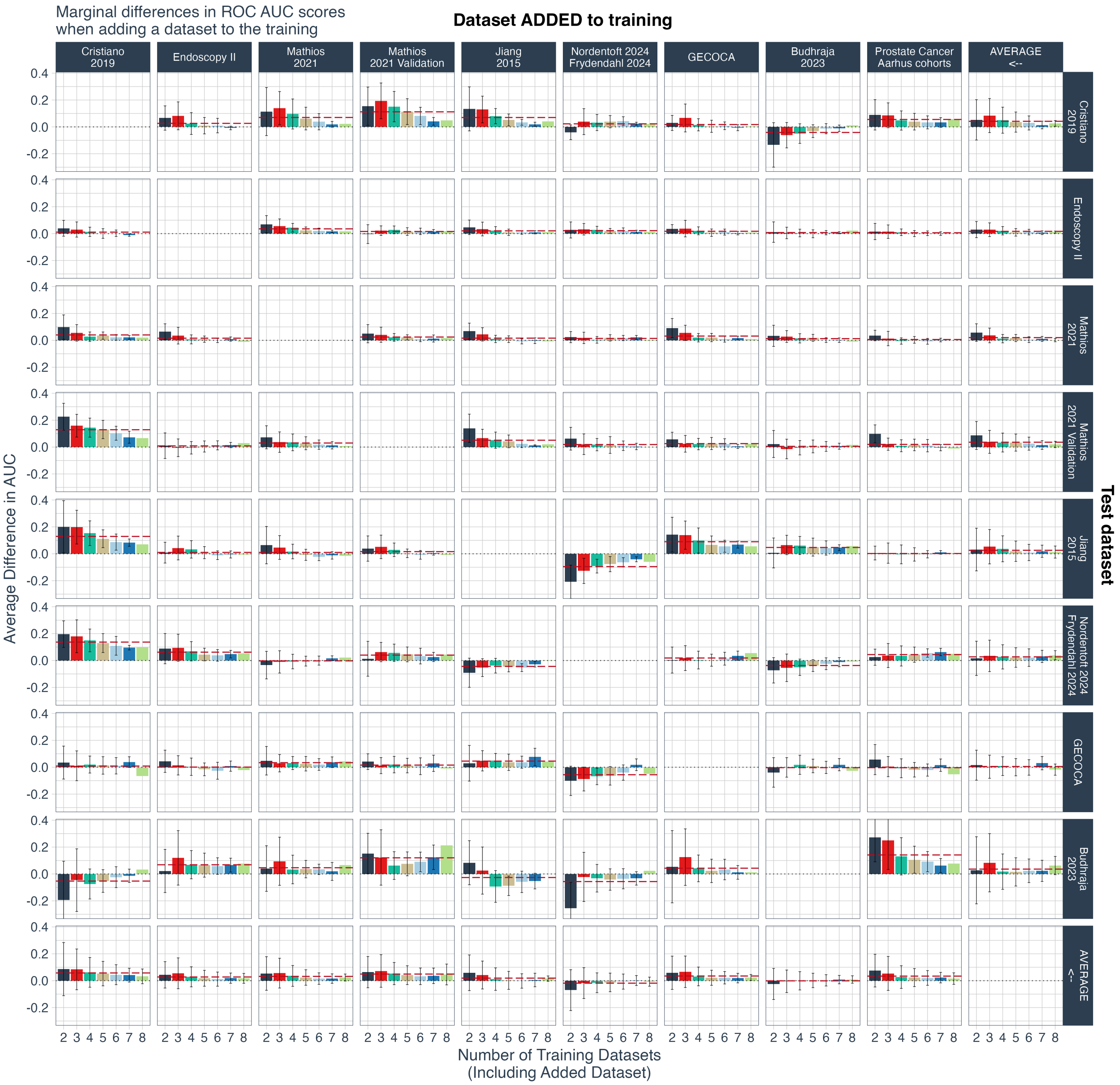


**Supplementary Figure 9**: Marginal differences in ROC AUC scores when *adding* a given dataset (column panes) to the training of pan-cancer vs. non-cancer-control LIONHEART classifiers and predicting the cancer statuses in the test dataset (row panes). Based on leave-one-dataset-out nested cross-validation results for all combinations of the datasets. The bars represent the average differences in ROC AUC scores between combinations that include the dataset and the same combinations with just that dataset removed. The bottom row contains the average marginal differences per added training dataset. (--) Dashed lines are the mean differences. Error bars represent one standard deviation. The bar colors specify the number of training datasets (i.e., the x-axis value).

| Predictor | Average Marginal Difference Δ | t-value | p-value | Corrected p-value |
|---|---|---|---|---|
| Nordentoft et al., 2024; Frydendahl et al., 2024 | -0.015 | -5.533 | <0.001 | <0.001 |
| Cristiano et al. 2019 | 0.060 | 15.852 | <0.001 | <0.001 |
| GECOCA | 0.034 | 12.981 | <0.001 | <0.001 |
| Mathios et al. 2021 Validation | 0.049 | 16.979 | <0.001 | <0.001 |
| Mathios et al. 2021 | 0.032 | 12.348 | <0.001 | <0.001 |
| Endoscopy II | 0.027 | 10.535 | <0.001 | <0.001 |
| Prostate Cancer, Aarhus cohorts | 0.032 | 11.662 | <0.001 | <0.001 |
| Jiang et al. 2015 | 0.014 | 4.990 | <0.001 | <0.001 |
| Budhraja et al. 2023 | -0.001 | -0.356 | 0.722 | 0.722 |

**Supplementary Table 4**: Summaries of per-dataset one-sample t-tests of marginal differences (Δ) in ROC AUC scores when adding a dataset to the training of pan-cancer vs. non-cancer-control LIONHEART classifiers. Based on leave-one-dataset-out nested cross-validation results for all combinations of the datasets. Marginal differences refer to the differences in ROC AUC scores between combinations that include the dataset and the same combinations with just that dataset removed. The p-values are corrected using the Benjamini-Hochberg procedure.

#### Pan-cancer classifier vs. cancer-type-specific classifier

| Test Dataset | Cross-Dataset ROC AUC | | Within-Dataset ROC AUC | N - | N + |
|---|---|---|---|---|---|
| | Single Cancer Model | Pan-Cancer Model | Single Cancer Model | | |
| **Task** | **Lung Cancer vs. Controls** | | | | |
| Cristiano et al., 2019 | 0.913 | 0.959 | 0.950 | 244 | 35 |
| Mathios et al., 2021 | 0.730 | 0.787 | 0.735 | 177 | 93 |
| Mathios et al., 2021 Validation Cohort | 0.882 | 0.940 | 0.962 | 385 | 46 |
| Jiang et al, 2015 | 0.926 | 0.939 | 0.616 | 38 | 10 |
| **Average** | 0.863 | 0.906 | 0.816 | | |
| **Weighted Average** | 0.853 | 0.905 | 0.883 | | |
| **Task** | **Colorectal Cancer vs. Controls** | | | | |
| Cristiano et al., 2019 | 0.939 | 0.961 | 0.993 | 244 | 27 |
| Endoscopy II | 0.718 | 0.728 | 0.6015 | 141 | 114 |
| Nordentoft et al., 2024; Frydendahl et al., 2024 | 0.515 | 0.781 | 0.956 | 45 | 127 |
| Jiang et al, 2015 | 0.826 | 0.729 | 0.522 | 38 | 10 |
| GECOCA | 0.536 | 0.618 | 0.729 | 34 | 134 |
| **Average** | 0.707 | 0.764 | 0.760 | | |
| **Weighted Average** | 0.717 | 0.787 | 0.804 | | |

**Supplementary Table 5**: ROC AUC scores for single-cancer-type vs. non-cancer-control classifiers (i.e., lung cancer vs. non-cancer-control and colorectal cancer vs. non-cancer-control). We compare to the predictions of the same samples made by the pan-cancer vs. non-cancer-control classifier from Supplementary Table 3. **Cross-dataset**: From nested leave-one-dataset-out cross-validation. Each value is the test score when trained on the other datasets and tested on the given dataset. The controls from datasets without the cancer type in question (lung or colorectal) were included in the training data. **Within-dataset**: Per-dataset regular nested cross-validation results for comparison. **N-**: Number of control subjects; **N+**: Number of lung/colorectal subjects.

#### Generalization to out-of-dataset cancer types

| Cancer Type | Cross-Cancer Type ROC AUC | | | | N - | N + |
|---|---|---|---|---|---|---|
| | LIONHEART | Depth in 1Mb bins | Short/Long Ratios in 1Mb bins | Fragment Length Distribution | | |
| Colorectal | 0.834 | 0.846 | 0.921 | 0.796 | 314.0 | 412 |
| Prostate | 0.884 | 0.916 | 0.935 | 0.674 | 275.0 | 361 |
| Lung | 0.670 | 0.607 | 0.555 | 0.705 | 141.6 | 184 |
| Bladder | 0.955 | 0.945 | 0.945 | 0.926 | 77.0 | 102 |
| Breast | 0.928 | 0.902 | 0.785 | 0.768 | 77.8 | 101 |
| Melanoma | 0.853 | 0.672 | 0.769 | 0.880 | 55.0 | 73 |
| Bile Duct | 0.954 | 0.802 | 0.872 | 0.871 | 46.8 | 61 |
| Pancreatic | 0.769 | 0.501 | 0.692 | 0.739 | 27.4 | 34 |
| Hepatocellular Carcinoma | 0.917 | 0.867 | 0.910 | 0.916 | 25.8 | 34 |
| Ovarian | 0.972 | 0.920 | 0.839 | 0.7165 | 21.8 | 28 |
| Gastric | 0.917 | 0.812 | 0.612 | 0.668 | 20.8 | 27 |
| Glioblastoma | 0.960 | 0.790 | 0.743 | 0.810 | 9.0 | 12 |
| Head and Neck Squamous Cell Carcinoma | 0.827 | 0.887 | 0.846 | 0.924 | 7.0 | 10 |
| Nasopharyngeal Carcinoma | 0.914 | 0.886 | 0.857 | 0.937 | 7.0 | 10 |
| **Average** | **0.882** | 0.811 | 0.806 | 0.809 | | |
| **Weighted Average** | **0.853** | 0.827 | 0.845 | 0.7678 | | |

**Supplementary Table 6:** ROC AUC scores per feature type and cancer type for cancer vs. non-cancer-control models from leave-one-cancer-type-out nested cross-validation with all datasets merged. **N-**: Average number of sampled non-cancer control subjects; **N+**: Number of cancer subjects.

#### External validation

We performed external validation on the dataset from Zhu et al. (2025).

| True Class | Stage | Prediction | | Fraction Correct |
|---|---|---|---|---|
| | | Control | Cancer | |
| Control | | 41 | 12 | 0.774 |
| Breast Cancer | N/A | 0 | 10 | 1.000 |
| Breast Cancer | II | 0 | 1 | 1.000 |
| Breast Cancer | IV | 2 | 18 | 0.900 |
| Colorectal Cancer | II | 0 | 5 | 1.000 |
| Colorectal Cancer | III | 0 | 11 | 1.000 |
| Colorectal Cancer | IV | 2 | 22 | 0.917 |
| Gastric Cancer | III | 0 | 1 | 1.000 |
| Gastric Cancer | IV | 7 | 17 | 0.708 |
| All Cancer | II | 0 | 6 | 1.000 |
| All Cancer | III | 0 | 12 | 1.000 |
| All Cancer | IV | 11 | 57 | 0.838 |

**Supplementary Table 7**: Prediction counts, overall and stratified on cancer type and stage. The probability threshold (0.49) was selected to maximize the expected sum of sensitivity and specificity (Max. Youden's J) and was calculated from the training data.

| Strata | ROC AUC | Sensitivity | Specificity (same controls) | Positive Predictive Value | Negative Predictive Value |
|---|---|---|---|---|---|
| **Overall** | 0.917 | 0.885 | 0.774 | 0.876 | 0.788 |
| **Cancer Type** | | | | | |
| Breast Cancer | 0.932 | 0.935 | 0.774 | 0.707 | 0.953 |
| Colorectal Cancer | 0.962 | 0.950 | 0.774 | 0.760 | 0.953 |
| Gastric Cancer | 0.826 | 0.720 | 0.774 | 0.600 | 0.854 |
| **Stage** | | | | | |
| II | 0.969 | 1.000 | 0.774 | 0.333 | 1.000 |
| III | 0.967 | 1.000 | 0.774 | 0.500 | 1.000 |
| IV | 0.893 | 0.838 | 0.774 | 0.826 | 0.788 |
| **Tumor Fraction Fragle Estimates** | | | | | |
| [0.0, 0.01] | 0.755 | 0.714 | 0.774 | 0.294 | 0.953 |
| (0.01, 0.10] | 0.821 | 0.750 | 0.774 | 0.667 | 0.837 |
| (0.10, 1.0) | 0.991 | 0.982 | 0.774 | 0.824 | 0.976 |
| **Tumor Fraction ichorCNA Estimates** | | | | | |
| [0.0, 0.03] | 0.814 | 0.706 | 0.774 | 0.667 | 0.804 |
| (0.03, 0.10] | 0.928 | 0.950 | 0.774 | 0.613 | 0.976 |
| (0.10, 1.0) | 0.995 | 1.000 | 0.774 | 0.778 | 1.000 |

**Supplementary Table 8**: Validation results overall and stratified by cancer type, stage, and tumor fraction intervals (Fragle- and ichorCNA-estimates separately). The probability threshold (0.49) was selected to maximize the expected sum of sensitivity and specificity (Max. Youden's J) and was calculated from the training data.

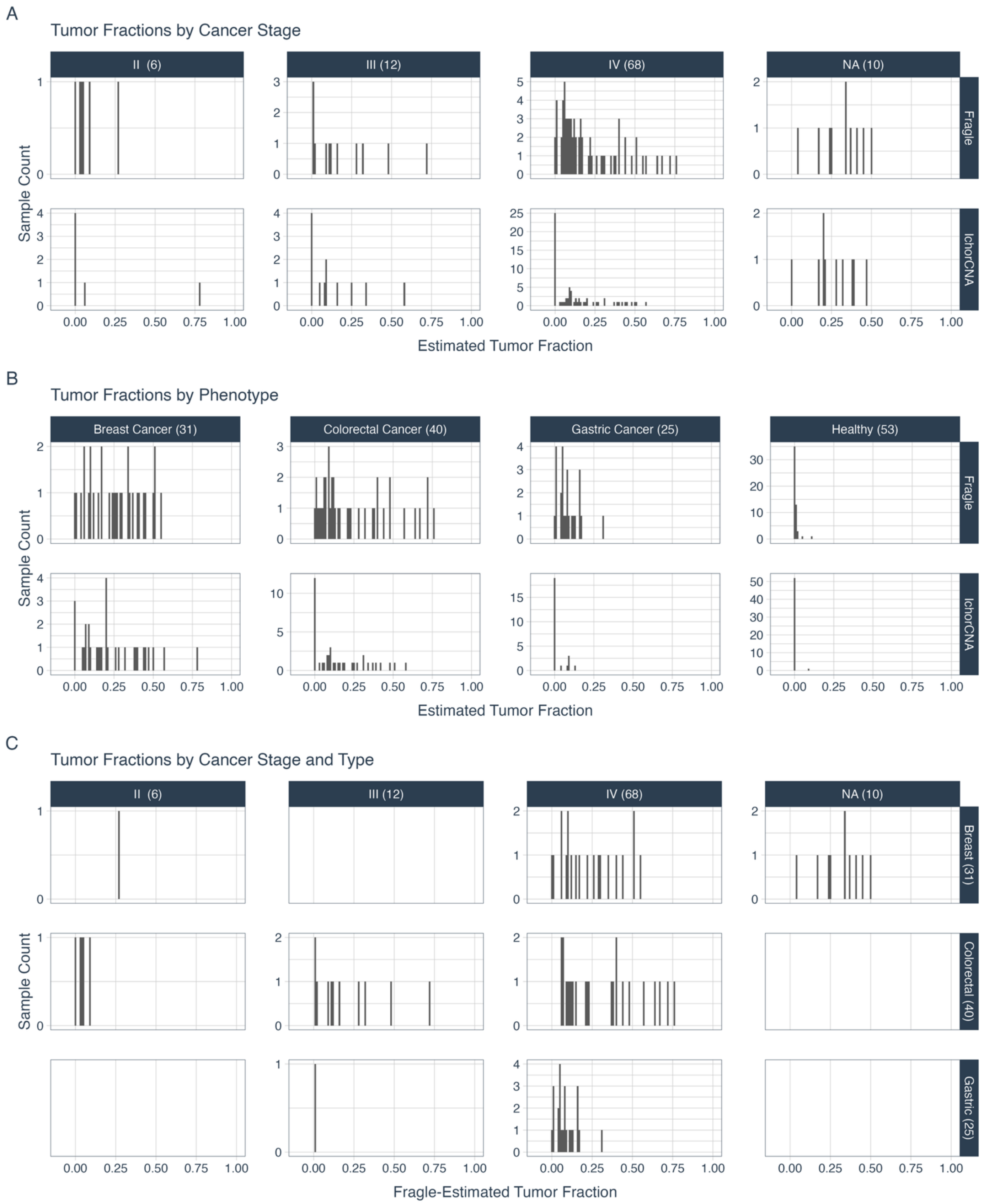


**Supplementary Figure 10:** Histograms of estimated tumor fractions (by IchorCNA or Fragle) in the external validation dataset from Zhu et al., (2023) per A) cancer stage, (B) phenotype (healthy or cancer type), and C) each combination of cancer stage and cancer type.

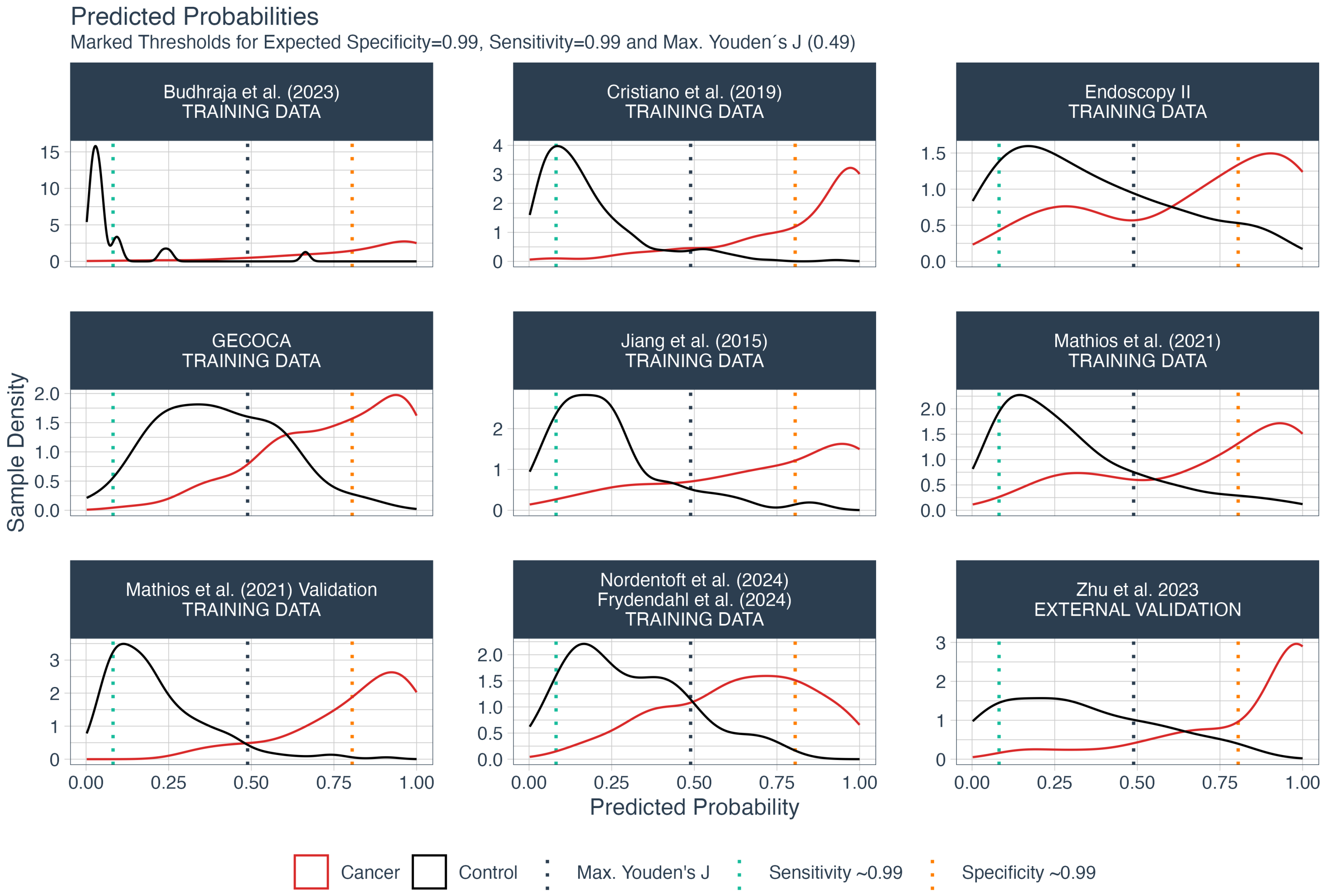


**Supplementary Figure 11:** Densities of predicted probabilities from the final pan-cancer classifier on its training and external validation datasets. Horizontal lines mark the thresholds yielding an expected (i.e., training-data-based) sensitivity and specificity of 0.99 and the expected maximum Youden's J threshold.

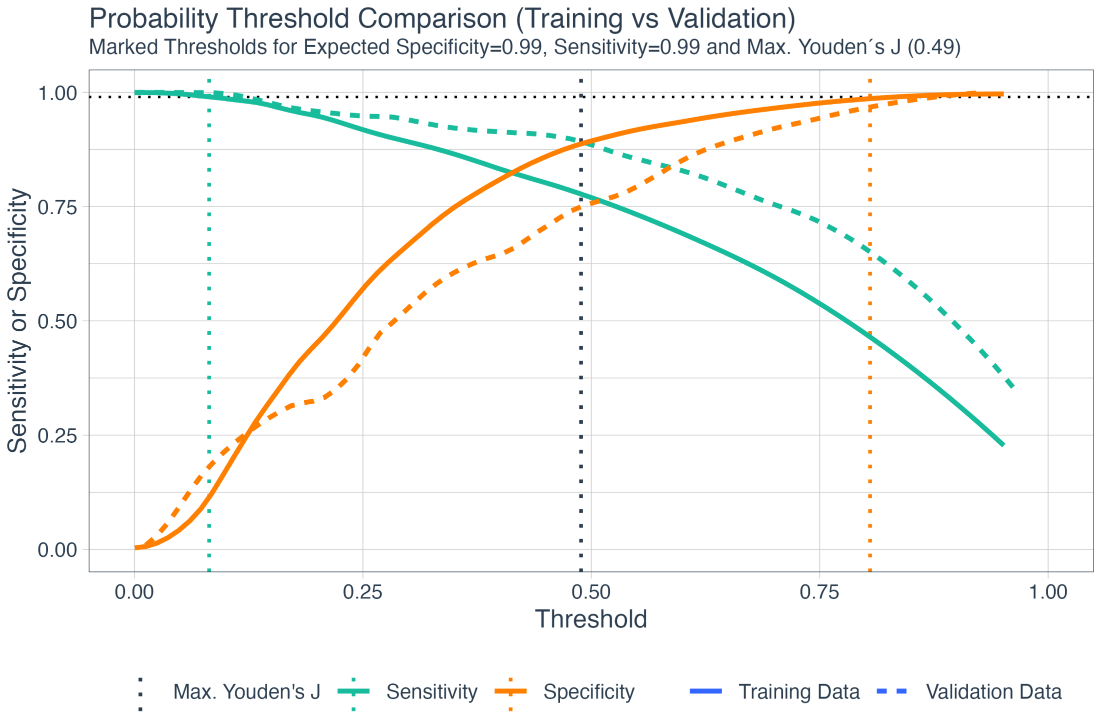


**Supplementary Figure 12:** Comparison of probability thresholds in the training and validation data with separate lines for sensitivities and specificities. Horizontal lines mark the thresholds yielding an expected (i.e., training-data-based) sensitivity and specificity of 0.99 and the expected maximum Youden's J threshold.

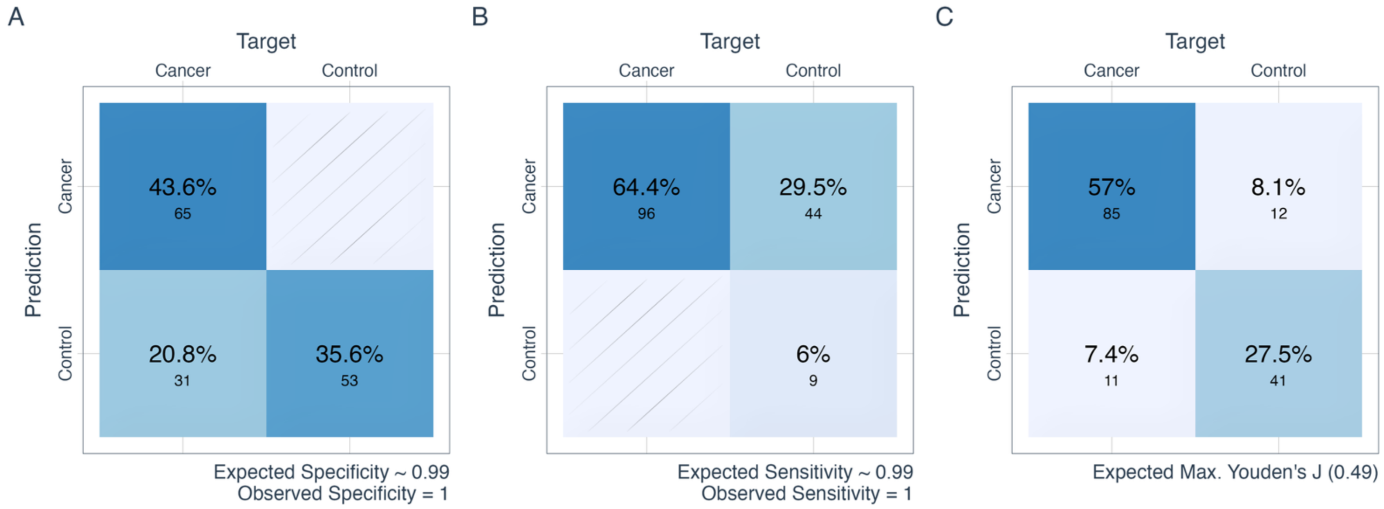


**Supplementary Figure 13:** Confusion matrices from the external validation. A and B compare expected and observed specificities (A) and sensitivities (B) with probability thresholds calculated on the *training* data and applied to the validation data. A) Confusion matrix using a probability threshold for an *expected specificity* of 0.99. B) Confusion matrix using a probability threshold for an *expected sensitivity* of 0.99. C) Confusion matrix using a probability threshold at the *expected maximum Youden's J*.

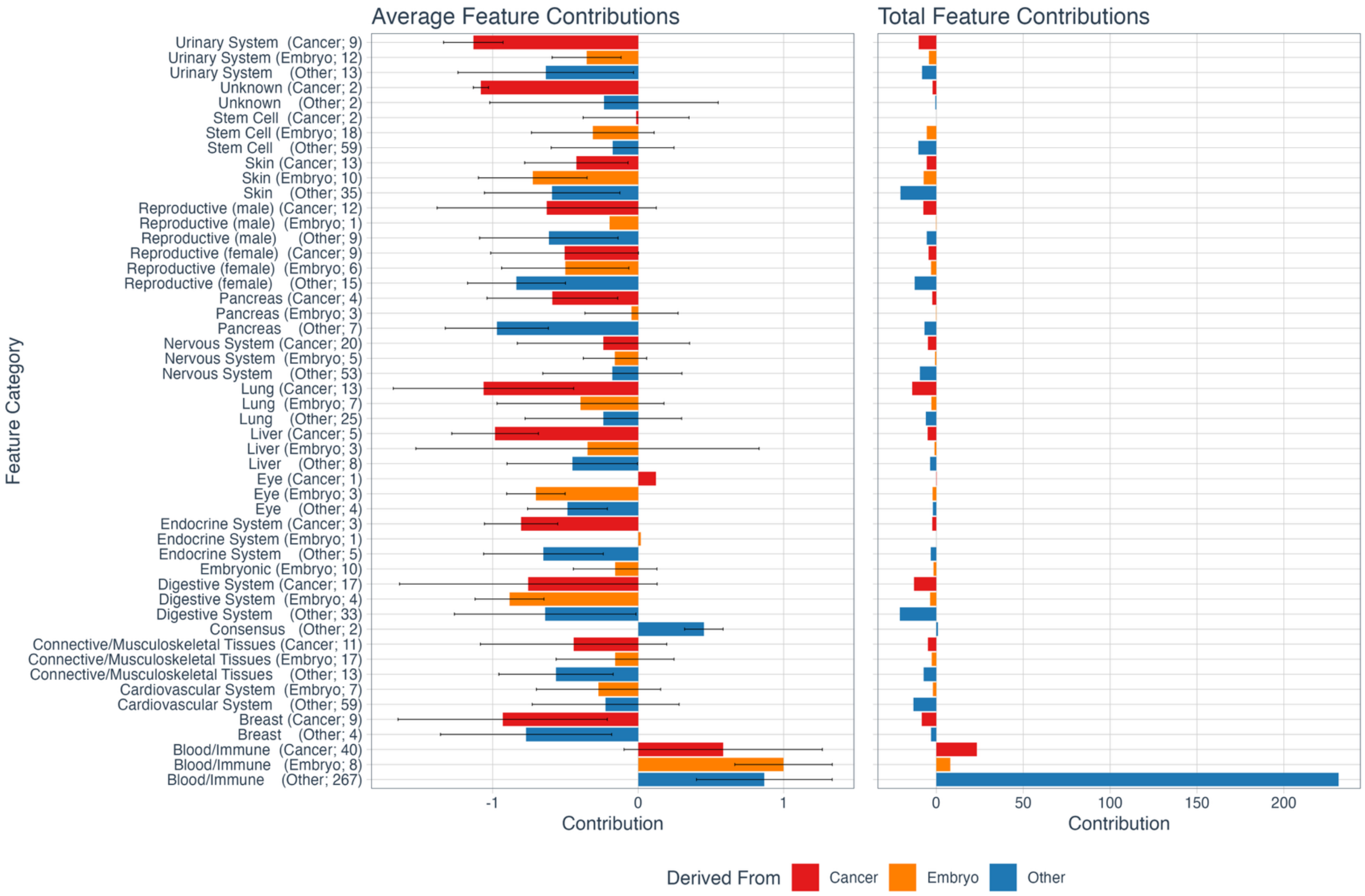


**Supplementary Figure 14:** Average (left) and total (right) contributions to the final pan-cancer classifier for each cell type category. Defined as the inverse transformation of the PCA transformation, standardization, and LASSO classification (i.e., $C = \beta_{LASSO} \cdot diag(\sigma) \cdot W_{PCA}$ , where $\beta_{LASSO}$ represents the model coefficients with shape (1, 85), $W_{PCA}$ represents the PCA components with shape (85, 898), and $\sigma$ represents the standard deviations used to scale during standardization with shape (85). The PCA components are already centered, so no inverse-centralization was necessary.) The blood/immune cell types contribute positively to the classification, meaning that higher LIONHEART scores for those cell types lead to a higher probability of cancer. The other cell type categories mainly contribute negatively to the classification. Error bars represent one standard deviation.

#### Methods

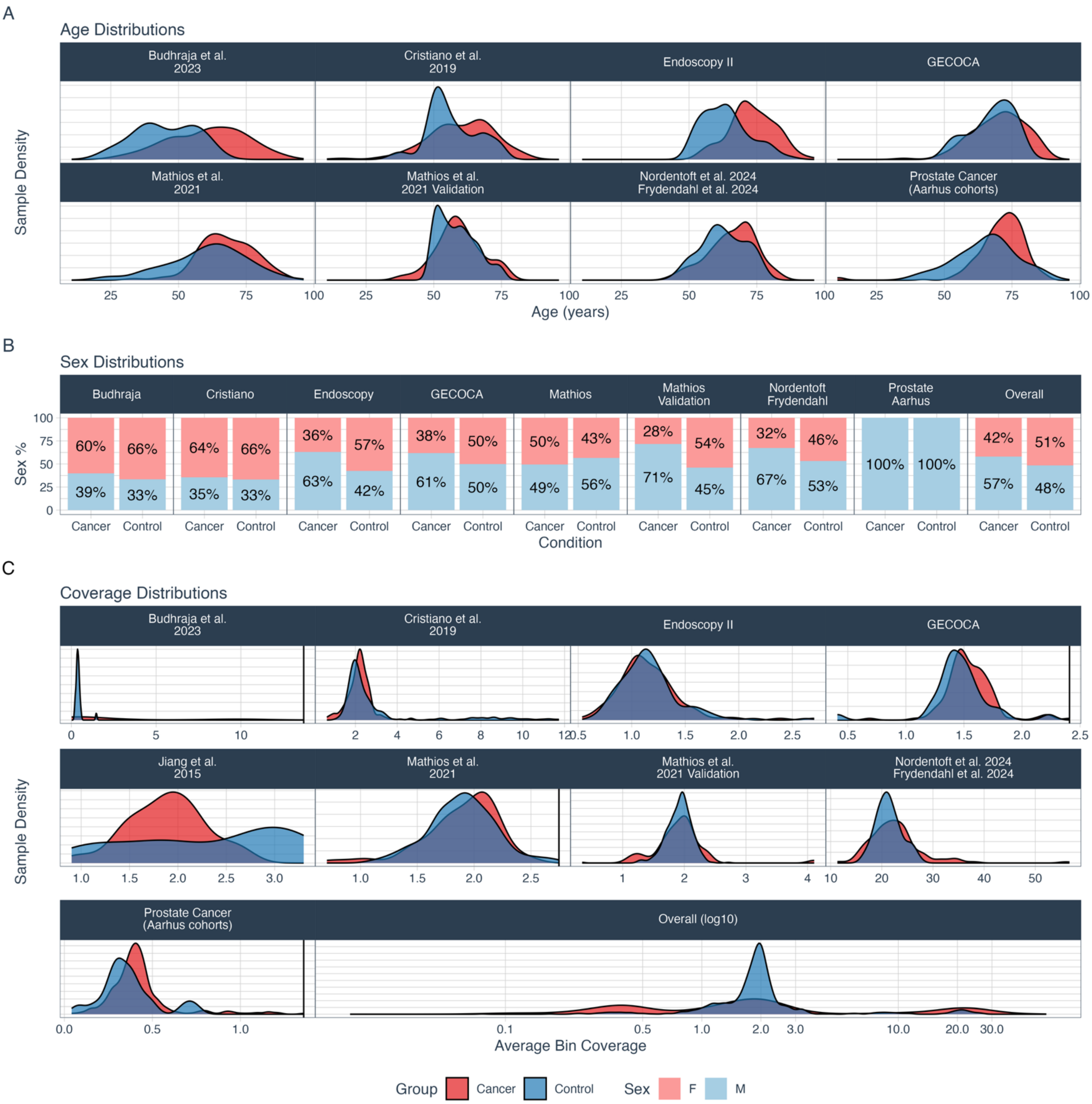


**Supplementary Figure 15**: A) Age densities per dataset and group. Cancer patients are older on average than control patients. No age data was available for the dataset from Jiang et al. B) Sex distributions per dataset and group. C) Average bin coverage per dataset and group before any corrections, but with truncation of extreme bin outliers. This is an estimate of the sequencing depth. **Exclusions**: 1 sample with an average coverage of 2.03 was excluded from the Prostate Cancer (Aarhus cohorts) plot. One sample with an average coverage of 5.65 was excluded from the Mathios et al. plot. Two samples with average coverages of 17.5 and 18.8 were excluded from the Budhraja et al. plot. 10 cancer samples with average coverages between 14.1 and 25.5 were excluded from the GECOCA plot. D) Overall average bin coverage with all samples. The x-axis is on a log10 scale.

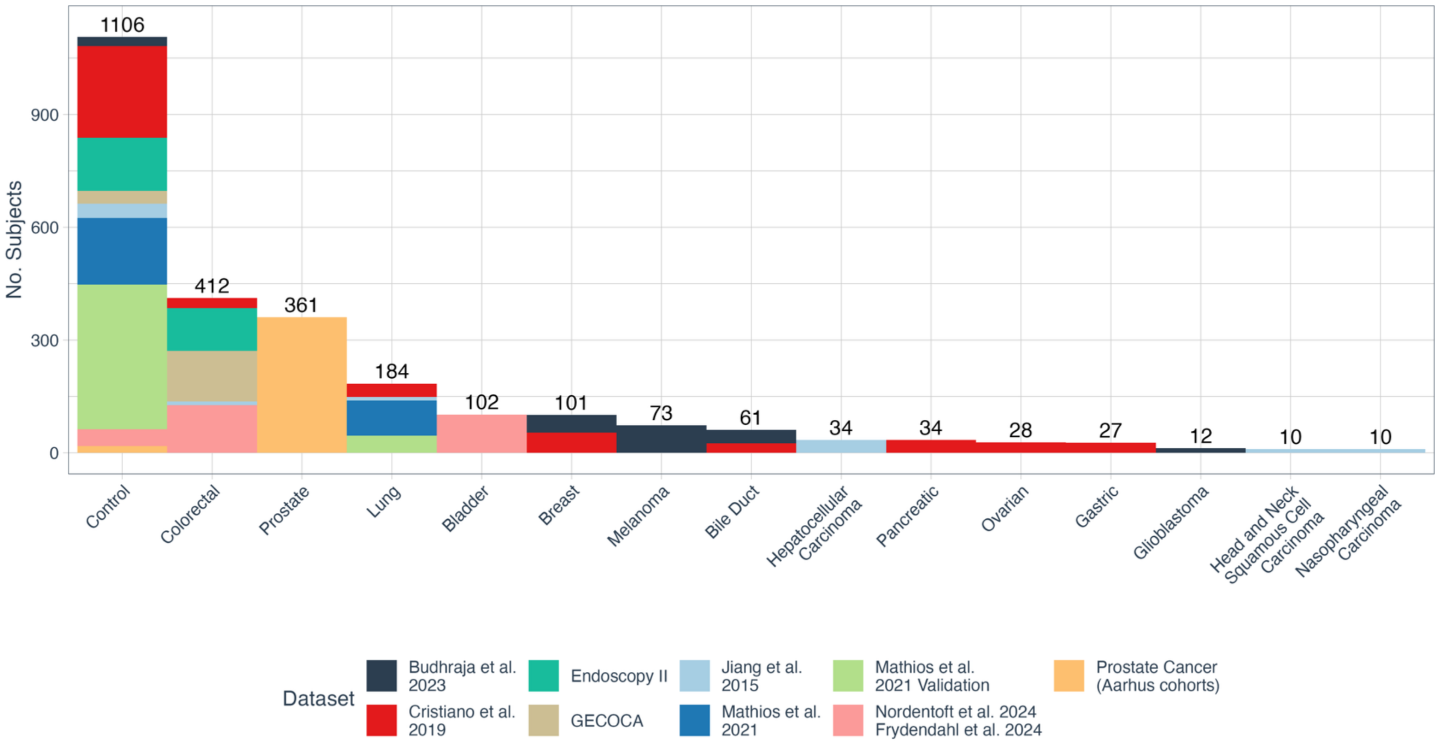


**Supplementary Figure 16**: Number of samples per condition and dataset (excluding the external validation dataset).

| Dataset | Target | Mean Bin Coverage | | | |
|---|---|---|---|---|---|
| | | Mean | Std. | Max. | Min. |
| Nordentoft et al., 2024; Frydendahl et al., (2024) | Cancer | 22.6 | 5.50 | 56.7 | 11.3 |
| Nordentoft et al., 2024; Frydendahl et al., (2024) | Control | 21.4 | 2.43 | 27.1 | 16.6 |
| Jiang et al, 2015 | Cancer | 1.89 | 0.40 | 2.74 | 0.90 |
| Jiang et al, 2015 | Control | 2.30 | 0.80 | 3.30 | 1.01 |
| Endoscopy II | Cancer | 1.16 | 0.31 | 2.70 | 0.54 |
| Endoscopy II | Control | 1.16 | 0.28 | 2.63 | 0.66 |
| Cristiano et al., 2019 | Cancer | 2.18 | 0.56 | 4.70 | 0.64 |
| Cristiano et al., 2019 | Control | 2.99 | 2.24 | 11.7 | 1.12 |
| Mathios et al., 2021 | Cancer | 1.89 | 0.33 | 2.50 | 0.70 |
| Mathios et al., 2021 | Control | 1.92 | 0.41 | 5.65 | 1.10 |
| Mathios et al., 2021 Validation Cohort | Cancer | 1.92 | 0.45 | 4.11 | 1.09 |
| Mathios et al., 2021 Validation Cohort | Control | 1.91 | 0.22 | 2.71 | 0.34 |
| Budhraja et al., 2023 | Cancer | 3.32 | 4.38 | 18.82 | 0.02 |
| Budhraja et al., 2023 | Control | 0.42 | 0.24 | 1.47 | 0.22 |
| Prostate Cancer (Aarhus cohorts) | Cancer | 0.43 | 0.20 | 2.03 | 0.04 |
| Prostate Cancer (Aarhus cohorts) | Control | 0.36 | 0.16 | 0.74 | 0.08 |
| GECOCA | Cancer | 2.89 | 4.86 | 25.4 | 0.69 |
| GECOCA | Control | 1.46 | 0.27 | 2.23 | 0.41 |
| Zhu et al. 2023 | Cancer | 1.14 | 0.22 | 1.56 | 0.54 |
| Zhu et al. 2023 | Control | 1.43 | 0.29 | 2.09 | 1.11 |

**Supplementary Table 9**: Mean bin coverage statistics per dataset and group before any corrections but with truncation of extreme bin outliers.

#### Processing cfDNA samples

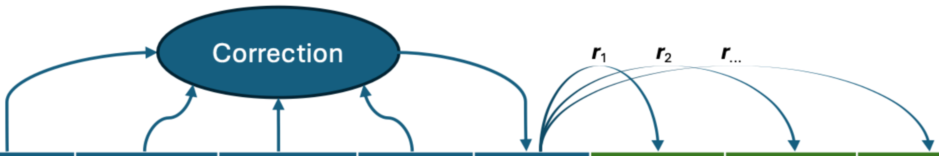


| Chromosome | Start | End | Fragment Coverage | GC Fraction (100bp Context) | Average Overlapping Insert Size | Average Coverage ($5^{+}$Mbp Context) | Corrected Coverage | % Open Chromatin Cell Type 1 | % Open Chromatin Cell type 2 | % Open Chromatin Cell Type ... |
|---|---|---|---|---|---|---|---|---|---|---|
| **chr1** | 20170 | 20180 | 14.3 | 0.63 | 167.3 | 2.37 | 13.2 | 0.9 | 0.0 | 0.0 |
| **chr1** | 20180 | 20190 | 3.2 | 0.52 | 163.2 | 2.37 | 3.9 | 1.0 | 0.0 | 0.0 |
| **chr1** | 20190 | 20200 | 0.0 | 0.38 | 159.2 | 2.37 | 0.0 | 1.0 | 0.3 | 0.0 |
| **...** | ... | ... | ... | ... | ... | ... | ... | ... | ... | ... |

**Supplementary Table 11**: Illustration of the relationship between variables used in the cfDNA processing with hypothetical data per 10bp bin. Purple: Chromosome coordinates (Chromosome, Start, End). Blue: Coverage and correction variables (average position-overlapping fragment coverage, GC fraction [0., 1.] in a 100bp context, average overlapping insert size). Green: Fraction of the positions that overlap an open chromatin site for each of the 898 cell types. **Steps**: 1) We truncate extreme outliers in the fragment coverage. 2) We reduce the GC bias in the fragment coverage. 3) We homogenize the distributions of overlapping insert sizes via corrections to the fragment coverage that remove noise and skewness and shift the mean to 166bp. 4) We reduce the effect of copy number alterations by dividing the fragment coverage by the average coverage in all overlapping 5Mb bins with a 500kb stride. 5) We calculate the Pearson correlation between the corrected fragment coverage and the open chromatin site fractions for each of the 898 cell types.

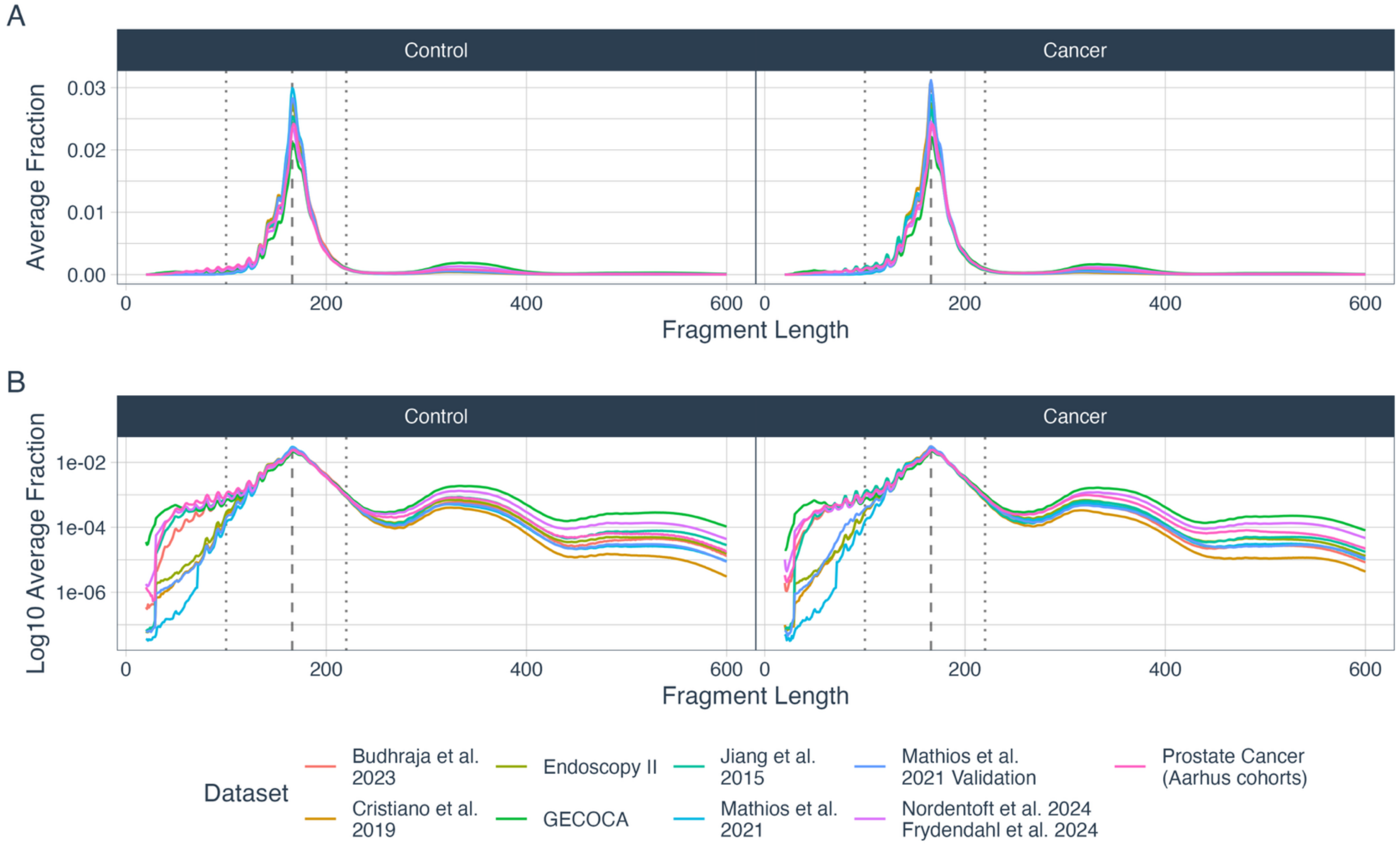


**Supplementary Figure 17**: Average fragment length distributions per dataset and cancer status for the 20-600bp range. A) Sum-to-one normalized average distributions. B) Log10-scaled sum-to-one normalized average distributions. (…) Dotted lines represent the 100bp and 220bp cutoffs used in the method. (---) Dashed line represents the 166bp expected mean fragment length.

*Bias correction*

The following GC fraction bin edges were used in the bin-based GC correction:

[0.00, 0.20, 0.22, 0.24, 0.25, 0.26, 0.27, 0.28, 0.29, 0.30, 0.31, 0.32, 0.33, 0.34, 0.35, 0.36, 0.37, 0.38, 0.39, 0.40, 0.41, 0.42, 0.43, 0.44, 0.45, 0.46, 0.47, 0.48, 0.49, 0.50, 0.51, 0.52, 0.53, 0.54, 0.55, 0.56, 0.57, 0.58, 0.59, 0.61, 0.62, 0.64, 0.67, 1.00]

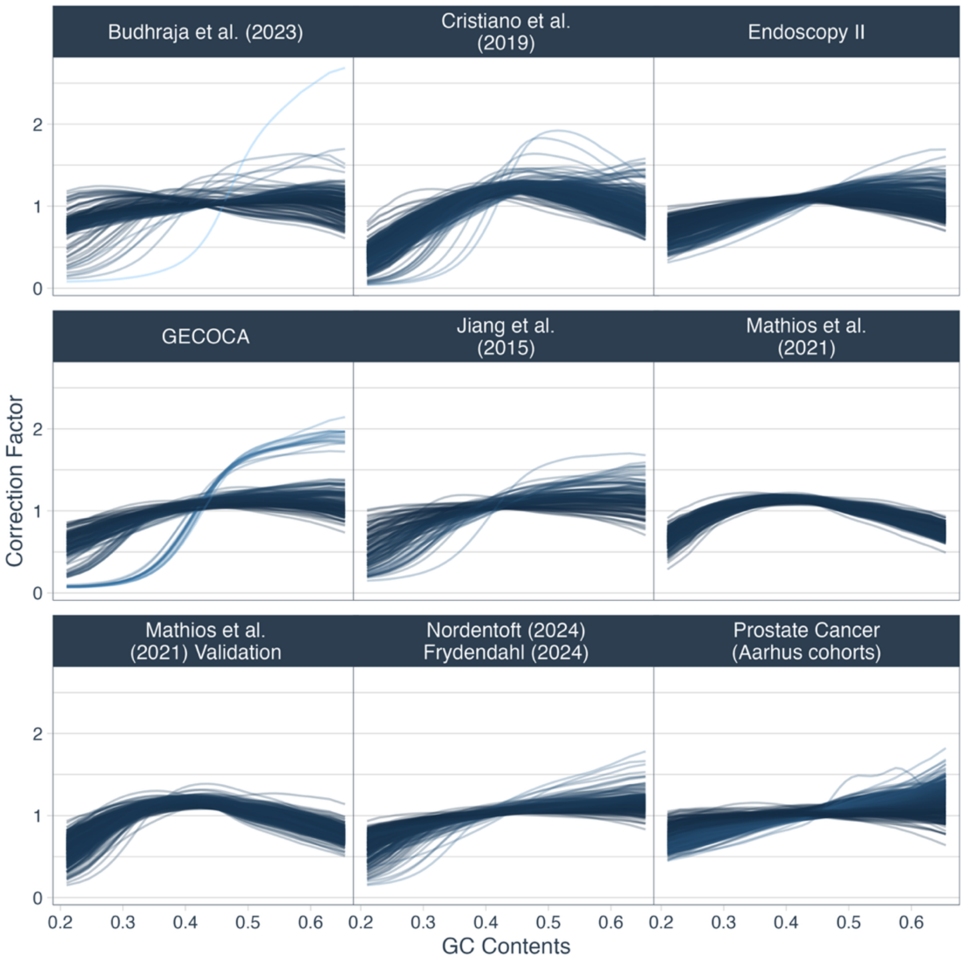


**Supplementary Figure 18**: GC correction factors. One line per sample. During correction, each coverage value is divided by the correction factor (y-axis) for the GC content (x-axis) in its 100bp context.

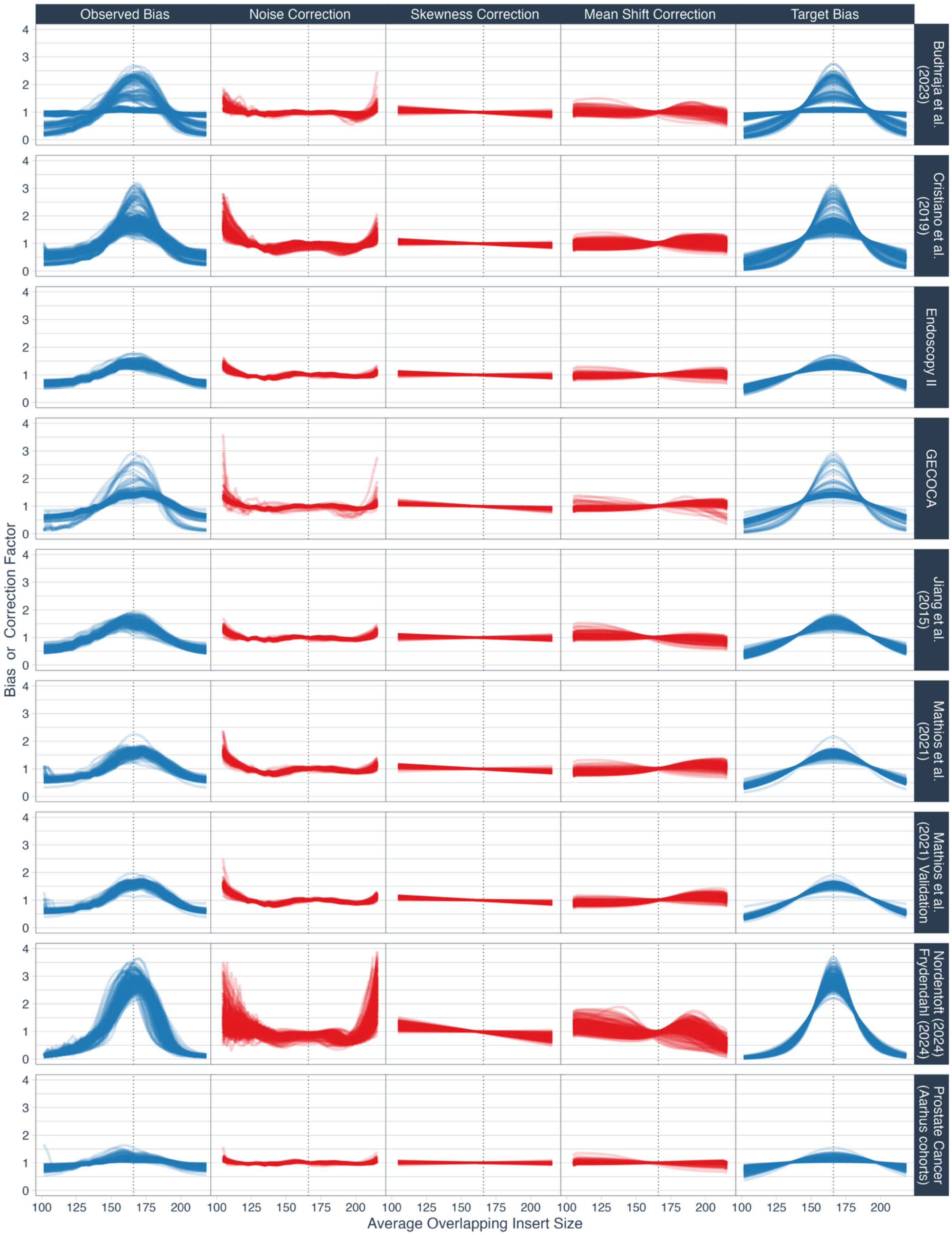
Observed Bias
Noise Correction
Skewness Correction
Mean Shift Correction
Target Bias
Budhraja et al. (2023)
Cristiano et al. (2019)
Endoscopy II
GECOCA
Jiang et al. (2015)
Mathios et al. (2021)
Mathios et al. (2021) Validation
Nordentoft (2024) Frydendahl (2024)
Prostate Cancer (Aarhus cohorts)
Bias or Correction Factor
Average Overlapping Insert Size
0
1
2
3
4
100
125
150
175
200

**Supplementary Figure 19**: Biases (blue) and correction factors (red) for the average overlapping insert size correction. Three corrections to the bin coverages transform the observed bias (first column) into the target bias (last column). This reduces skewness and sets the same mean for all the biases. Horizontal dotted line: The target mean at 166bp.

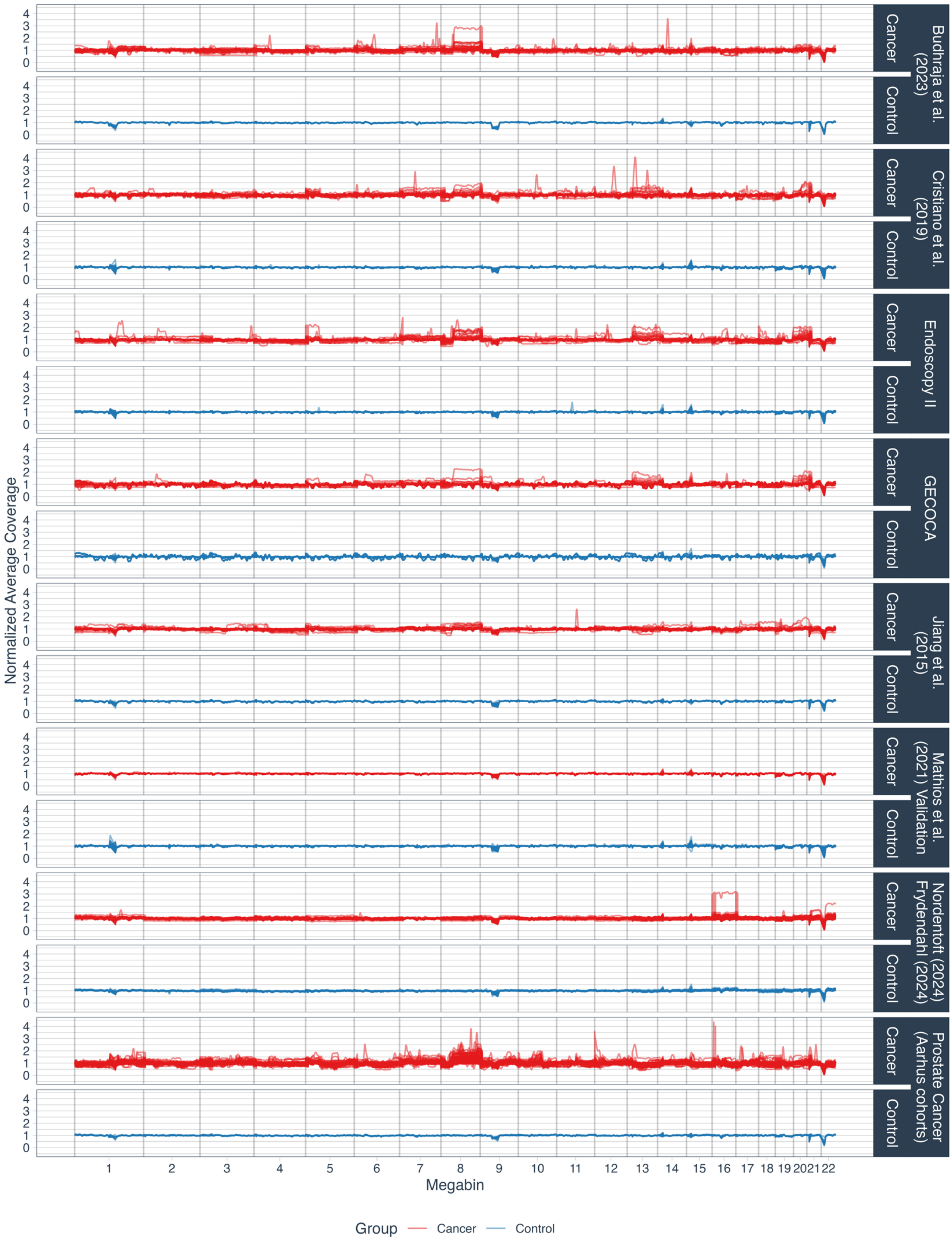

Budhraja et al. (2023)
Cristiano et al. (2019)
Endoscopy II
GECOCA
Jiang et al. (2015)
Mathios et al. (2021) Validation
Nordentoft (2024) Frydendahl (2024)
Prostate Cancer (Aarhus cohorts)
Cancer
Control
Normalized Average Coverage
Megabin
Group
Cancer
Control

**Supplementary Figure 20**: Normalized bin scaling factors. One average coverage value per 500kb stride, averaged from all overlapping 5Mb windows.

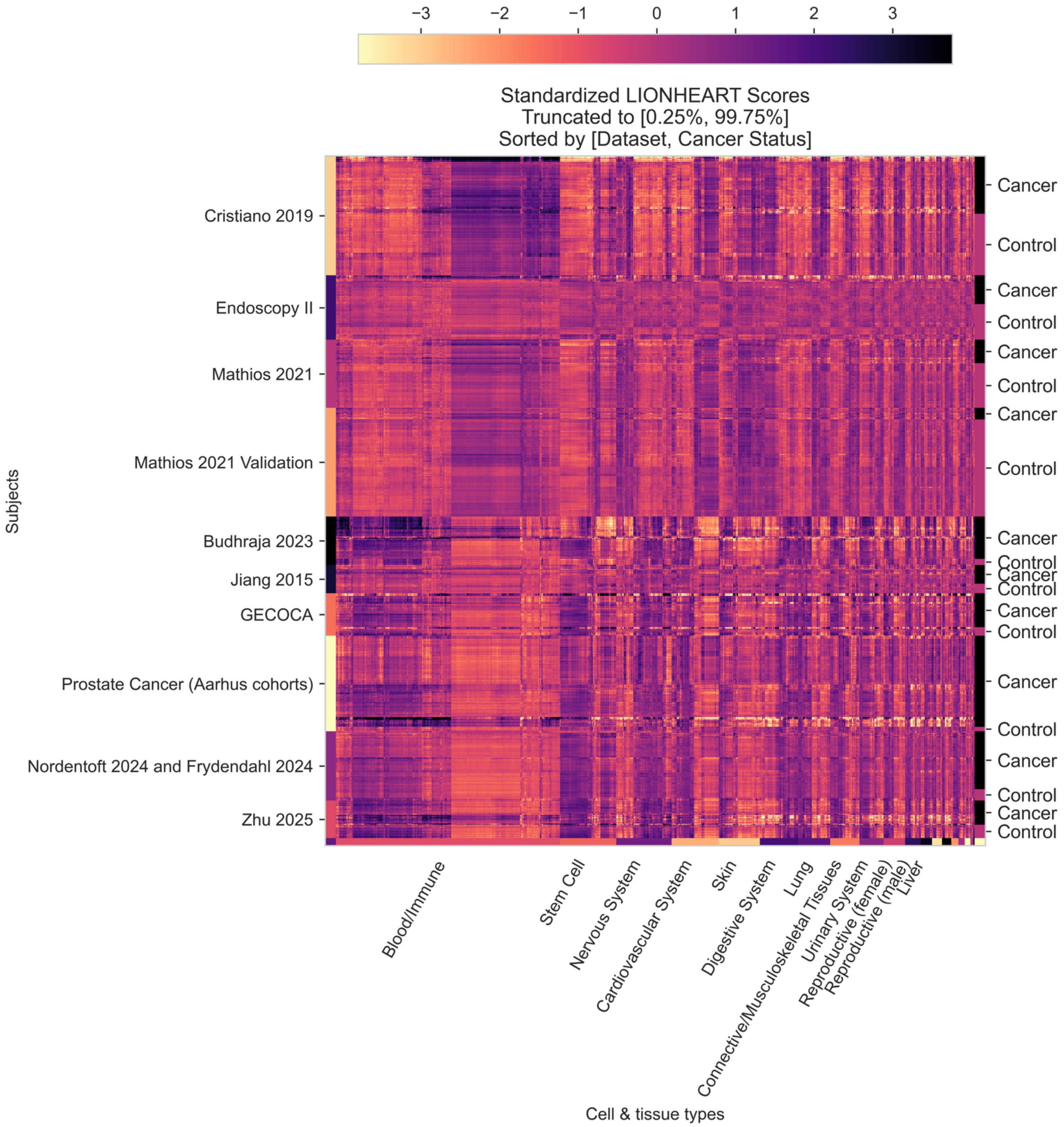


**Supplementary Figure 21**: Standardized LIONHEART scores. Every row is a sample, and every column is a cell type. Extreme outliers were truncated for visualization purposes. The clear lab / cohort effects support the need for multiple diverse datasets to learn the generalizing cancer/control signal. Some of these differences may be exaggerated by the standardization.

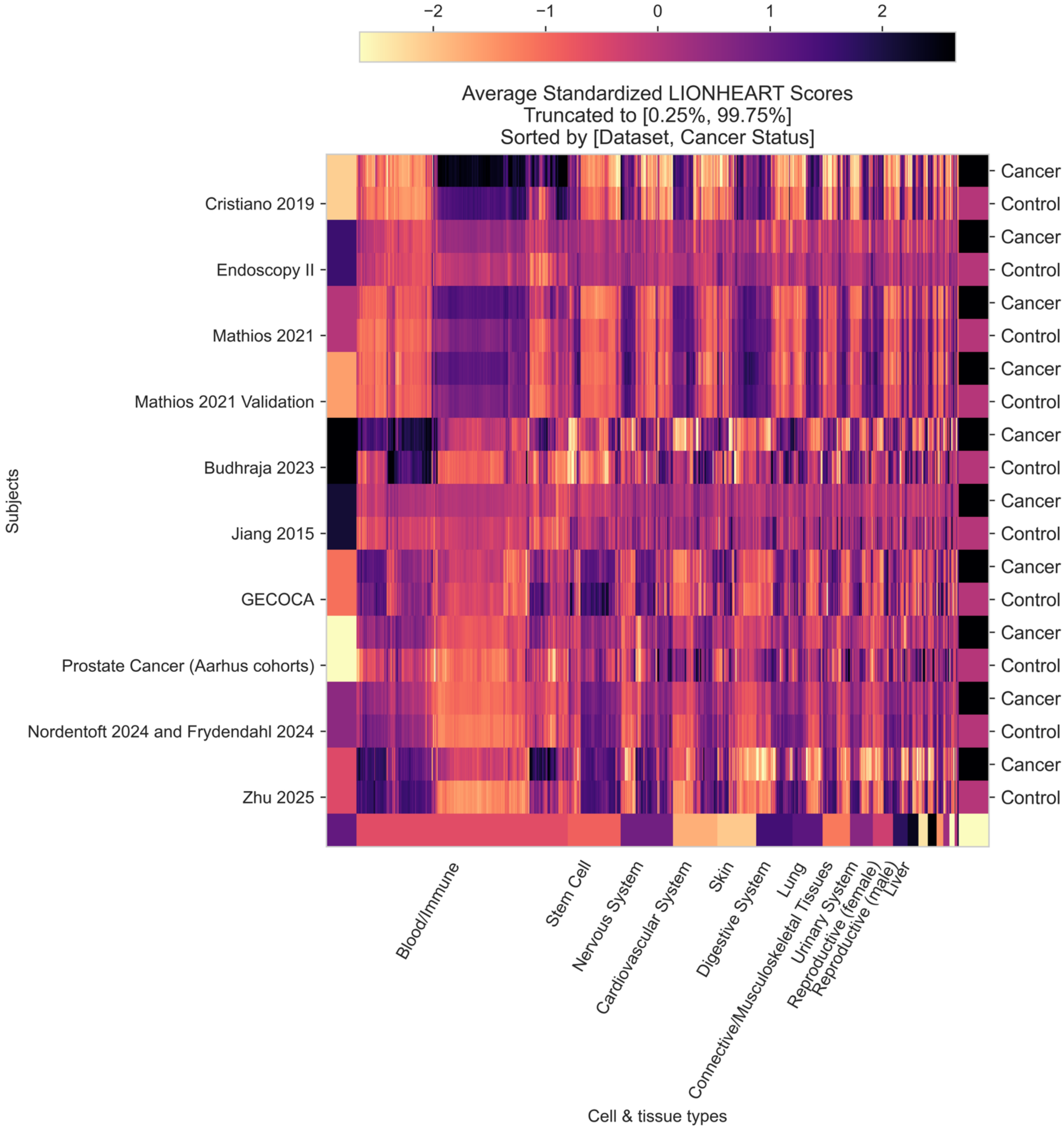


**Supplementary Figure 22**: Average standardized LIONHEART scores. Every row has the average scores of the samples in a given dataset and phenotype (cancer/control), and every column is a cell type. Extreme outliers were truncated for visualization purposes.

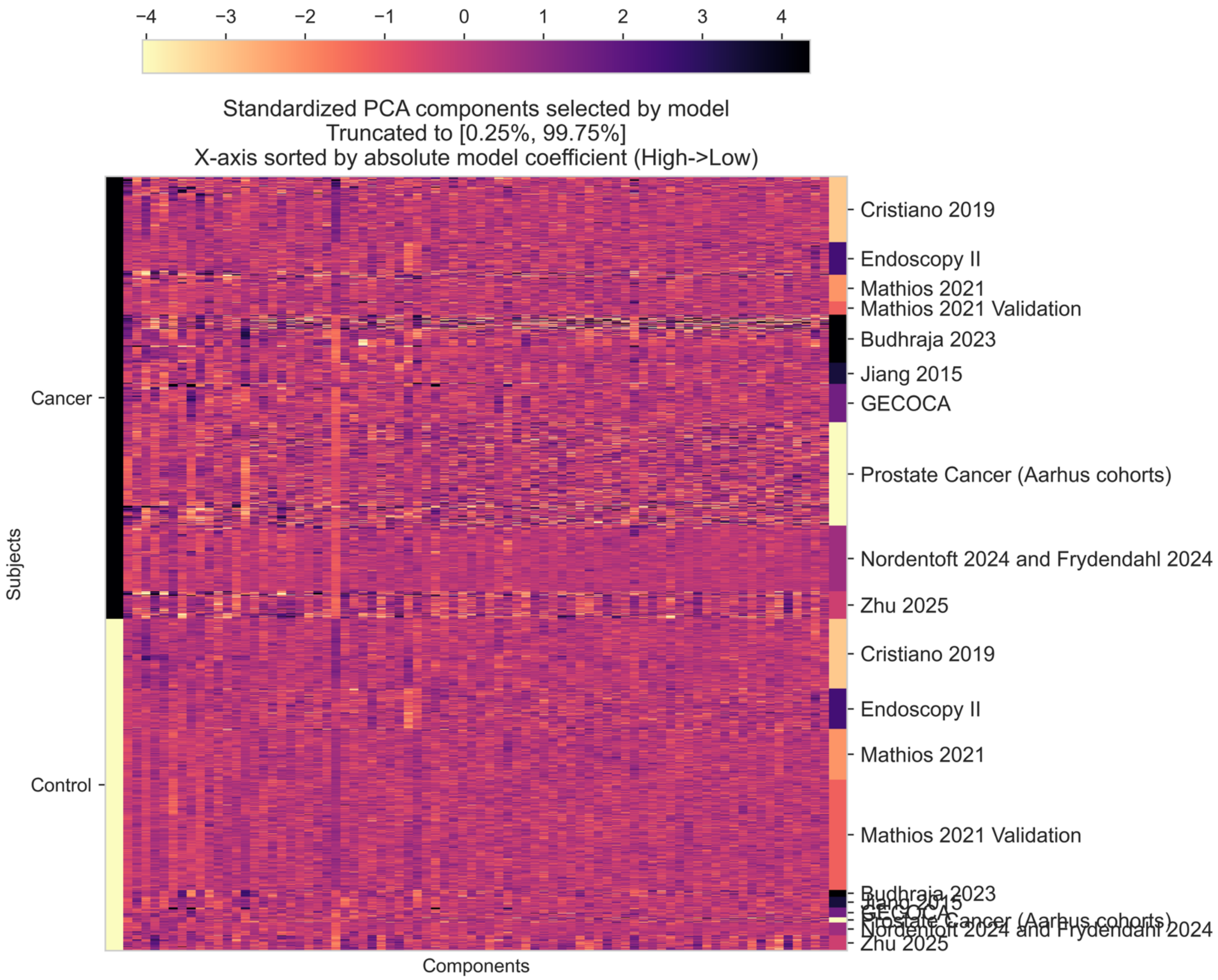


**Supplementary Figure 23:** The standardized PCA-transformed LIONHEART scores for the 78 components selected by the final model via LASSO (i.e., an absolute model coefficient greater than 1e-5). Extreme outliers were truncated for visualization purposes. Every row is a sample, and every column is a principal component. The x-axis is sorted by the highest to lowest absolute model coefficients.
