## Supplementary Note 1 for "Cross-dataset pan-cancer detection: Correlating cell-free DNA fragment coverage with open chromatin sites across cell types"

### Average Overlapping Insert Size Correction

*Ludvig Renbo Olsen, Nov. 2024*

In this note, we introduce the "average overlapping insert size correction" integrated in the calculation of **LIONHEART** scores.

The code to calculate and apply the correction is available in the `lionheart` python package using:

```
from lionheart.features.correction import
calculate_insert_size_correction_factors, correct_bias
```

#### Correction of cfDNA sequencing coverage

A core concept of LIONHEART is to increase the generalization of fragmentomics-based machine learning models across datasets (cohorts, labs) to improve the likelihood of future successful clinical implementation. One approach is to reduce the differences ("biases") between the datasets on variables hypothesized to be independent from the main signal (the main signal being the sequencing coverage reductions in accessible chromatin regions of cell types present in cfDNA).

A common bias is the GC bias, where we often see reduced sequencing coverage for fragments with more extreme GC fractions (i.e., $\frac{G+C}{G+C+A+T}$, where each variable represents a base count). The GC bias vary by sample and dataset and is influenced by technical processing steps such as PCR amplification. When correcting this bias, features often seem more biologically meaningful and similar across samples.

Before explaining the average overlapping insert size bias correction, let's go through an example of GC bias for one sample per dataset:

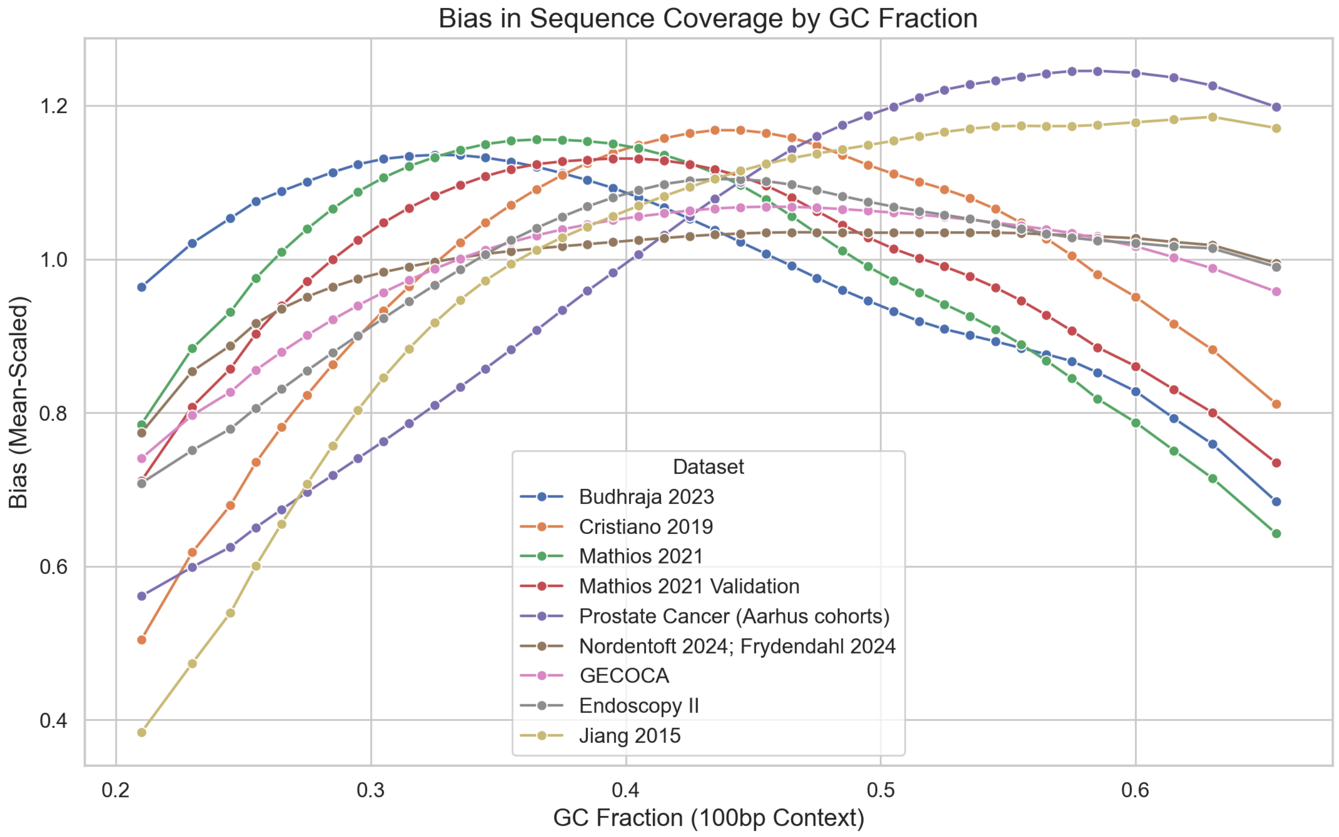


**Supplementary Note 1, Figure 1**: GC biases in cfDNA samples. Each line is the bias of a single sample.

#### GC correction

The GC bias is measured as the average sequencing coverage for each interval of GC fractions in the 100bp context around each 10bp bin. We divide the vector of average coverages with its own mean to get a "correction factor" vector centered around 1, as this will keep the values on the same scale after correction. To correct the bias, we divide each coverage value by the correction factor of its respective GC fraction:

---

Let the data be split into GC-content bins labelled $b = 1, 2, \ldots, B$ with each coverage value $j$ (i.e. 10bp bin) belonging to one $b$.

##### Calculate the mean coverage in each GC bin

$$\bar{c}_b = \frac{1}{n_b} \sum_{j \in \text{bin } b} c_j$$

##### Calculate the average mean coverage

$$\bar{c}_{\text{mean}} = \frac{1}{B} \sum_{b=1}^{B} \bar{c}_b$$

##### Compute the correction factor for each bin

$$f_b = \frac{\bar{c}_b}{\bar{c}_{\text{mean}}}$$

##### Apply the correction to every coverage value

For every $j$ in bin $b$,

$$c'_j = \frac{c_j}{f_b}$$

| Symbol | Meaning |
|---|---|
| $c_j$ | raw coverage value $j$ |
| $n_b$ | number of coverage values in bin $b$ |
| $\bar{c}_b$ | mean coverage in bin $b$ |
| $\bar{c}_{\text{mean}}$ | mean of the $\bar{c}_b$ values across all $B$ bins |
| $f_b$ | correction factor for bin $b$ |
| $c'_j$ | GC-corrected coverage value $j$ |

---

After this correction, the GC bias will simply be flat:

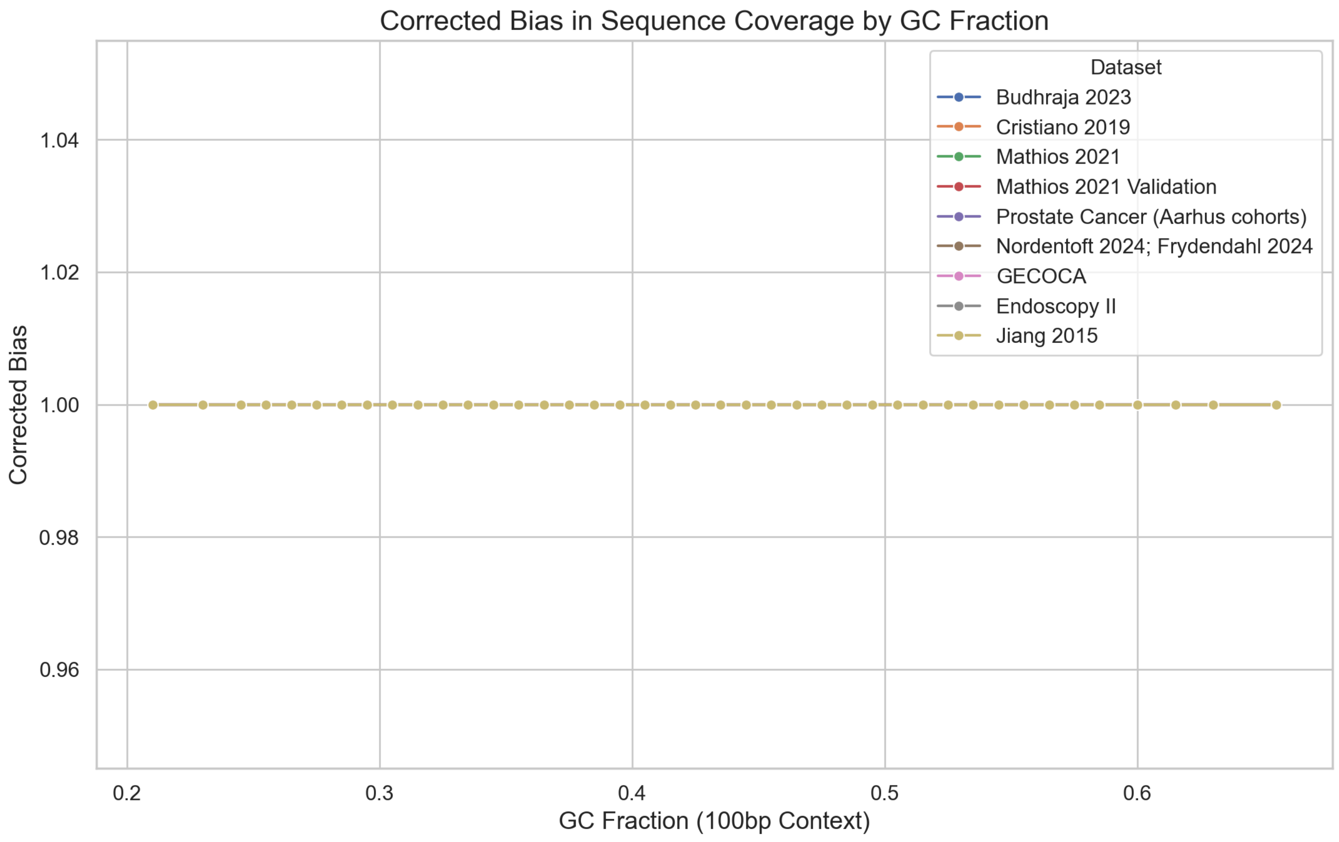

**Supplementary Note 1, Figure 2**: The corrected (flattened) GC bias.

#### Average Overlapping Insert Size Bias

While we know that the global fragment length distribution can be used to detect cancer, there are also dataset-specific changes in the distribution that can reduce generalization between datasets. For instance, we can observe different filtering of short and long fragments based on the applied bead ratio and choice of sequencer.

While investigating these differences, we discovered a bias in the fragment length distributions *overlapping* each 10bp coverage bin. We hypothesized that reducing this bias could increase the model generalization.

Reducing this bias may also remove useful cancer signal from the fragment lengths, but since this is not the signal we seek to quantify (i.e., the chromatin accessibility effect on coverage), removing it could actually improve the interpretability of LIONHEART scores. If we later want to include the fragment length cancer signal in the classifier, we can always add specific length features. The same point can be made for our normalization of copy number alterations, where we would rather add separate features that specifically capture those cancer signals. We can think of this approach as a sort of "deconvolution" of the cancer signals, for increased model interpretability.

Let's look at the biases for a single sample per dataset:

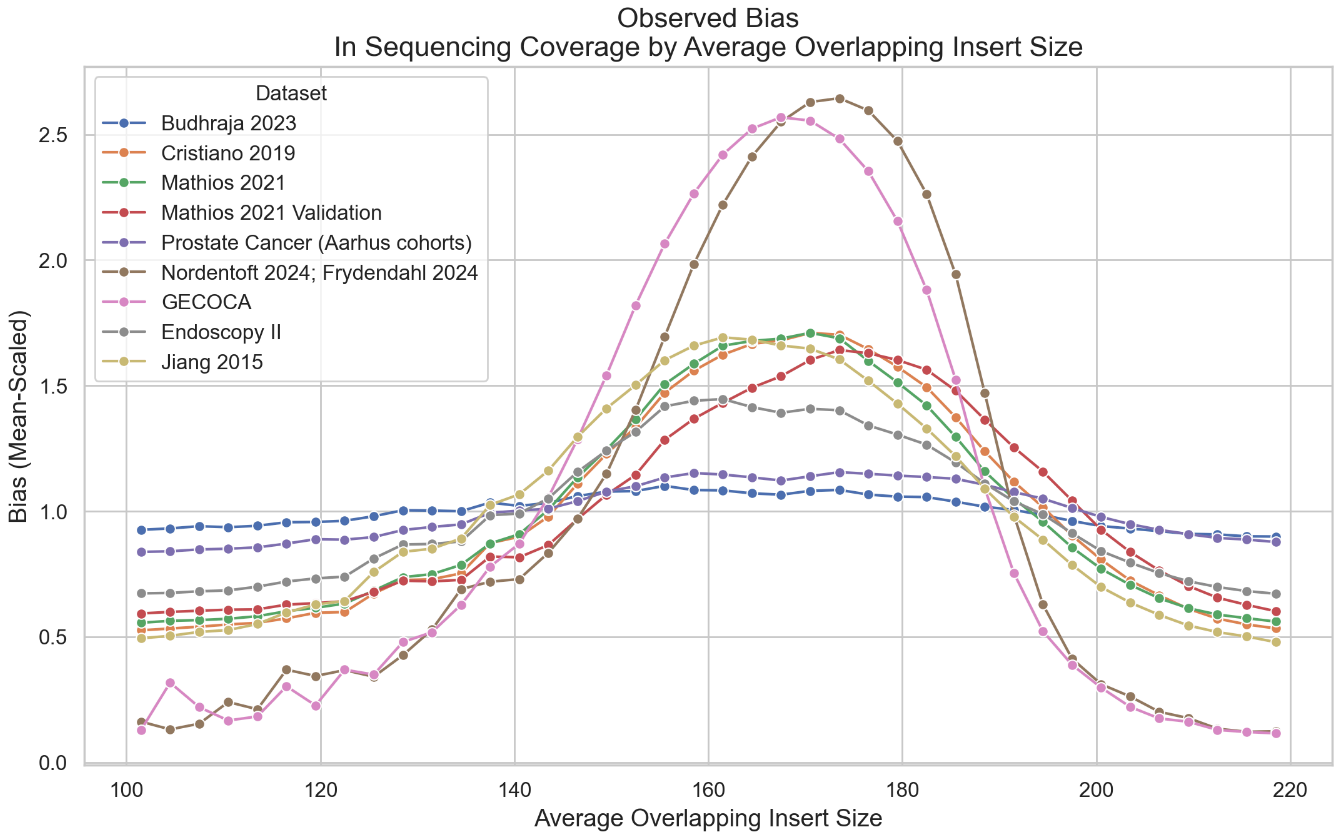


**Supplementary Note 1, Figure 3**: The observed bias in sequencing coverage by average overlapping insert size for a single sample per dataset.

The biases seem to differ between the datasets in a few dimensions:

- Differences in spread (i.e., the height of the distribution)
- Shifts in the peak positions (i.e., the distribution means are at different average overlapping insert sizes (x-axis))
- Differences in skewness

The differences in spread should mostly be due to the sequencing depth and the Central Limit Theorem [ https://en.wikipedia.org/wiki/central_limit_theorem ]. The higher the depth, the closer the average overlapping insert size of a bin will be to the global mean. Since this difference is a statistically expected effect of sequencing depth and not an actual bias in the coverage values, we don't want to "correct" this. In fact, we need to ensure that our transformations respect this effect (more on this in a minute).

Based on **Supplementary Figure 15**, both the shifts in peak position (mean values) and the skewness of the distributions seem to be dataset-specific. Let's see the average mean and skewness values for all samples per dataset (chromosome 1):

| | Dataset | Skewness | | Mean Insert Size | |
|---|---|---|---|---|---|
| | | mean | std | mean | std |
| 0 | Budhraja 2023 | −0.17 | 0.16 | 167.11 | 5.82 |
| 1 | Cristiano 2019 | −0.25 | 0.10 | 172.25 | 4.64 |
| 2 | Endoscopy II | −0.22 | 0.07 | 173.23 | 4.05 |
| 3 | GECOCA | −0.29 | 0.08 | 175.25 | 3.66 |
| 4 | Jiang 2015 | −0.15 | 0.12 | 166.38 | 6.25 |
| 5 | Mathios 2021 | −0.30 | 0.09 | 175.42 | 4.46 |
| 6 | Mathios 2021 Validation | −0.31 | 0.06 | 175.41 | 3.20 |
| 7 | Nordentoft 2024; Frydendahl 2024 | −0.59 | 0.24 | 172.99 | 4.77 |
| 8 | Prostate Cancer (Aarhus cohorts) | −0.04 | 0.04 | 167.89 | 3.77 |

**Supplementary Note 1, Table 1**: The means and standard deviations of the estimated mean and skewness parameters for all samples per dataset. The mean average overlapping insert size ranges from ~166bp to ~175bp, while the skewness values range from -0.59 to -0.04.

We set out to 1) remove the skewness and 2) enforce the same mean average overlapping insert size for all samples while 3) only minimally impacting the spread.

Let's go through the three transformations we performed to achieve this.

#### Estimating Mean and Skewness

First, let's look a bit closer at how we estimated the mean and skewness values in the first place. To do this, we need to understand the effect of the Central Limit Theorem on our bias:

For each 10bp bin, we have *some* number of overlapping fragments (i.e., the coverage). According to the Central Limit Theorem, the more fragments overlap a bin, the closer the average insert size should be (on average) to the global average overlapping insert size. (...and breathe...). So, if 10 fragments overlap a 10bp bin, we expect the average of their insert sizes to be closer to the overall average insert size than if only 2 fragments overlap the bin.

Since the coverage vary per 10bp bin, the effect of the Central Limit Theorem is different per 10bp bin (or rather, per unique number of observed overlapping fragments).

Let's visualize some (simulated) fragments and how the number of overlapping fragments changes the *average overlapping insert size* value in 10bp bins:

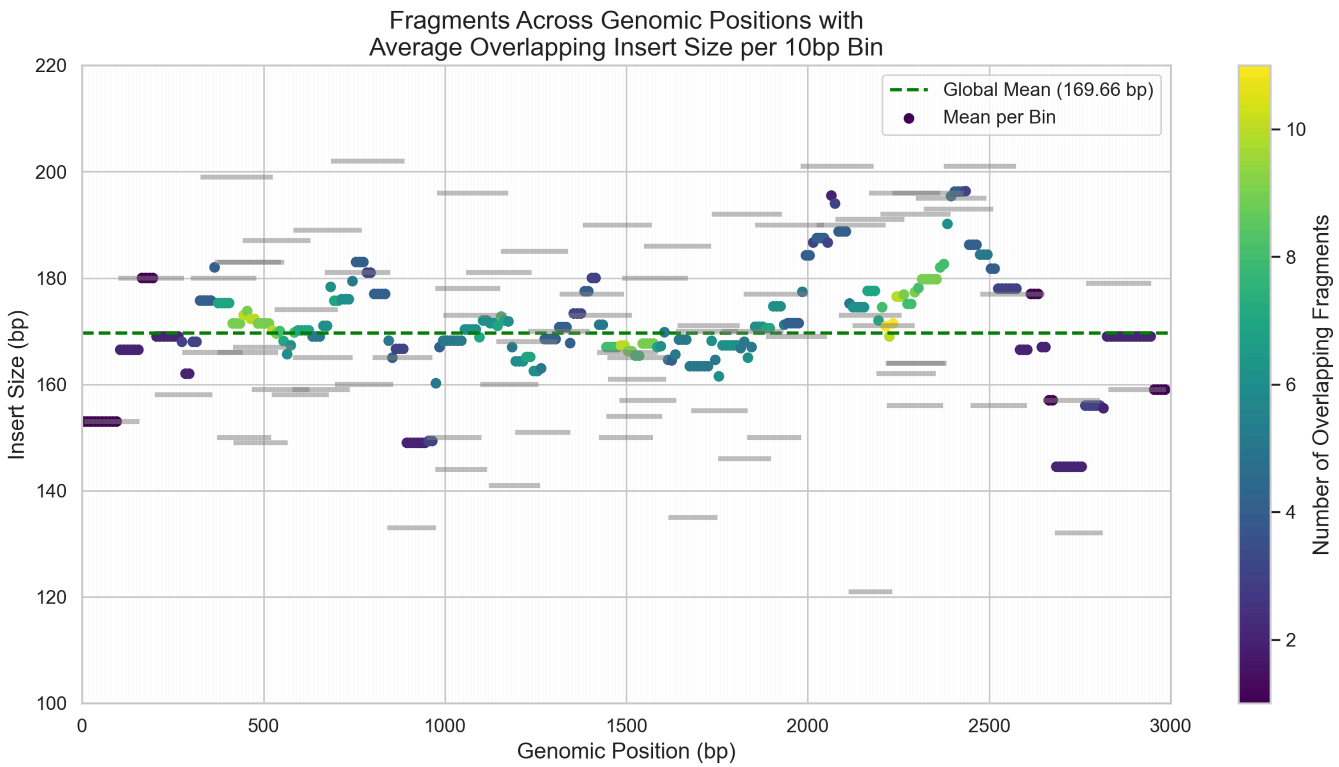


**Supplementary Note 1, Figure 4**: Fragments distributed throughout a genomic region (simulated data). The more fragments overlap, the closer the average overlapping insert size is to the global mean.

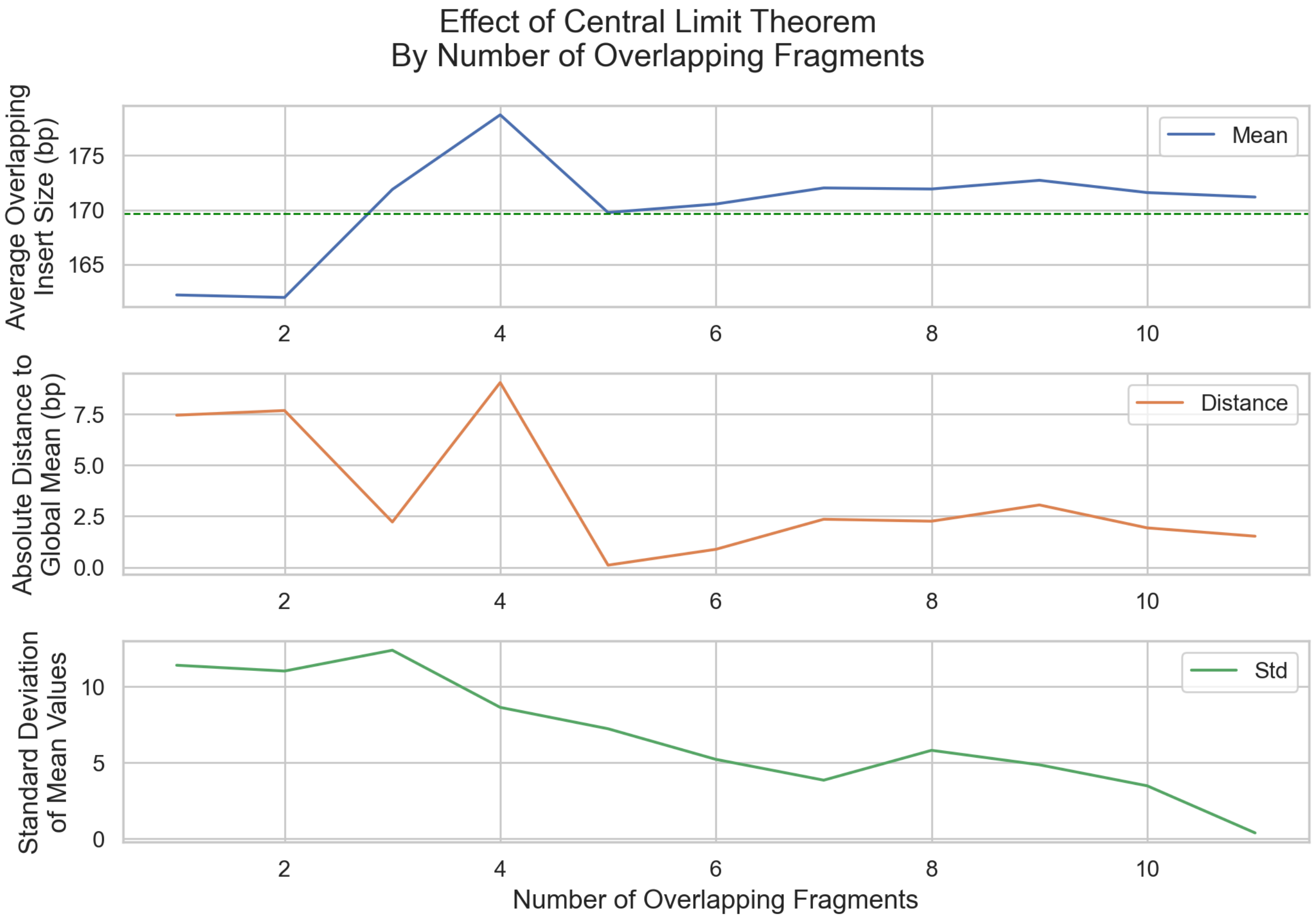


**Supplementary Note 1, Figure 5**: Metrics about the average overlapping insert size given a number of overlapping fragments. With more overlapping fragments, the average gets closer to the global mean, and the standard deviation of the averages is reduced.

In these plots, we see how the average values generally tend towards to the global mean when the number of overlapping fragments increases. *Since this is a small sample, the effect is not perfect, but it will be on a whole genome scale*.

So, the Central Limit Theorem affects each bin differently. How do we then fit a distribution to the observed bias to get the mean and skewness values?

Here is the observed bias for a single sample:

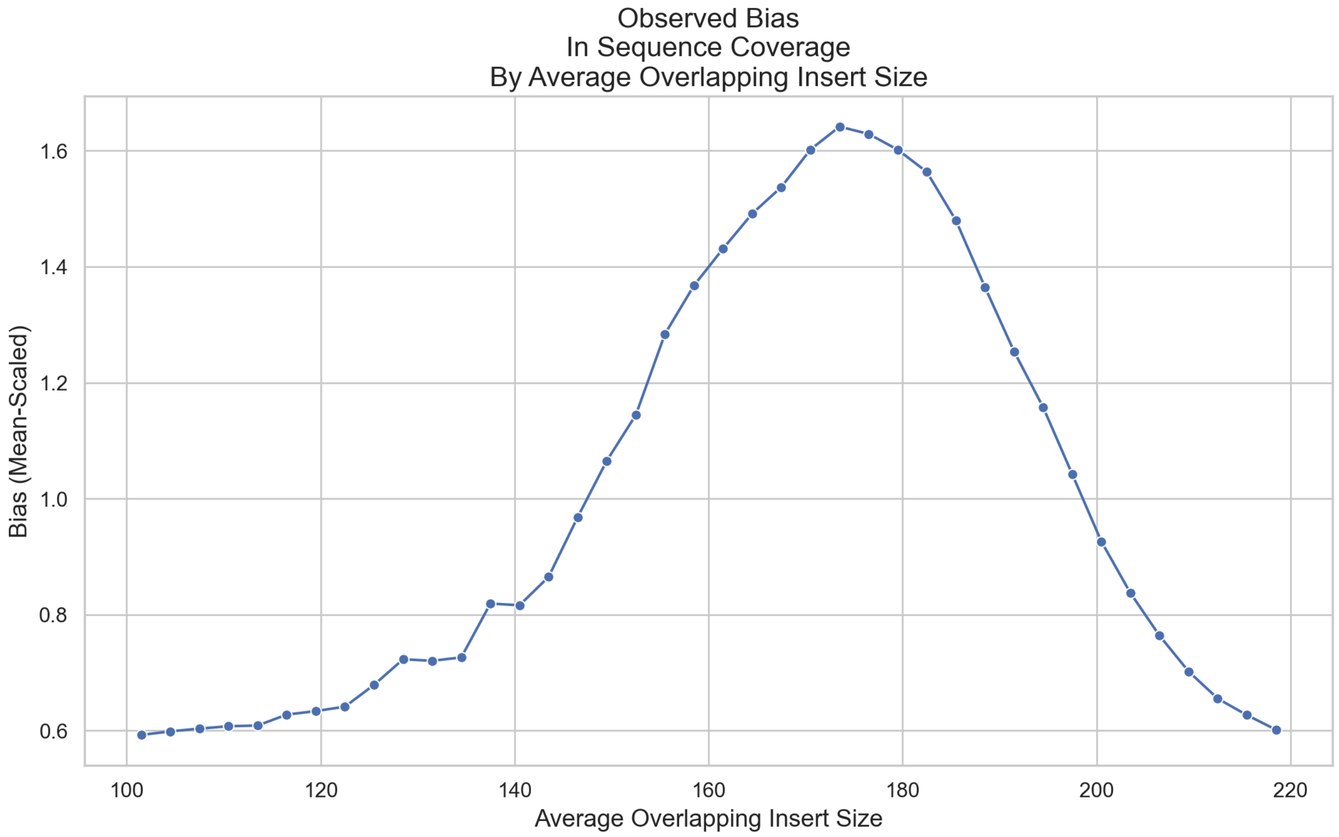


**Supplementary Note 1, Figure 6**: The observed bias for a single cfDNA sample.

We first want to fit a distribution that tells us the skewness of the observed bias. This can be done with a skewed student's t distribution, truncated to the possible values [100bp, 220bp].

As mentioned, we expect the spread of the distribution to mostly be an effect of the Central Limit Theorem. And since this effect works on a per-bin basis, we will want to model this effect for each unique number of overlapping fragments (i.e. coverage values) in our data.

**Mixture of Skewed Student's t Distributions**: In line with the Central Limit Theorem, the bin-wise spread of the bias distribution should be inversely proportional to the square root of the coverage depth. To account for this, we fit a mixture of skewed student's t distributions (one per unique coverage value) using a spread appropriately scaled by the coverage value. It's important to note that all distributions *share the optimized mean and skewness parameters*. Finally, we sum the distributions, weighted by the frequency of each coverage value.

Let's see the fitted distribution on top of the observed bias:

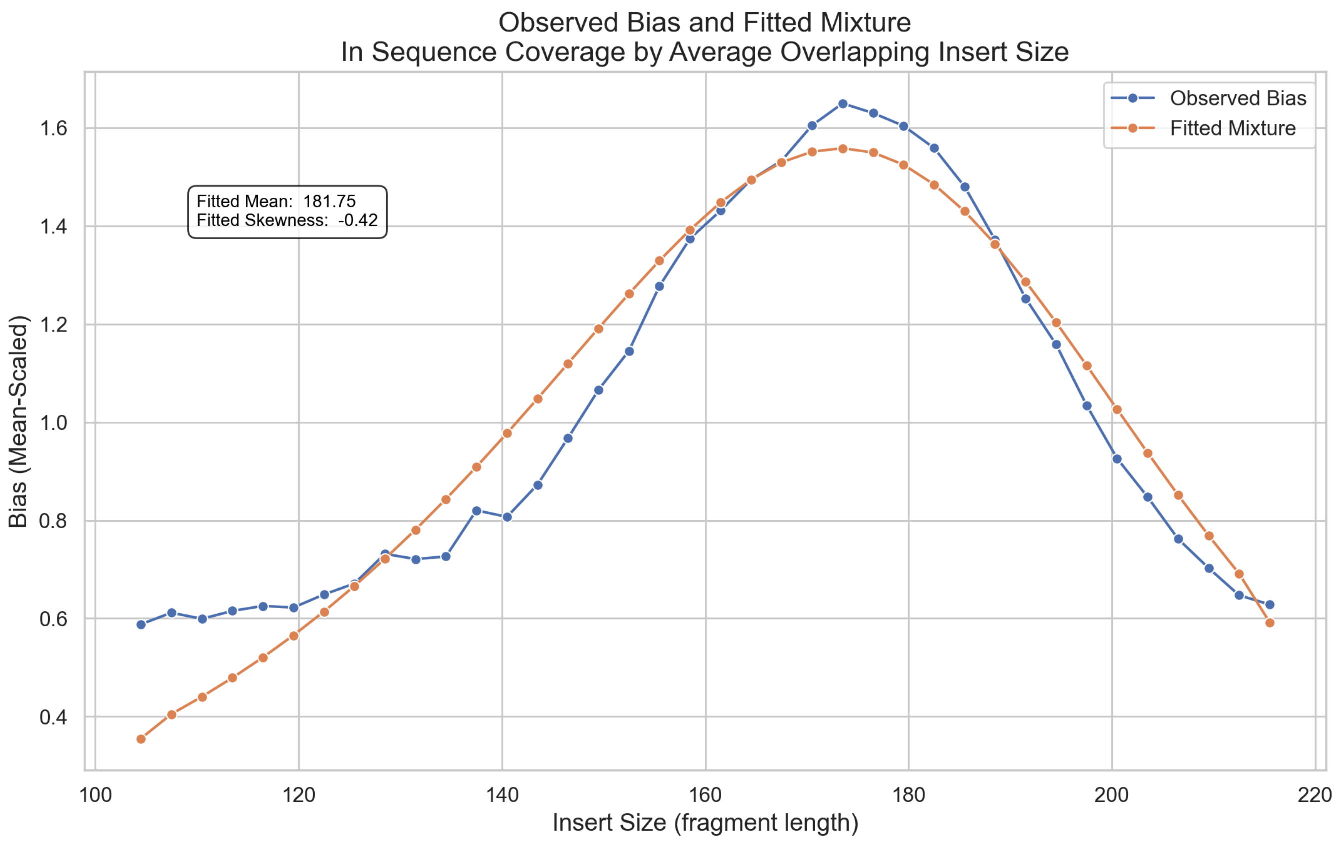


**Supplementary Note 1, Figure 7**: The fitted mixture distribution and the observed bias.

#### Step 1 - Removing "Noise" and Skewness

Based on this fitted mixture distribution, we will perform two transformations to the coverages:

1. Remove the difference between the observed and fitted distributions. For lack of a better word, we refer to this as "noise". Removing these differences will allow us to get a more precise mean value later.
2. Remove the skewness from the bias distribution.

##### Noise Correction

The first transformation divides each coverage value by the relative difference between the observed and fitted distributions for its average overlapping insert size. We used this same principle in the GC correction.

---

Insert-size bins are labelled $b = 1, 2, \ldots, B$ (each bin spans 3 bp) with each coverage value $j$ (i.e. 10bp bin) belonging to one $b$. Due to the GC correction, we start with the "once-corrected" coverage.

Let $t'_b$ denote the expected coverage for bin $b$ from the $t$-mixture model.

#### Calculate the mean once-corrected coverage in each bin

$$\bar{c}'_b = \frac{1}{n_b} \sum_{j \in \text{bin } b} c'_j$$

##### Compute the noise-correction factor for each bin

$$g_b = \frac{\bar{c}'_b}{t'_b}$$

##### Apply the correction to every coverage value

For every $j$ in bin $b$,

$$c''_j = \frac{c'_j}{g_b}$$

| Symbol | Meaning |
|---|---|
| $c'_j$ | coverage value $j$ *after* the previous correction |
| $n_b$ | number of coverage values in insert-size bin $b$ |
| $\bar{c}'_b$ | mean of the $c'_j$ values in bin $b$ |
| $t'_b$ | fitted $t$-mixture mean for bin $b$ |
| $g_b$ | noise-correction factor for bin $b$ |
| $c''_j$ | coverage value $j$ *after* noise correction |

---

Skewness Correction

For the second transformation, we calculate a correction factor that removes the skewness from the bias when we divide the coverages by it:

---

Insert-size bins are labelled $b = 1, 2, \ldots, B$ (each bin spans 3 bp) and have mid-point sizes $s_b$ (in bp). Each coverage value $j$ (i.e. 10bp bin) belong to one $b$.

##### Compute the mean insert size across bins

$$\bar{s} = \frac{1}{B} \sum_{b=1}^{B} s_b$$

##### Compute the skewness-correction factor for each bin

$$h_b = \bar{s}\,(1 - \gamma) + 1 + \gamma\, s_b$$

##### Apply the correction to every coverage value

For every $j$ in bin $b$,

$$c_j''' = \frac{c_j''}{h_b}$$

| Symbol | Meaning |
|---|---|
| $c_j''$ | coverage value $j$ *after* the previous (noise) correction |
| $s_b$ | mid-point insert size of bin $b$ (bp) |
| $\bar{s}$ | mean of the $s_b$ values across all $B$ bins |
| $\gamma$ | skewness coefficient of the insert-size distribution |
| $h_b$ | skewness-correction factor for bin $b$ |
| $c_j'''$ | coverage value $j$ *after* skewness correction |

---

Let's visualize these two correction factors to see what the coverages are divided by (given the average overlapping insert size of the bin):

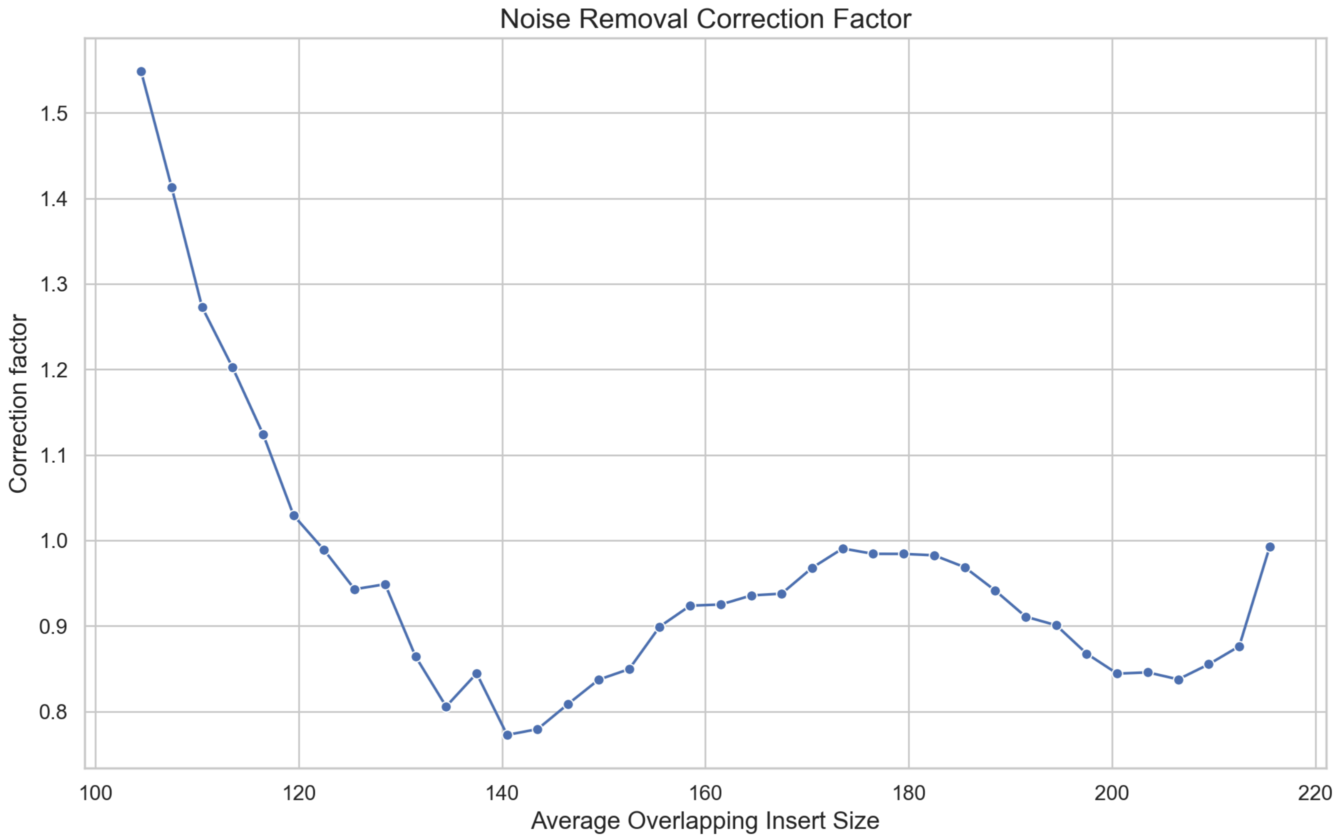


**Supplementary Note 1, Figure 8**: The correction factor for removing the "noise" from the bias.

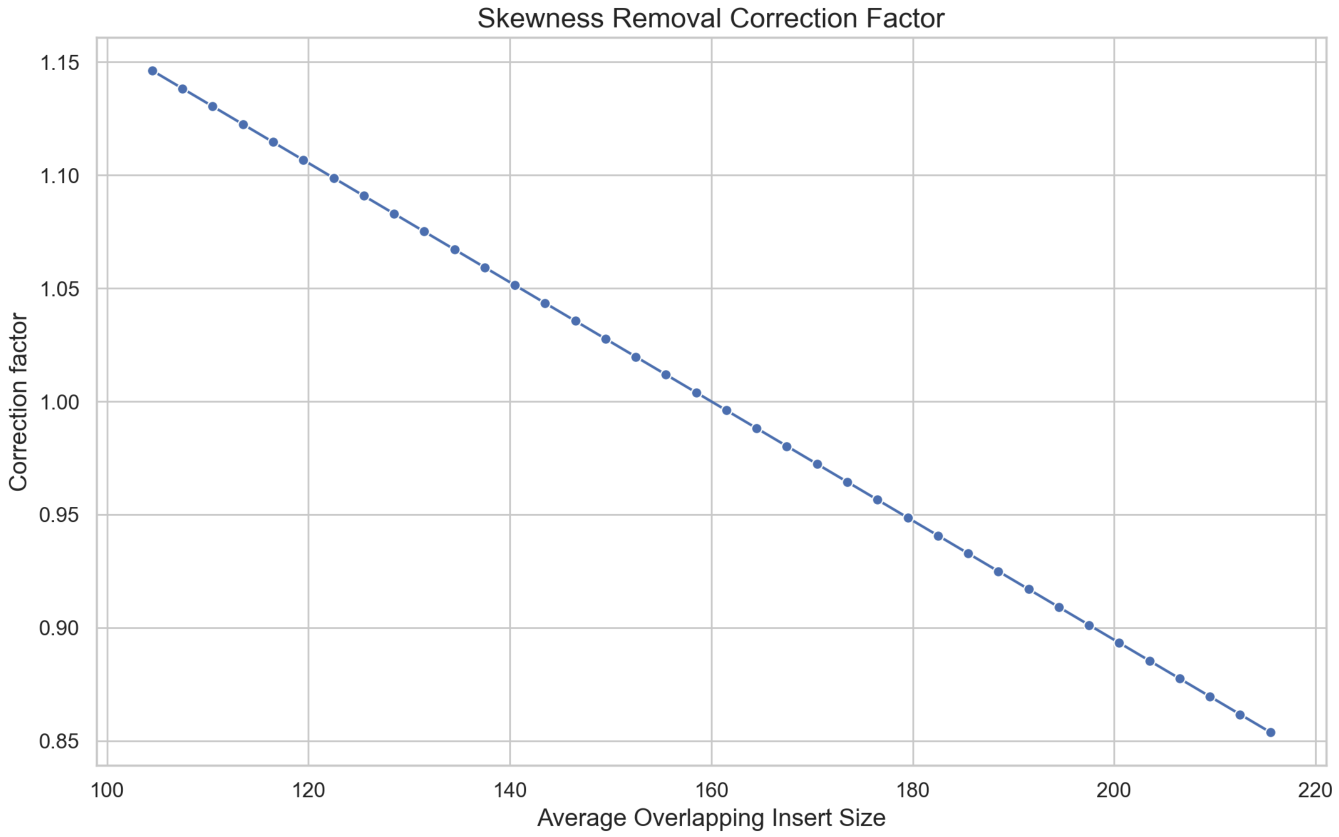


**Supplementary Note 1, Figure 9**: The correction factor for removing the skewness from the bias.

After applying these corrections, the bias now looks like this:

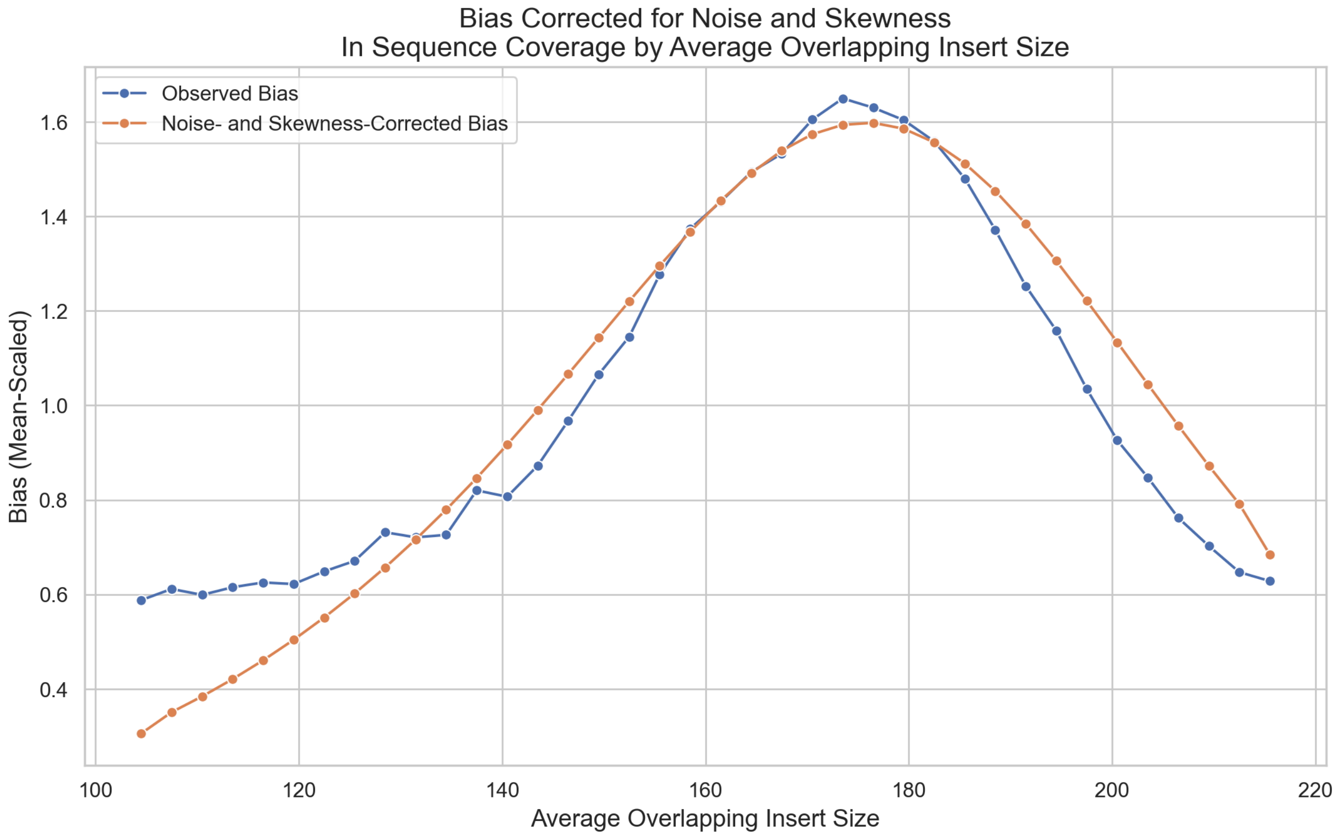


**Supplementary Note 1, Figure 10**: The noise- and skewness-corrected bias and the initially observed bias.

#### Step 2 - Shifting the Mean

Having removed the "noise" and skewness, we now want to shift the mean of the distribution to 166bp.

After removing the skewness, the mean of the bias in the coverages will have changed. Hence, we refit the mixture of student's t distributions on the corrected coverages:

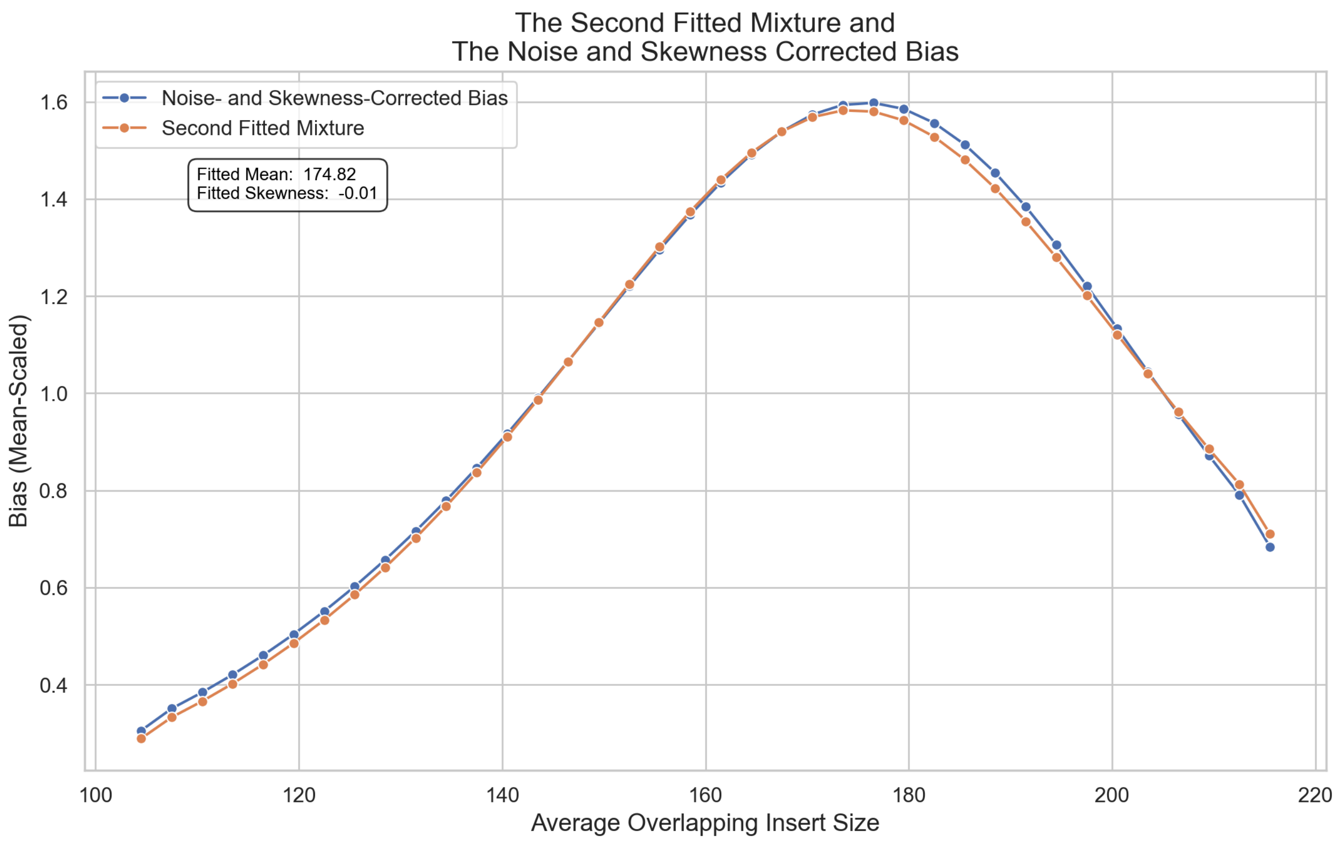


**Supplementary Note 1, Figure 11**: The (2nd) fitted mixture distribution on top of the noise- and skewness-corrected bias. Whereas the originally observed bias had a mean of ~181bp, the new fitted mixture has a mean of ~174bp.

As with the noise and GC corrections, we can create the mean shift by simply dividing each (thrice-corrected) coverage value by the relative difference of the noise- and skewness-corrected bias and the second fitted mixture distribution ***with its mean set to 166bp***:

---

Insert-size bins are labelled $b = 1, 2, \ldots, B$ (each bin spans 3 bp) with each coverage value $j$ (i.e. 10bp bin) belonging to one $b$.

Let $t_b^{\prime\prime(166)}$ denote the expected coverage for bin $b$ from the **second** $t$-mixture model, with the global mean fixed at 166 bp.

#### Calculate the mean thrice-corrected coverage in each bin

$$\bar{c}_b^{\prime\prime\prime} = \frac{1}{n_b} \sum_{j \in \text{bin } b} c_j^{\prime\prime\prime}$$

#### Compute the mean-shift correction factor for each bin

$$m_b = \frac{\bar{c}_b^{\prime\prime\prime}}{t_b^{\prime\prime(166)}}$$

##### Apply the correction to every coverage value

For every $j$ in bin $b$,

$$c_j'''' = \frac{c_j'''}{m_b}$$

| Symbol | Meaning |
|---|---|
| $c_j'''$ | coverage value $j$ *after* the skewness correction |
| $n_b$ | number of coverage values in bin $b$ |
| $\bar{c}_b'''$ | mean of the $c_j'''$ values in bin $b$ |
| $t_b''^{(166)}$ | expected coverage for bin $b$ from the second fitted $t$-mixture ($\mu = 166$ bp) |
| $m_b$ | mean-shift correction factor for bin $b$ |
| $c_j''''$ | coverage value $j$ *after* mean-shift correction |

---

The mean-shift correction factor looks as follows:

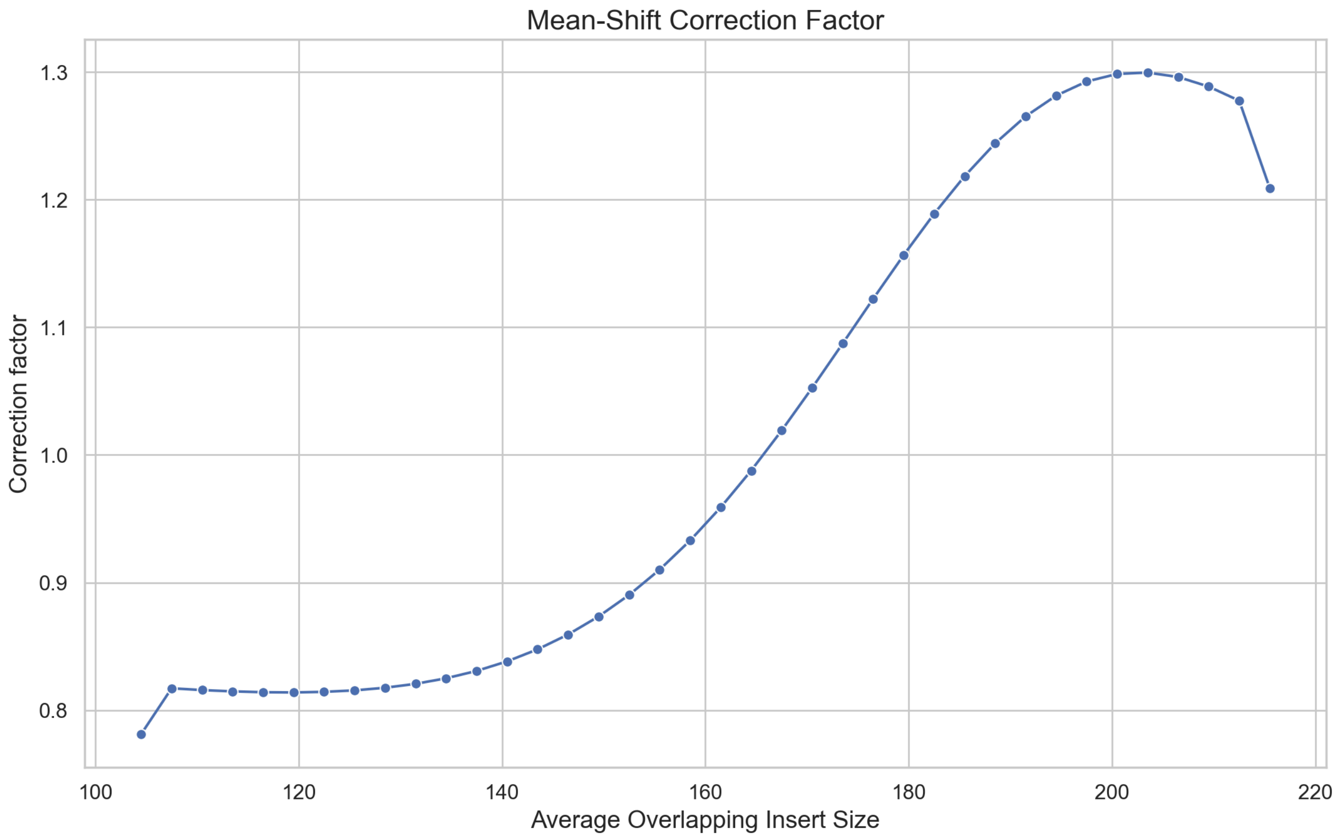


**Supplementary Note 1, Figure 12**: The correction factor for shifting the distribution mean to 166bp.

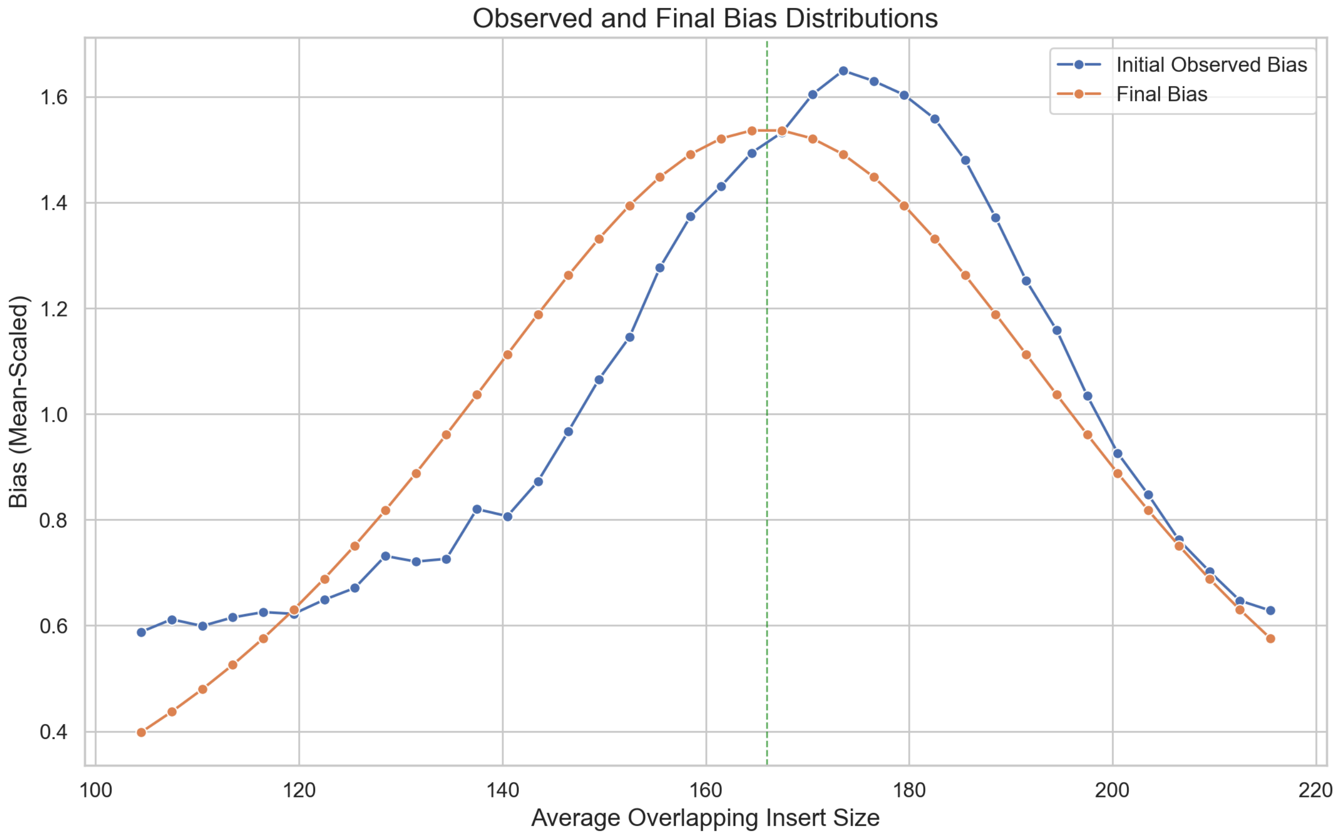


**Supplementary Note 1, Figure 13**: The final bias distribution on top of the initially observed bias distribution.

### Before and after for all samples

When only observing a single sample, it can be difficult to imagine the usefulness of applying these corrections to the coverage values. As the entire point is to make the samples and datasets more similar (for increased model generalization), we should instead watch the effect of the corrections on multiple samples.

**Supplementary Figure 15** (in the main supplementary materials file) shows the observed and final ("target") biases, as well as the 3 correction factors, for all samples.

Here, we will compare the samples we initially plotted before and after the corrections:

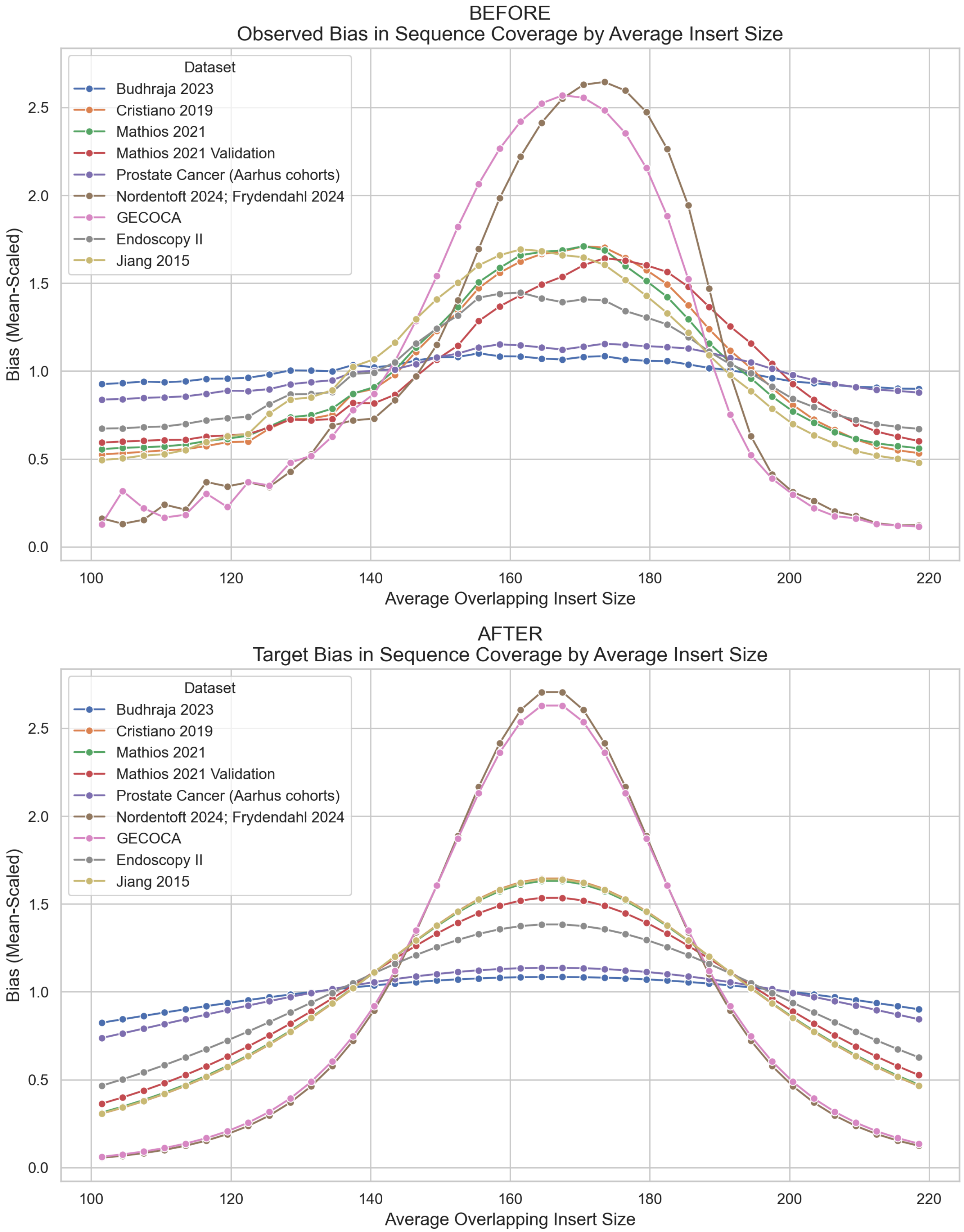


**Supplementary Note 1, Figure 14**: The observed and target biases for one sample per dataset.

After the three corrections, the bias distributions are more similar between the datasets, while leaving the spread (peakiness) reasonably close to the original distributions.
